# Alcohol, Cardiovascular Disease, and Industry Influence: A Meta-review

**DOI:** 10.64898/2026.03.18.26348685

**Authors:** Su Golder, Oscar Lau, Greg Hartwell, Laurence Blanchard, Anna Gibson, Catriona Crookes, Lowri Foster Davies, Rebecca E Glover

**Author notes:** Corresponding Author: Su Golder BSc (hons), MSc, FRSA, PhD, Associate Professor, Department of Health Sciences, University of York, York, YO10 5DD. Funding: This project (18037) is funded by the Better Methods, Better Research (BMBR) programme, an MRC and NIHR partnership. The views expressed in this publication are those of the author(s) and not necessarily those of the MRC, NIHR or the Department of Health and Social Care. Ethical Approval: Ethical approval was not required for this project, as only publicly available secondary data was used.

## Abstract

**Objectives:** This meta-review compares methodological and reporting approaches between systematic reviews examining alcohol dose and cardiovascular disease (CVD) and assesses whether alcohol industry involvement is associated with divergent conclusions.

**Methods:** KSR Evidence was searched 6 May 2025 to update a cohort of 60 systematic reviews from previous review. Reviews were included if they examined any dose-response relationship between alcohol consumption and CVD. Two reviewers independently screened records and extracted data on review characteristics, and citations. Methodological quality was appraised using AMSTAR 2. For a matched sample of reviews with and without known alcohol industry funding, the overlap of included primary studies was compared using Corrected Covered Area (CCA) analysis.

**Results:** Thirty additional systematic reviews met the inclusion criteria, yielding 90 systematic reviews (1996–2025). Most (60.0%, 54/90) concluded that alcohol had a cardioprotective effect, whereas 31.1% (28/90) concluded no evidence of protection, and 8.9% (8/90) were inconclusive. Twenty reviews (22.0%) had declared or inferred alcohol industry funding or author connection; all but one reported a protective effect at lower doses, the other was inconclusive.

Industry-connected reviews were cited more often (mean 575.9 vs 193.0, p=0.0002) and more commonly examined overall CVD rather than specific conditions (such as hypertension or stroke). Study overlap was low (CCA 2.59%) and 99% of reviews were rated as critically low quality.

**Conclusions:** The fragmented evidence base is of poor methodological quality with selective inclusion of studies. Alcohol industry connections are strongly associated, with conclusions favouring alcohol consumption, highlighting the need for independent high-quality systematic reviews.

## Introduction

Alcohol consumption plays a causal role in over 200 diseases, injuries and other health conditions, and has been described as an entrenched economic, cultural, and social construct (Naimi et al., 2013). It is a leading risk factor for global disease burden(Bryazka et al., 2022), with risk increasing with consumption. Despite this, alcohol’s overall association with health is complicated by long-standing uncertainty regarding the potential cardioprotective effects of low to moderate consumption (Oppenheimer & Bayer, 2020), often referred to as a ‘J-curve’, which has led to confusion among both consumers and decision-makers (Whitman et al., 2015). These conflicting findings are not only evident in primary research but in secondary analysis such as systematic reviews and meta-analysis (McCambridge & Golder, 2024). This inconsistency is particularly striking in systematic reviews on the topic, given the rigorous methods and predefined protocols that they are expected to follow(Higgins JPT, 2024). In principle, systematic reviews addressing the same questions should yield similar results due to overlap in included studies, methods and analysis (Higgins JPT, 2024). Systematic reviews are also widely regarded as the ‘gold standard’ of evidence base in public health (Dekkers et al., 2019; Higgins JPT, 2024), and are used to inform guidelines and health policy, making their conclusions even more salient.

Systematic reviews on alcohol and cardiovascular health are characterised by a high degree of discordance. In our previous study of 60 systematic reviews examining the impact of alcohol on cardiovascular health, we found controversy regarding the purported benefits of low to moderate alcohol consumption: 39 reviews reported some form of protective effect, two were inconclusive, and 19 concluded that even small amounts of alcohol were harmful (Golder & McCambridge, 2021).

While systematic reviews are expected to include all studies meeting their inclusion criteria, and not exclude studies based on their funding source unless flawed biases are identified, emerging evidence indicates that the alcohol industry plays a significant role in shaping this evidence base(Bartlett & McCambridge, 2021, 2023; McCambridge & Mialon, 2018; McCambridge et al., 2023; Mitchell, Lesch, & McCambridge, 2020). Reviews with alcohol industry funding or authors with a history of alcohol industry funding are more likely to report cardiovascular protective effects of low to moderate alcohol drinking compared with those without industry ties (Golder & McCambridge, 2021; McCambridge & Golder, 2024). This narrative then extends to websites of the alcohol industry and the social aspects/public relations organisations it funds (Peake et al., 2021).

The influence of the alcohol industry on scientific evidence extends beyond direct funding of individual studies(Bartlett & McCambridge, 2021, 2023; McCambridge & Mialon, 2018; Mitchell, Lesch, & McCambridge, 2020). It includes the design and framing of research questions, selective publication and citation practices, ghost writing, and the strategic use of systematic reviews to amplify favourable narratives (McCambridge et al., 2023). Collectively, these activities form part of broader corporate political strategies designed to shape policy-relevant science. Such practices can subtly bias the evidence synthesis process, leading to divergent interpretations even when methodological protocols are similar. Understanding how industry contributes to systematic reviews of alcohol consumption and CVD remains limited.

We undertook an in-depth methodological investigation in the form of a meta-review (Closs et al., 2016). Our aim was to update our previous cohort of systematic reviews on alcohol dose-response and CVD and use this larger, more current, cohort to compare the approaches employed in systematic reviews with and without alcohol industry connections.

## Methods

### Literature search and eligibility criteria

A literature search was conducted on the 6th of May 2025 to identify all systematic reviews examining the effects of a dose response relationship between alcohol intake and CVD published since our previous study. All CVD-related outcomes were examined, including coronary heart disease, atrial fibrillation, heart attack, congenital heart disease, heart failure and stroke as well as hypertension and biomarkers. To be included, studies needed to be a systematic review, or meta-analysis. All types of alcohol were considered. No limit was imposed on language and publication date.

The search was performed in KSR Evidence database via OVID using synonyms related to two key concepts: alcohol and CVD.

(alcohol* OR drinker* OR drinking OR beer OR wine OR spirits).ti,ab AND (Cvd OR Cardio* OR chd OR heart OR cardiac OR coronary OR myocard* OR angina OR ischemic attack OR ischaemic attack OR atrial OR aortic disease OR aortic aneurysm OR ventricular dysfunction OR mortality OR stroke OR intracerebral hemorrhage OR cerebrovascular OR stroke OR vascular OR blood pressure OR hypertension).ti,ab

All retrieved records were imported into Covidence for screening. Two reviewers independently screened titles and abstracts, with any discrepancies resolved through discussion, or consultation with a third reviewer. Full-text screening was conducted using the same approach.

### Data Extraction

Data extraction was conducted using Elicit.com and verified by two reviewers. Extracted information included: journal characteristics (journal title, article citation count and year of publication), review inclusion criteria (intervention - dose and type of alcohol beverage, outcome measures, number of included studies, study design of included studies, analytical approach, consideration of confounding factors including sick quitters (individuals who stop drinking alcohol because of health problems), and any subgroup analysis.

We also recorded each review’s conclusions as ‘for’ or ‘against’ a protective cardiovascular effect, as well as information on study support (declared funding sources, any stated conflicts of interests, and any known/unstated alcohol industry connections among authors). Information on individual authors’ alcohol industry connections was additionally identified through web searches using Google, the Web of Science suite of databases, and the social media platform LinkedIn.

We also selected the sample of systematic reviews with direct industry funding and general CVD outcomes and matched them by outcome to a sample of systematic reviews without known industry funding. For each review, we extracted the list of the included studies to compare and assess the degree of overlap (details below).

### Quality Assessment

The quality of the reviews was assessed using AMSTAR 2 (Shea et al., 2017), a validated and widely accepted tool for evaluating the methodological quality of systematic reviews, and suitable for facilitating cross-review comparisons.

### Analysis

The analysis was primarily descriptive and comparative, examining the differences between systematic reviews with and without known alcohol industry connections in relation to journal characteristics, inclusion criteria, types of analysis, citation metrics, and AMSTAR 2 rating.

With respect to comparing the included primary studies within the reviews we extracted the included studies list from the six systematic reviews funded by industry with general CVD outcomes and matched by outcome to the five reviews with no known industry funding with general CVD outcomes. We created a matrix for the included studies from each review, and then calculated the Corrected Covered Area (CCA) (Hennessy & Johnson, 2020) and the proportion of unique included studies in each review in this sample.

## Results

Our searches of KSR Evidence retrieved 968 records of which 868 were excluded at the title and abstract stage, and a further 69 were excluded at the full text screening stage (see Table S1: Excluded Studies and reasons for exclusion). The 30 newly identified systematic reviews were then amalgamated with the 60 reviews included in our previous network analysis which had the same search strategy and eligibility criteria(Golder & McCambridge, 2021). In total therefore, we identified 90 systematic reviews for inclusion which evaluated the impact of alcohol consumption dose response on CVD (Arafa et al., 2023; Bagnardi et al., 2008; Barbalho et al., 2020; Bouajila et al., 2024; Briasoulis, Agarwal, & Messerli, 2012; Brien et al., 2011; Britton et al., 2000; Cecchini et al., 2024; Chen et al., 2008; Cleophas, 1999; Colpani et al., 2018; Corrao et al., 2000; Costanzo et al., 2010, 2011; Del Giorno et al., 2022; Di Castelnuovo et al., 2006; Di Castelnuovo et al., 2002; Di Federico et al., 2023; Ding et al., 2021; Drogan et al., 2012; Estruch & Hendriks, 2021; Fuchs & Fuchs, 2021; Gallagher et al., 2017; Giannopoulos et al., 2022; Green et al., 2014; Grindal et al., 2022; Hrelia et al., 2022; Huang et al., 2014; Huang et al., 2017; Hwang, Piano, & Phillips, 2021; Jiang et al., 2022; Jung et al., 2020; Karpyak et al., 2014; Kelso, Sheps, & Cook, 2015; Khatiwada et al., 2025; Kodama et al., 2011; Koppes et al., 2006; Krittanawong et al., 2022; Larsson, Drca, & Wolk, 2014; Larsson, Orsini, & Wolk, 2015; Larsson, Wallin, & Wolk, 2018; Larsson et al., 2016; Lippi, Mattiuzzi, & Franchini, 2015; Liu et al., 2020; Luceron-Lucas-Torres et al., 2023; Marcos et al., 2021; Mazzaglia et al., 2001; McFadden et al., 2005; Mostofsky et al., 2016; Naame, Li, & Huang, 2019; O’Keefe et al., 2021; O’Neill et al., 2018; Okojie et al., 2020; Padilla, Michael Gaziano, & Djoussé, 2010; Patra et al., 2010; Peng et al., 2020; Rehm et al., 2017; Reynolds et al., 2003; Rimm et al., 1996; Rimm et al., 1999; Roerecke, Gual, & Rehm, 2013; Roerecke et al., 2017; Roerecke & Rehm, 2010, 2011, 2012, 2014a, 2014b; Roerecke et al., 2018; Ronksley et al., 2011; Samokhvalov, Irving, & Rehm, 2010; Sheng et al., 2024; Spaggiari et al., 2020; Spencer et al., 2017; Sperkowska et al., 2021; Stockwell et al., 2016; Tasnim et al., 2020; Taylor et al., 2009; van de Luitgaarden et al., 2021; Visontay et al., 2022; Weaver et al., 2020; Wilkens et al., 2021; Xin et al., 2001; Yang et al., 2021; Yang et al., 2016; Ye, Chen, & Bao, 2019; Yoon et al., 2020; Yuan et al., 2024; Zhang et al., 2014; Zhang et al., 2022; Zhang et al., 2015; Zhao et al., 2017; Zheng et al., 2015) (see Table S2: Characteristics of Included Studies).

### Review conclusions

These 90 systematic reviews came to opposing conclusions, with (60.0%, 54/90) identifying a protective effect in line with a J-shaped curve, 31.1% (28/90) identifying a harmful impact even at low doses, and 8.89% (8/90) being inconclusive or showing mixed results. Below, we disaggregate these differences by funding source.

### Alcohol industry funding and connections

Twenty systematic reviews (20/90, 22.2%) were found to have a connection with the alcohol industry, of which eight had a declaration of alcohol industry funding within the manuscript and a further twelve had authors with connections to industry, or who had received alcohol industry funding outside of the systematic review. All 20 reviews concluded a protective effect from low or moderate consumption with the exception of one which concluded mixed results (van de Luitgaarden et al., 2021) (Figure 1). This systematic review involved one author who obtained a PhD six years previously funded by the alcohol industry and concluded that they were unable to reach any conclusion(van de Luitgaarden et al., 2021).

**Figure 1:**
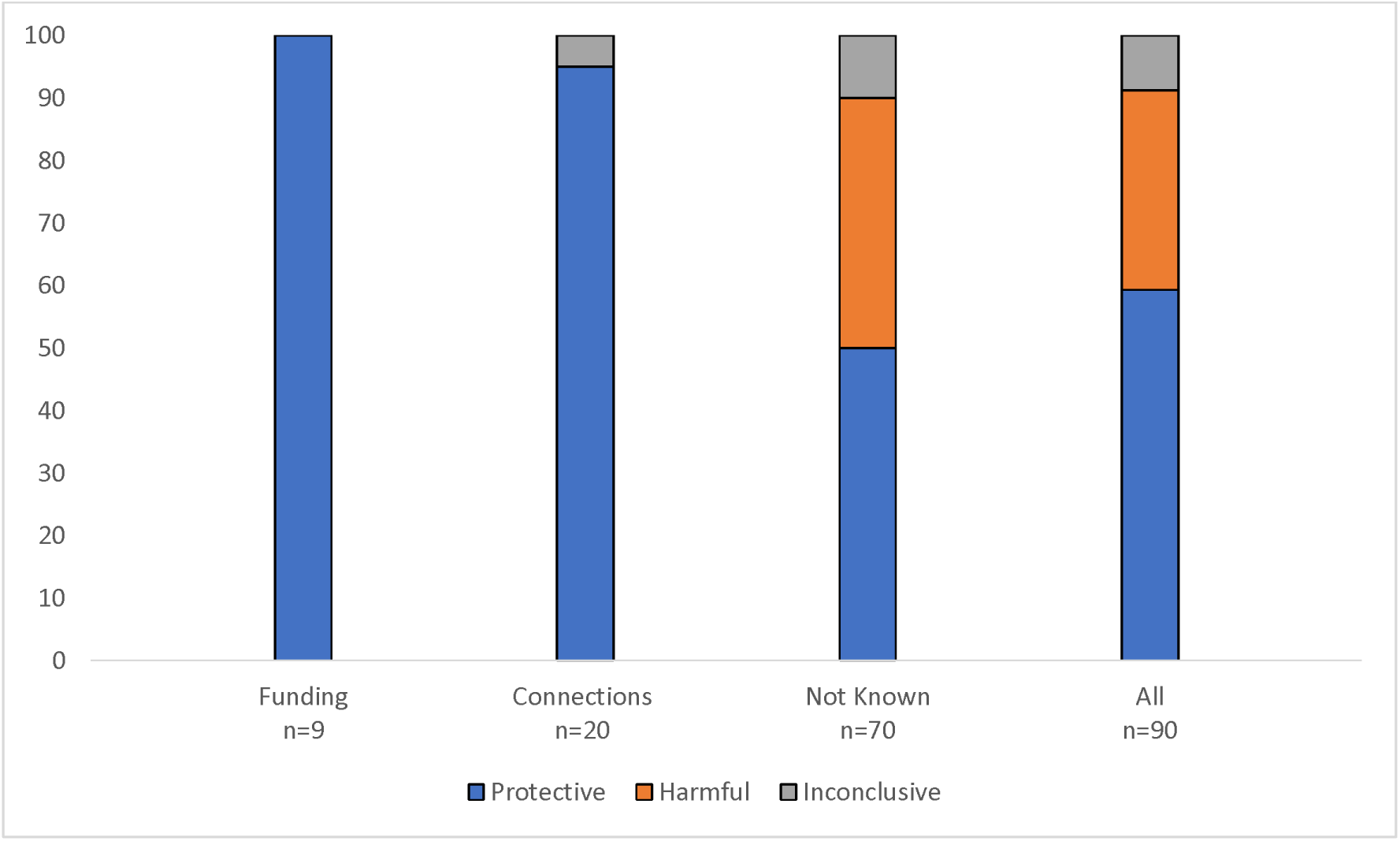
Systematic review conclusions about the association between alcohol consumption and CVD by alcohol industry funding or authors’ industry connections status.

*Funding: systematic reviews with declaration stating directly funded by the alcohol industry. Connections: systematic reviews with funding and/or affiliations with the alcohol industry*.

### Year of publication

The systematic reviews were published between 1996 and 2025, with the greatest number (9 reviews) appearing in 2021. Before 2012, the percentage of systematic reviews with alcohol industry funding or alcohol industry connections was much higher at 46.4% (13/28), compared to 11.3% (7/62) from 2012. The proportion of systematic reviews that had connections with the alcohol industry ranged from 0% to 100% per year (Figure 2).

**Figure 2:**
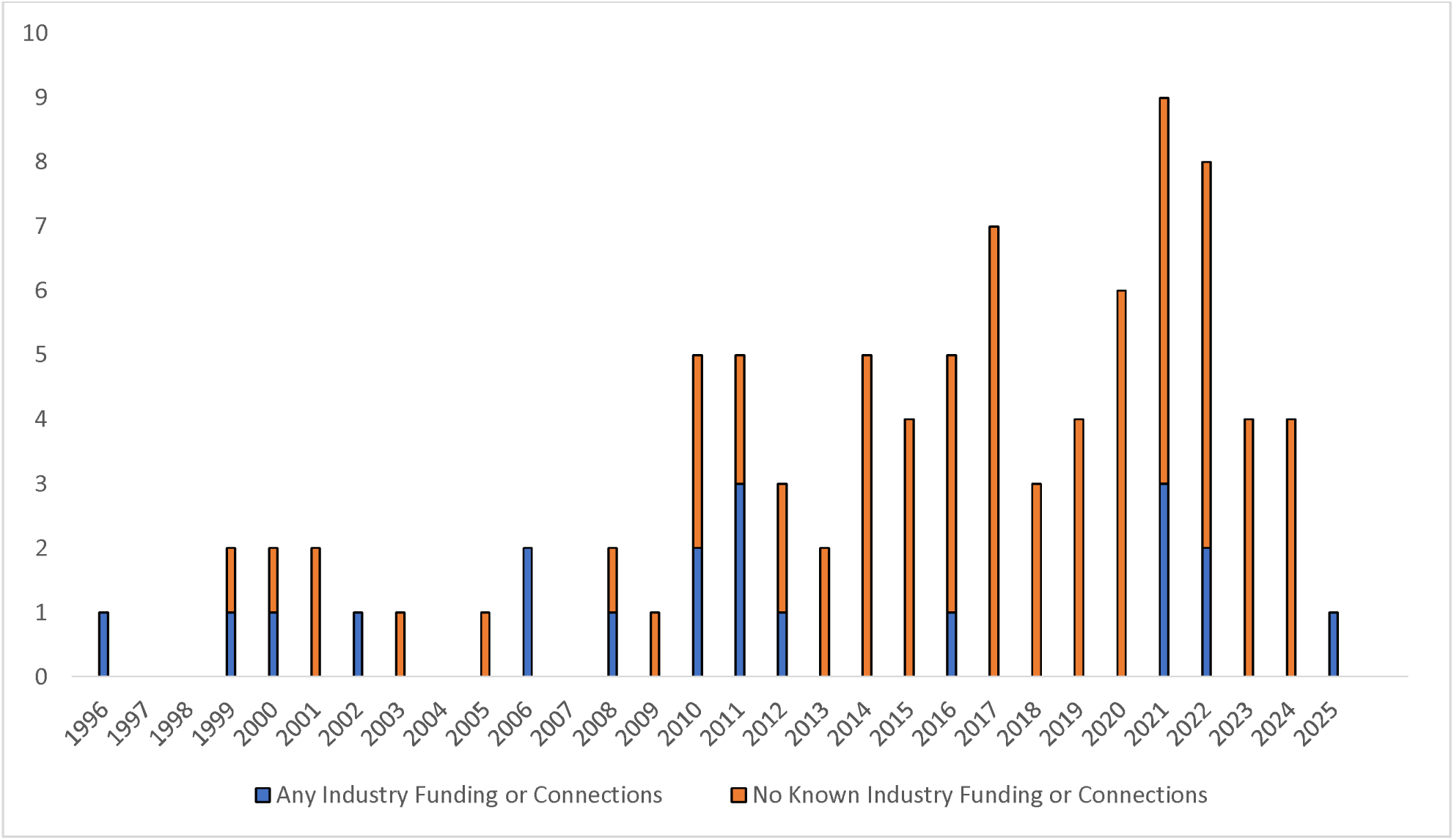
Year of publication by alcohol industry funding or connections status.

### Types of journals

The types of journals in which alcohol industry funded or connected research was published differed slightly from those with no known industry support (Table 1). The most common type of journal for systematic reviews with alcohol industry funding or affiliation was general medicine journals (such as BMJ in three reviews, Archives of Internal Medicine, Journal of the American College of Cardiology, and Circulation) (6/20, 30%), followed by nutrition journals (4/20, 20%) whereas the most common type of journal for systematic reviews without any known alcohol industry funding or connections were cardiovascular journals (20/70, 28.6%) and general medicine journals (16/70, 22.9%).

**Table 1:** Type of journal systematic review published in by funding or affiliation status.

| Type of journal | General medicine | Cardiovascular | Alcohol | Nutrition | Other | Epidemiology |
| --- | --- | --- | --- | --- | --- | --- |
| Industry funded (n=9) | 2/9 (22.2%) | 1/9 (11.1%) | 1/9 (11.1%) | 3/9 (33.3%) | 1/9 (11.1%) | 1/9 (11.1%) |
| Any Industry funding or connection (n=20) | 6/20 (30%) | 3/20 (15%) | 1/20 (5%) | 4/20 (20%) | 3/20 (15%) | 3/20 (15%) |
| No known industry connection (n=70) | 16/70 (22.9%) | 20/70 (28.6%) | 3/70 (4.29%) | 9/70 (12.9%) | 12/70 (17.1%) | 10/70 (14.3%) |

### Types of exposure

The type of alcoholic beverage evaluated within the systematic reviews did not differ by whether alcohol industry funding or connection was present (Table 2). For systematic reviews without any known alcohol industry funding or connection, 87.1% (61/70) evaluated any type of alcoholic beverage compared to 85% (17/20) of reviews with alcohol industry funding or association.

**Table 2:** Type of alcoholic beverage by funding or affiliation status.

| Type of alcoholic beverage | Wine | Beer | Wine and Beer | Spirits | Any |
| --- | --- | --- | --- | --- | --- |
| Industry funded (n=9) | 0/9 (0%) | 1/9 (11.1%) | 0/9 (0%) | 0/9 (0%) | 8/9 (88.9%) |
| Any industry connections (n=20) | 1/20 (5%) | 1/20 (5%) | 1/20 (5%) | 0/20 (0%) | 17/20 (85%) |
| No known industry connection (n=70) | 7/70 (10%) | 1/70 (1.43%) | 0/70 (0%) | 1/70 (1.43%) | 61/70 (87.1%) |

The systematic reviews varied in how they measured alcohol dose intake. Whilst half of the systematic reviews used categorical data with differing thresholds 45/90 (50.0%), others narratively reported the measures in the individual included studies 25/90 (27.8%), whilst others used continuous measures such as grams per day or drinks per day 13/90 (14.4%), some reviews did not report the alcohol dose 4/90 (4.4%) and 3/90 (3.3%) looked at alcohol reduction.

### Outcome measures

The main outcome measures were CVD (43/90, 47.8%), hypertension/blood pressure (15/90, 16.7%), atrial fibrillation (11/90, 12.2%), stroke (8/90, 8.89%), other vascular outcomes (7/90, 7.78%), heart failure (4/90, 4.44%) and biomarkers (4/90, 4.44%). As shown in Table 3, most systematic reviews with alcohol industry funding or connections evaluated general cardiovascular outcomes (17/20) with a further two reviews measuring biomarkers and one heart failure. In contrast, reviews with no known alcohol industry connections also investigated other outcomes including hypertension/blood pressure (15/70), atrial fibrillation (11/70), stroke (8/70) and other vascular outcomes (7/70) with mixed results in addition to cardiovascular outcomes (26/70), biomarkers (2/70) and heart failure (3/70).

**Table 3:** Outcome measures by alcohol industry funding or affiliation status and author conclusions.

| Outcomes measured | Number of SRs with no known alcohol industry connections (n=70) | Number of SRs with alcohol industry connections (n=20) | Number of SRs supportive of protective effect (n=54) | Number of SRs supportive of harmful effect (n=28) | Number of SRs inconclusive or mixed results (n=8) |
| --- | --- | --- | --- | --- | --- |
| CVD (n=43) | 26 (60.5%) | 17 (39.5%) | 33 (77%) | 8 (19%) | 2 (5%) |
| Hypertension/BP (n=15) | 15 (100%) | 0 (0%) | 3 (20%) | 11 (73.33%) | 1 (6.67%) |
| Atrial Fibrillation (n=11) | 11 (100%) | 0 (0%) | 0 (0%) | 9 (82%) | 2 (18%) |
| Biomarker (n=4) | 2 (50%) | 2 (50%) | 4 (100%) | 0 (0%) | 0 (0%) |
| Heart Failure (n=4) | 3 (75%) | 1 (25%) | 4 (100%) | 0 (0%) | 0 (0%) |
| Stroke (n=8) | 8 (100%) | 0 (0%) | 4 (50%) | 1 (13%) | 3 (38%) |
| Other vascular (n=7) | 7 (100%) | 0 (0%) | 4 (57%) | 2 (29%) | 1 (14%) |

BP: Blood pressure; SR: Systematic review

### Number and type of included studies

The number of included studies in each systematic review ranged from 5 to 114, with a mean of 31.7 in the systematic reviews with any alcohol industry funding or connections and a mean of 22.4 in the systematic reviews with no known industry connections.

Of those systematic reviews with any connection with the alcohol industry, 30% (6/20) included experimental studies which is slightly higher than the 27.1% (19/70) of those without any known alcohol industry connections.

### Comparison of included study citations

Of the eight systematic reviews with industry funding declarations within the manuscript, six focused on CVD as the outcome, and two on biomarkers. The six with CVD as the outcome were selected for more detailed analysis of the included primary studies as they could be compared with five systematic reviews with no known alcohol industry funding with the same population, intervention and outcome.

These 11 systematic reviews had a total of 204 included primary studies, of which 163 (80%) were unique to one systematic review, 34 were included in two reviews, six studies were included in three reviews, and one was included in four reviews (Supplementary Table S3: Matrix of included studies in 11 systematic reviews). When we compared the overlap of these included studies using the CCA we obtained a percentage of 2.59% ((204-163)/(11*163-163)). This is considered low overlap (Hennessy & Johnson, 2020). The percentage of included studies that were unique in the systematic reviews with declared alcohol industry funding was higher (mean 82%, range 61% to 96%) than that for the systematic reviews with no known alcohol funding (mean 45%, range 29% to 53%) (Table 4).

**Table 4:**
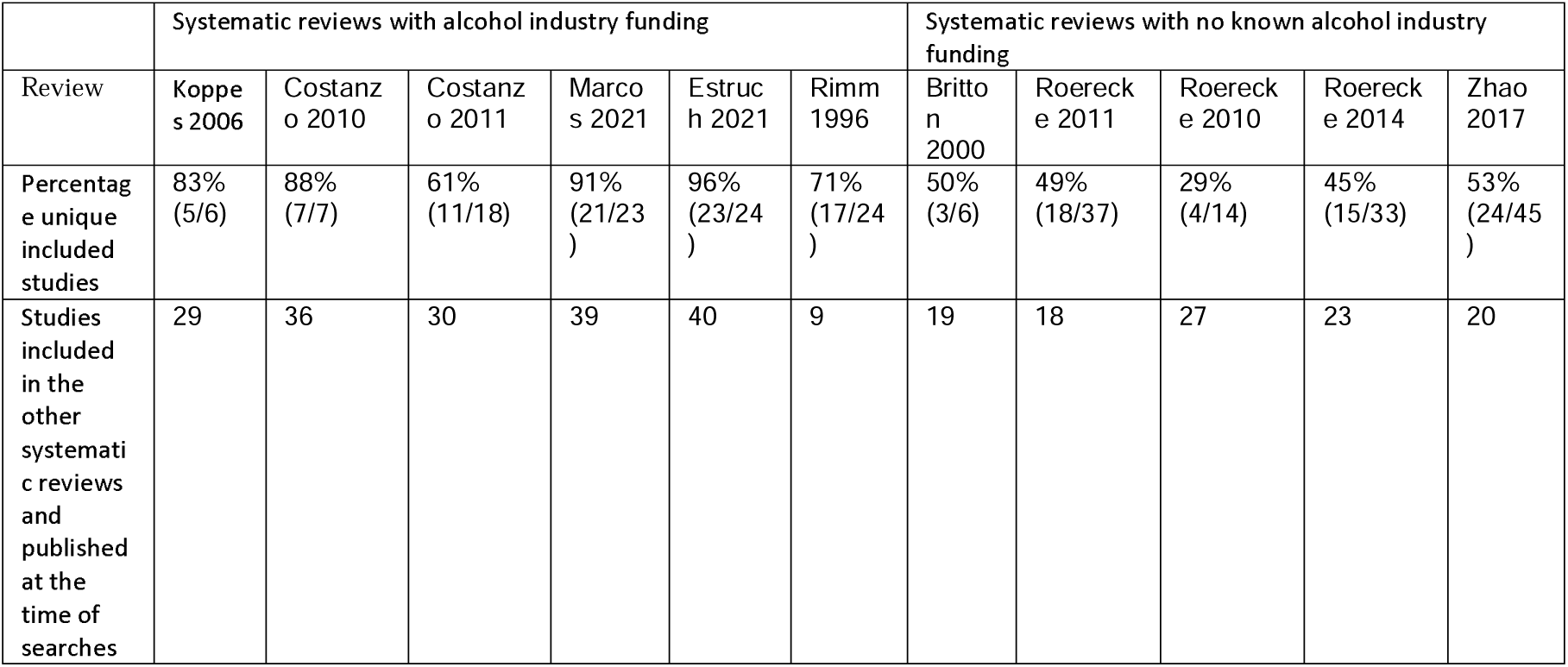
Percentage of unique included studies in 11 systematic reviews.

|  | Systematic reviews with alcohol industry funding |  |  |  |  |  | Systematic reviews with no known alcohol industry funding |  |  |  |  |
| --- | --- | --- | --- | --- | --- | --- | --- | --- | --- | --- | --- |
| Review | Koppe<br>s 2006 | Costanz<br>o 2010 | Costanz<br>o 2011 | Marco<br>s 2021 | Estruc<br>h 2021 | Rimm<br>1996 | Britto<br>n<br>2000 | Roereck<br>e 2011 | Roereck<br>e 2010 | Roereck<br>e 2014 | Zhao<br>2017 |
| Percentag<br>e unique<br>included<br>studies | 83%<br>(5/6) | 88%<br>(7/7) | 61%<br>(11/18) | 91%<br>(21/23<br>) | 96%<br>(23/24<br>) | 71%<br>(17/24<br>) | 50%<br>(3/6) | 49%<br>(18/37) | 29%<br>(4/14) | 45%<br>(15/33) | 53%<br>(24/45<br>) |
| Studies<br>included<br>in the<br>other<br>systemati<br>c reviews<br>and<br>published<br>at the<br>time of<br>searches | 29 | 36 | 30 | 39 | 40 | 9 | 19 | 18 | 27 | 23 | 20 |

### Methodological Quality

Methodological quality was assessed using AMSTAR 2 (Supplementary Table S4). Only one review which had no known alcohol industry connections was rated as high quality – this was a Cochrane review on hypertension (Tasnim 2020). All the other reviews were rated as critically low quality (89/90, 98.9%). Even if we consider a ‘partial yes’ equivalent to a ‘yes’, only the Cochrane review would a high rating, however, five reviews would now have a low rating as opposed to a critically low rating(Brien et al., 2011; Ronksley et al., 2011; Spencer et al., 2017; Stockwell et al., 2016; Zhao et al., 2017).

This meant that we are unable to consider the impact of funding status on quality assessment ratings. The main reasons for obtaining a ‘critical low’ rating included a lack of a review protocol and a lack of a list of excluded studies with reasons.

### Analysis within the systematic reviews

Most of the systematic reviews undertook some form of meta-analysis (68.9%, 62/90). Of those with alcohol industry funding or connections 60% (12/20) provided a meta-analysis compared to 71.4% (50/70) of those with no known industry connections.

Among reviews with alcohol industry funding or connections, 6/20 (30%) took account of the impact of sick quitters on the results compared to 20/70 (28.6%) of reviews without industry funding or connections (Table 5).

**Table 5:** Citations, included studies, analysis and conclusion of systematic reviews.

|  | Mean Times cited | Mean number of included studies | Considered sick quitters in analysis | Undertook subgroup analysis | Included experimental study designs | Author's conclusion states protective effect | Author's conclusion states harmful effect | Author's conclusion mixed or inconclusive |
| --- | --- | --- | --- | --- | --- | --- | --- | --- |
| Industry funded (n=9) | 460.8 | 31.7 | 33.8% (3/9) | 55.6% (5/9) | 55.6% (5/9) | 100% (9/9) | 0% (0/9) | 0% (0/9) |
| Any Industry funding or connections (n=20) | 575.85 | 31.7 | 30% (6/20) | 60% (12/20) | 30% (6/20) | 95% (19/20) | 0% (0/20) | 5% (1/20) |
| No known industry connections (n=70) | 193.0 | 22.4 | 28.6% (20/70) | 71.4% (50/70) | 27.1% (19/70) | 50% (35/70) | 40% (28/70) | 10% (7/70) |
| All (n=90) | 278.1 | 24.5 | 32.2% (29/90) | 68.9% (62/90) | 27.8% (25/90) | 60% (54/90) | 31.1% (28/90) | 8.89% (8/90) |

Of those with industry funding or connections, 12/20 (60%) undertook some form of subgroup analysis whereas 50/70 (71.4%) of those without known industry connections undertook this. Subgroup analysis tended to focus on sex (37/50), age (13/50) and geographical region (17/50).

### Citations to reviews

Reviews with alcohol industry funding or connections had a mean of 575.85 citations (highest 2231), while systematic reviews with no known industry funding or connection had a mean of 193.0 citations (range 0 to 1411). An unpaired t test was performed to compare the number of citations between the two groups (P value = 0.0002). The number of citations per year for industry-supported or associated reviews was 32.0, compared to 17.1 per year for those with no known funding or association (P value = 0.0071). Thus, the difference for citation comparisons is statistically significant even after accounting for earlier or later year of publication.

## Discussion

We evaluated 90 systematic reviews examining the effects of alcohol consumption on CVD. Whilst we only found 20 of these reviews to have alcohol industry connections (funding, history of funding or affiliation), we hypothesize that the true number of reviews with any connection to the alcohol industry is likely to be higher as some industry affiliations will probably not have been visible in the sources examined, leading to Type II error in any industry/non-industry statistical comparison. In addition, many of these systematic reviews are likely to incorporate primary studies that themselves received alcohol industry funding or support, which was beyond the scope of this work to assess. As a result, comparisons between reviews with reported industry connections and those without is likely underestimating the true influence of industry funding and should be regarded as a minimum estimate.

Nevertheless, we found some important differences between industry and non-industry funded reviews. Systematic reviews with observed alcohol industry connections were more likely to identify a protective effect of low to moderate alcohol consumption on CVD, evaluate broader outcomes, include experimental studies, be published in nutrition or general medicine journals and have a higher number of citations. On the other hand, systematic reviews without any known alcohol industry connections were more likely to have narrower outcomes (e.g. stroke, hypertension), be published in cardiovascular journals, conduct subgroup analysis, and have a lower number of citations. Until 2011, most systematic reviews were linked with the alcohol industry. Given that these suggest the presence of a protective effect for alcohol on CVD, it might explain why the ‘J-curve’ has been promoted for decades and continues to be.

The variation in the definitions employed in the reviews for defining alcohol consumption, complicates any comparisons between the reviews. The different categories and definitions of ‘low’ and ‘moderate’ drinking may partially reflect differences over time as well as differences internationally (Litten & Gardner, 2022).

The quality assessment ratings using AMSTAR 2 were unable to illuminate whether there were any quality patterns by alcohol industry connection status as 99% of the systematic reviews were rated as critically low. Only one review (a Cochrane Review(Tasnim et al., 2020) – not industry funded) was rated as high. Having them all rated the same is not helpful to distinguish differences in methodological quality. It should be noted that AMSTAR 2 often rates a high proportion of reviews as low or critically low quality (De Santis et al., 2023).

It is worth noting that the high-quality rated review only included RCTs which compared the effects of alcohol versus placebo on blood pressure on a very short-time frame (a few hours)(Tasnim et al., 2020). While it is well executed, it should be noted that the acute effects of alcohol consumption are understood to be very different from the effects of chronic exposure. Arguably, given that asking participants to drink a certain amount of alcohol during several years is impractical and especially unethical, observational studies such as cohorts are better positioned to evaluate the topic despite being less robust due to confounding and underreporting of alcohol consumption(Stockwell, Zhao, & Macdonald, 2014).

Our original study of 60 systematic reviews highlighted that despite a large number of systematic reviews in this area controversy remained on the cardioprotective effect of alcohol. Five years on and the basic findings on industry bias have been replicated with a further 30 systematic reviews. This is line with other research which indicates that alcohol’s purported health benefits continue to be promoted by industry(Clay et al., 2025).

The high number of poor-quality rated systematic reviews in this area and limited study overlap between studies is disappointing. In addition to wasting funds, poor quality reviews add confusion in the evidence base and make it challenging to identify the most robust and reliable reviews. As far back as 1994, Altman infamously stated that ‘we need less research, better research and research done for the right reasons’(Altman, 1994). Fewer high quality systematic reviews with comprehensive search strategies may be preferable to 90 poor quality systematic reviews.

We believe that while other, more subtle differences may exist in addition to those we identified, our findings may be generalisable to other topic areas. Meta research on systematic reviews of the harms on gabapentin uncovered different results for systematic reviews even when the same data sources are used (Qureshi et al., 2022). These differences were attributed to different selections of harms and different analyses. This can be compared to the different cardiovascular outcomes selected in our cohort of systematic reviews. Moreover, research on primary studies indicates that misclassification bias extends much further than sick quitters (Naimi et al., 2017). A more in-depth analysis may be able to ascertain the true nuances of the review methodologies that lead to opposing conclusions.

## Limitations

Despite checking the self-declarations within the manuscripts (within the funding and conflict of interest statements) and checking individual authors on the internet, web of science core collection databases and a social media platform (LinkedIn) we are likely to have missed many connections with the alcohol industry including funding, other support, or affiliations given the extensive efforts industries make in order to obfuscate their influence on science and scientists.

Due to the large volume of systematic reviews, we were unable to perform a comprehensive analysis of primary study overlap, quality, and funding of the included primary studies. We did, however, undertake an analysis on a small sample of reviews which indicated little overlap between the primary studies included in systematic reviews with and without known alcohol industry funding. Further research could be undertaken to compare the included primary studies in more detail.

We were also unable to examine the prevalence of spin (reporting practices that distort the interpretation of research results, making the findings appear more favourable than the data actually support) within the systematic reviews and any discordance between results and conclusions due to logistical constraints. Previous studies have indicated that spin in the abstracts may not be more pronounced in industry funded articles; however, discordance has been observed in primary studies (Qureshi et al., 2024).

## Conclusions

This meta-review highlights that systematic reviews with alcohol industry connections consistently favoured cardioprotective interpretations of low or moderate alcohol consumption, were more widely cited, and often differed in study selection and outcome selection compared to reviews with no known alcohol industry connections. All reviews but one were critically low in quality and - using a sample of reviews - there was minimal overlap in the included studies. Overall, this is a complex field of study that exhibits sponsorship and needs more rigorous analyses, both in primary studies and syntheses.

## Data Availability

All data produced in the present work are contained in the manuscript or publicly available.

## Supplementary Material

**Supplementary Table S1:**
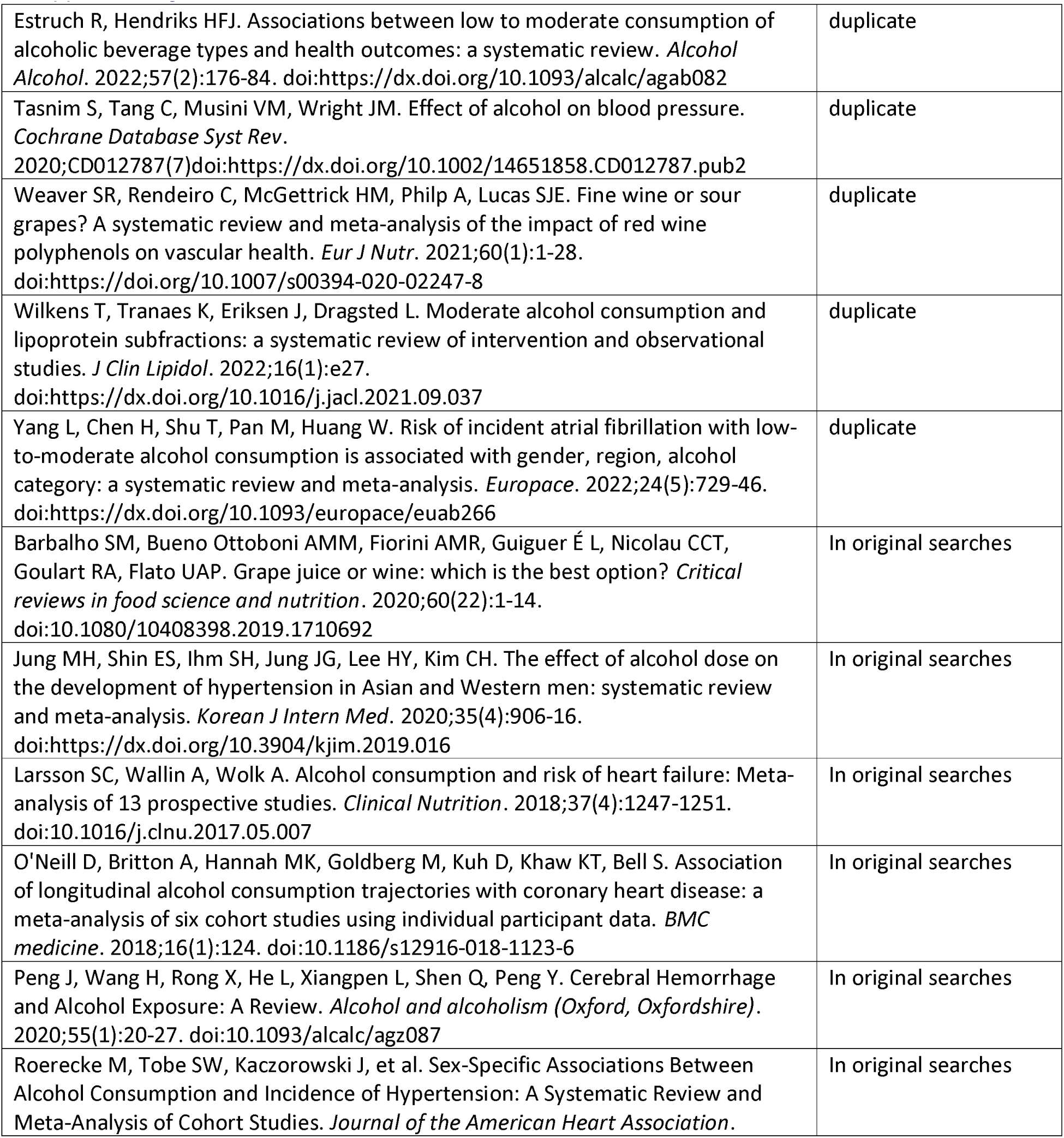
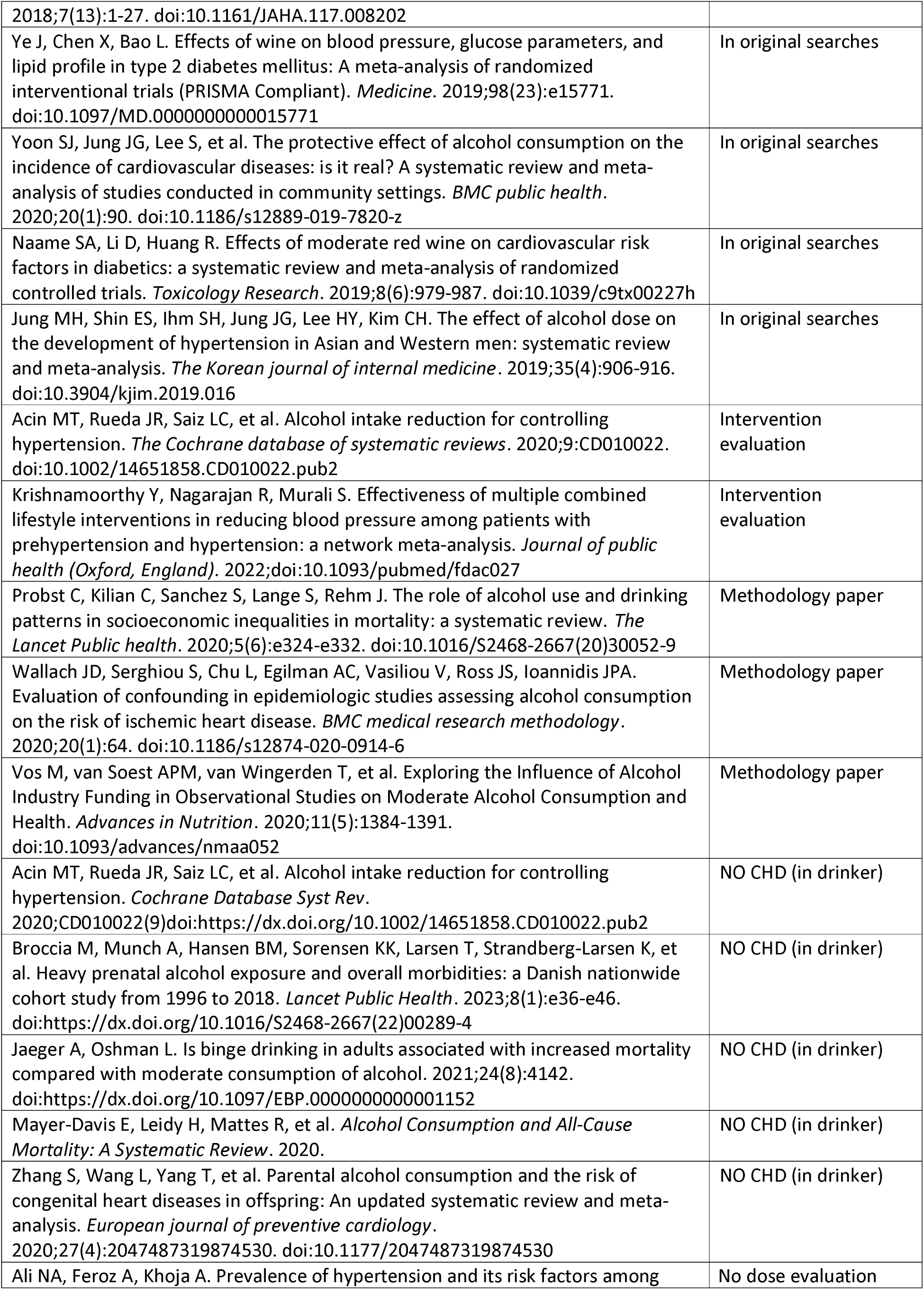
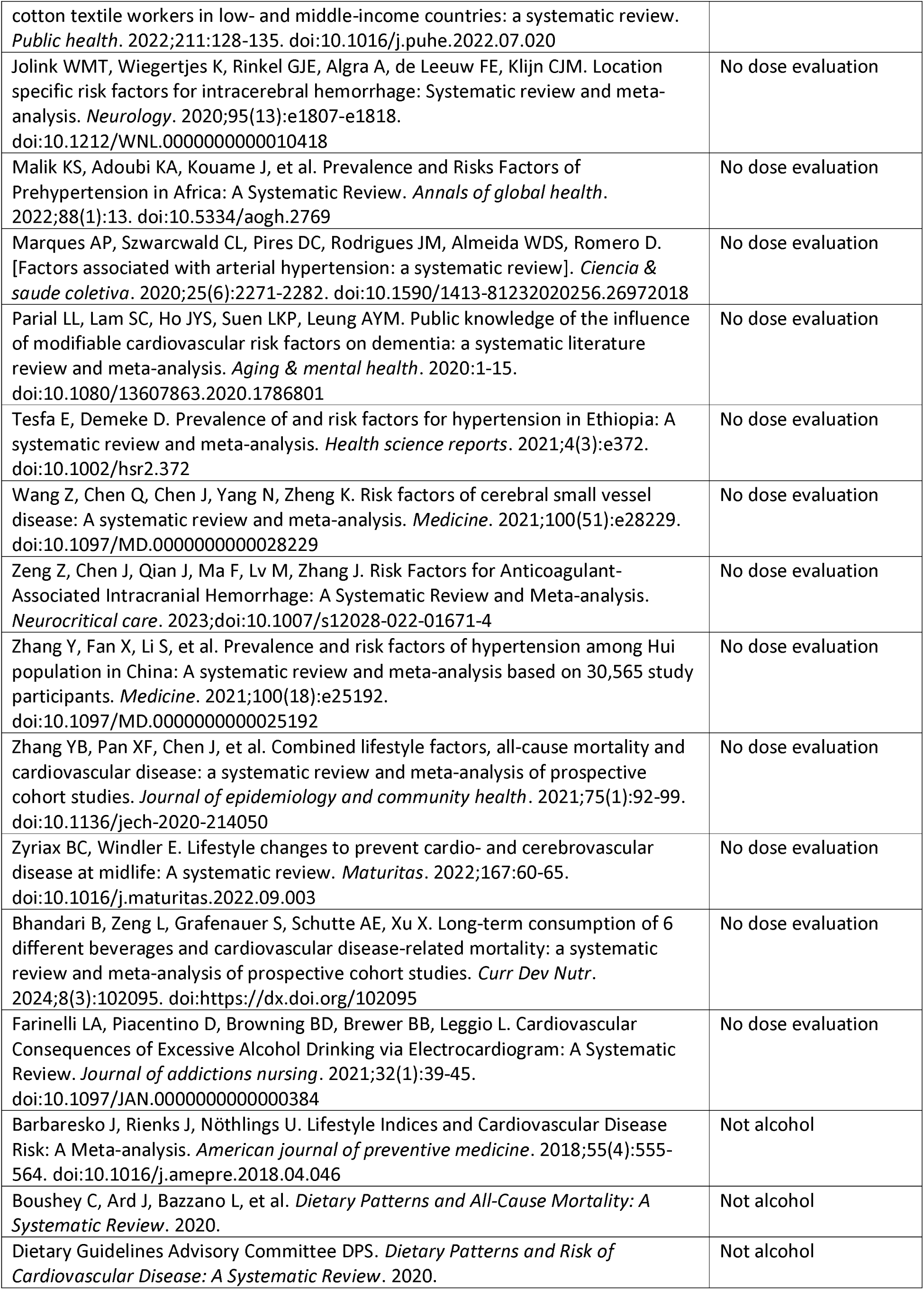
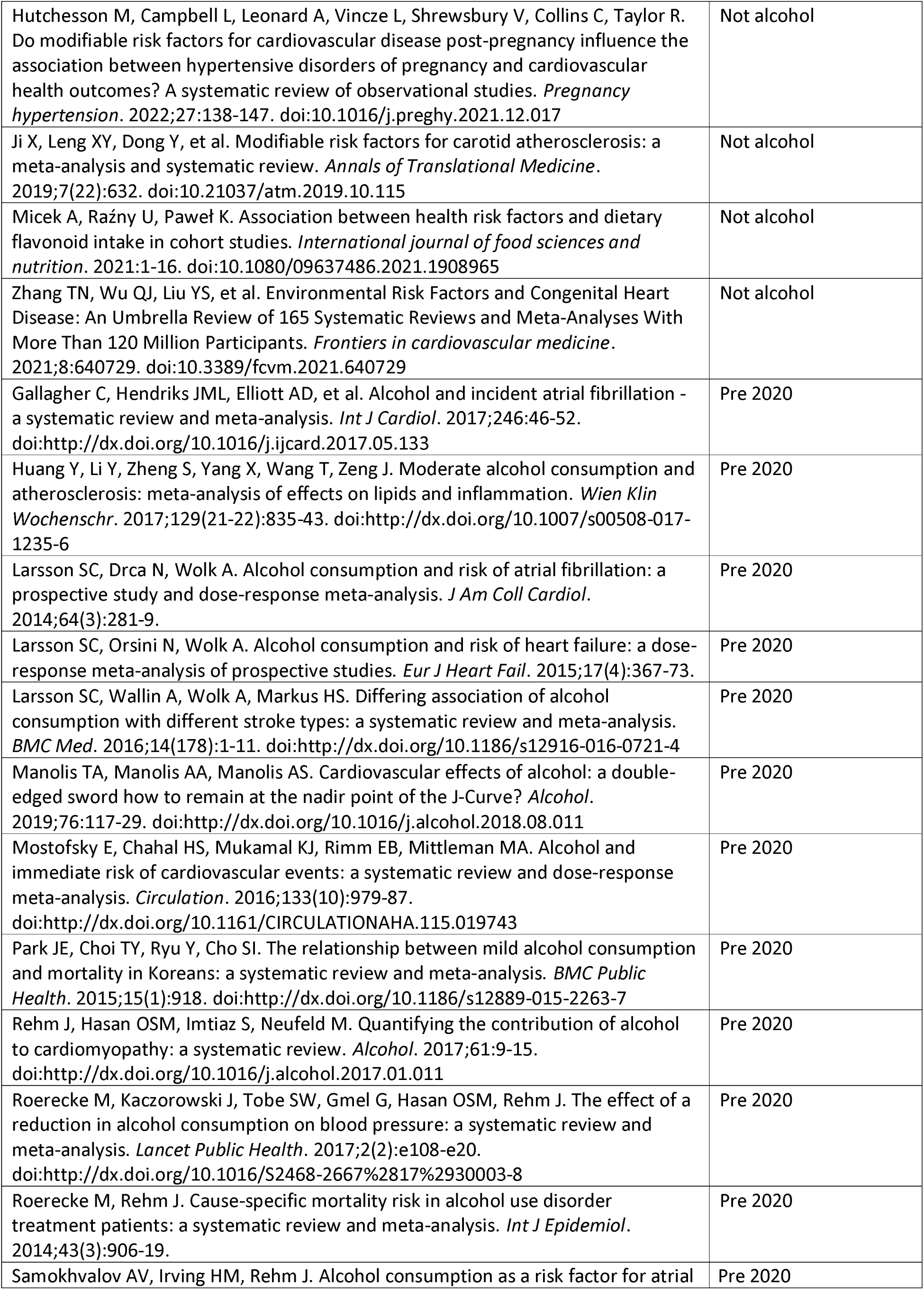
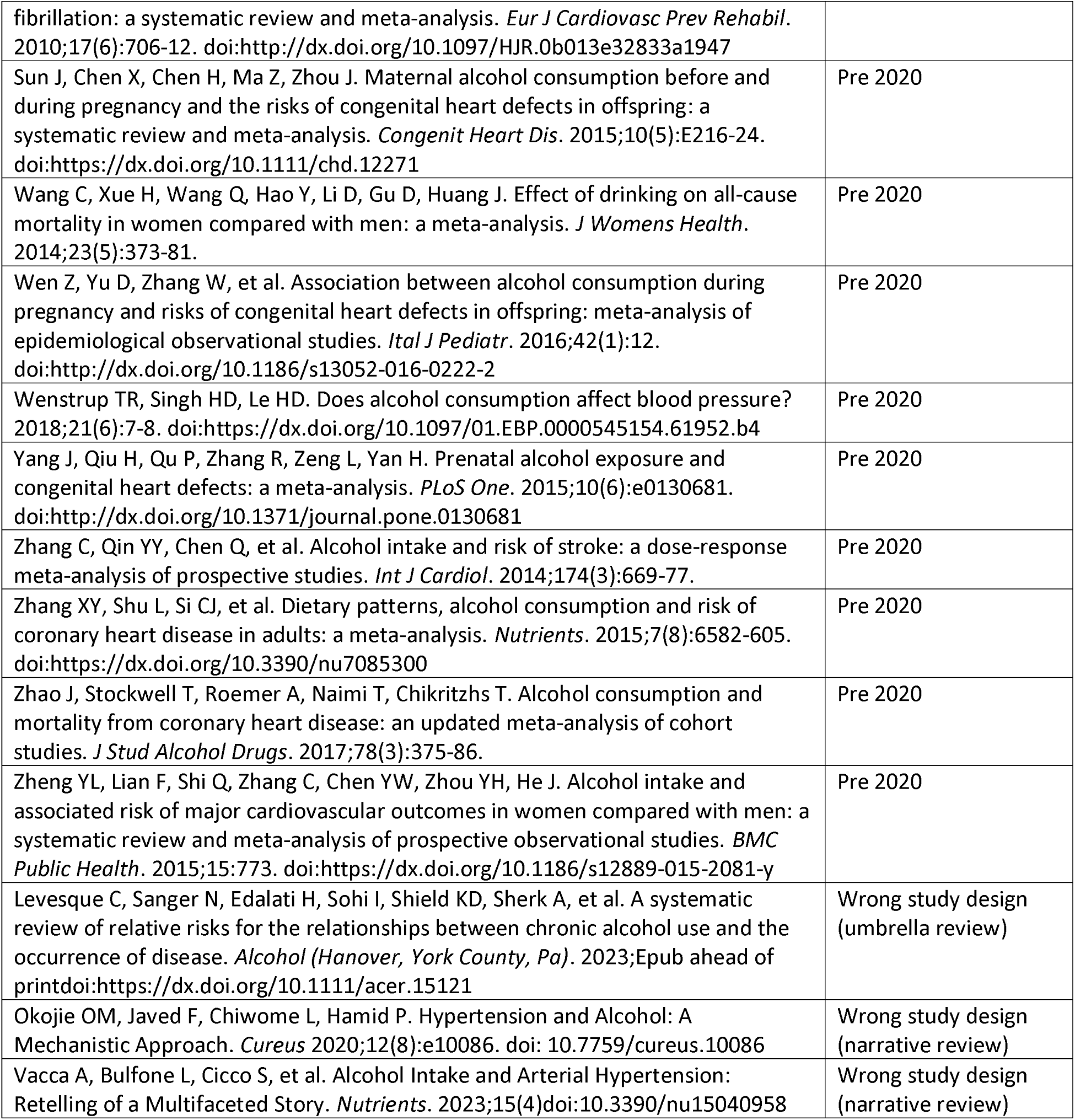
Excluded Studies and reasons for exclusion.

|  |  |
| --- | --- |
| Estruch R, Hendriks HFJ. Associations between low to moderate consumption of alcoholic beverage types and health outcomes: a systematic review. *Alcohol Alcohol*. 2022;57(2):176-84. doi:https://dx.doi.org/10.1093/alcalc/agab082 | duplicate |
| Tasnim S, Tang C, Musini VM, Wright JM. Effect of alcohol on blood pressure. *Cochrane Database Syst Rev*. 2020;CD012787(7)doi:https://dx.doi.org/10.1002/14651858.CD012787.pub2 | duplicate |
| Weaver SR, Rendeiro C, McGettrick HM, Philp A, Lucas SJE. Fine wine or sour grapes? A systematic review and meta-analysis of the impact of red wine polyphenols on vascular health. *Eur J Nutr*. 2021;60(1):1-28. doi:https://doi.org/10.1007/s00394-020-02247-8 | duplicate |
| Wilkens T, Tranaes K, Eriksen J, Dragsted L. Moderate alcohol consumption and lipoprotein subfractions: a systematic review of intervention and observational studies. *J Clin Lipidol*. 2022;16(1):e27. doi:https://dx.doi.org/10.1016/j.jacl.2021.09.037 | duplicate |
| Yang L, Chen H, Shu T, Pan M, Huang W. Risk of incident atrial fibrillation with low-to-moderate alcohol consumption is associated with gender, region, alcohol category: a systematic review and meta-analysis. *Europace*. 2022;24(5):729-46. doi:https://dx.doi.org/10.1093/europace/euab266 | duplicate |
| Barbalho SM, Bueno Ottoboni AMM, Fiorini AMR, Guiguer É L, Nicolau CCT, Goulart RA, Flato UAP. Grape juice or wine: which is the best option? *Critical reviews in food science and nutrition*. 2020;60(22):1-14. doi:10.1080/10408398.2019.1710692 | In original searches |
| Jung MH, Shin ES, Ihm SH, Jung JG, Lee HY, Kim CH. The effect of alcohol dose on the development of hypertension in Asian and Western men: systematic review and meta-analysis. *Korean J Intern Med*. 2020;35(4):906-16. doi:https://dx.doi.org/10.3904/kjim.2019.016 | In original searches |
| Larsson SC, Wallin A, Wolk A. Alcohol consumption and risk of heart failure: Meta-analysis of 13 prospective studies. *Clinical Nutrition*. 2018;37(4):1247-1251. doi:10.1016/j.clnu.2017.05.007 | In original searches |
| O'Neill D, Britton A, Hannah MK, Goldberg M, Kuh D, Khaw KT, Bell S. Association of longitudinal alcohol consumption trajectories with coronary heart disease: a meta-analysis of six cohort studies using individual participant data. *BMC medicine*. 2018;16(1):124. doi:10.1186/s12916-018-1123-6 | In original searches |
| Peng J, Wang H, Rong X, He L, Xiangpen L, Shen Q, Peng Y. Cerebral Hemorrhage and Alcohol Exposure: A Review. *Alcohol and alcoholism (Oxford, Oxfordshire)*. 2020;55(1):20-27. doi:10.1093/alcalc/agz087 | In original searches |
| Roerecke M, Tobe SW, Kaczorowski J, et al. Sex-Specific Associations Between Alcohol Consumption and Incidence of Hypertension: A Systematic Review and Meta-Analysis of Cohort Studies. *Journal of the American Heart Association*. | In original searches |
| 2018;7(13):1-27. doi:10.1161/JAHA.117.008202 |  |
| Ye J, Chen X, Bao L. Effects of wine on blood pressure, glucose parameters, and lipid profile in type 2 diabetes mellitus: A meta-analysis of randomized interventional trials (PRISMA Compliant). *Medicine*. 2019;98(23):e15771. doi:10.1097/MD.00000000000015771 | In original searches |
| Yoon SJ, Jung JG, Lee S, et al. The protective effect of alcohol consumption on the incidence of cardiovascular diseases: is it real? A systematic review and meta-analysis of studies conducted in community settings. *BMC public health*. 2020;20(1):90. doi:10.1186/s12889-019-7820-z | In original searches |
| Naame SA, Li D, Huang R. Effects of moderate red wine on cardiovascular risk factors in diabetics: a systematic review and meta-analysis of randomized controlled trials. *Toxicology Research*. 2019;8(6):979-987. doi:10.1039/c9tx00227h | In original searches |
| Jung MH, Shin ES, Ihm SH, Jung JG, Lee HY, Kim CH. The effect of alcohol dose on the development of hypertension in Asian and Western men: systematic review and meta-analysis. *The Korean journal of internal medicine*. 2019;35(4):906-916. doi:10.3904/kjim.2019.016 | In original searches |
| Acin MT, Rueda JR, Saiz LC, et al. Alcohol intake reduction for controlling hypertension. *The Cochrane database of systematic reviews*. 2020;9:CD010022. doi:10.1002/14651858.CD010022.pub2 | Intervention evaluation |
| Krishnamoorthy Y, Nagarajan R, Murali S. Effectiveness of multiple combined lifestyle interventions in reducing blood pressure among patients with prehypertension and hypertension: a network meta-analysis. *Journal of public health (Oxford, England)*. 2022;doi:10.1093/pubmed/fdac027 | Intervention evaluation |
| Probst C, Kilian C, Sanchez S, Lange S, Rehm J. The role of alcohol use and drinking patterns in socioeconomic inequalities in mortality: a systematic review. *The Lancet Public health*. 2020;5(6):e324-e332. doi:10.1016/S2468-2667(20)30052-9 | Methodology paper |
| Wallach JD, Serghiou S, Chu L, Egilman AC, Vasiliou V, Ross JS, Ioannidis JPA. Evaluation of confounding in epidemiologic studies assessing alcohol consumption on the risk of ischemic heart disease. *BMC medical research methodology*. 2020;20(1):64. doi:10.1186/s12874-020-0914-6 | Methodology paper |
| Vos M, van Soest APM, van Wingerden T, et al. Exploring the Influence of Alcohol Industry Funding in Observational Studies on Moderate Alcohol Consumption and Health. *Advances in Nutrition*. 2020;11(5):1384-1391. doi:10.1093/advances/nmaa052 | Methodology paper |
| Acin MT, Rueda JR, Saiz LC, et al. Alcohol intake reduction for controlling hypertension. *Cochrane Database Syst Rev*. 2020;CD010022(9)doi:https://dx.doi.org/10.1002/14651858.CD010022.pub2 | NO CHD (in drinker) |
| Broccia M, Munch A, Hansen BM, Sorensen KK, Larsen T, Strandberg-Larsen K, et al. Heavy prenatal alcohol exposure and overall morbidities: a Danish nationwide cohort study from 1996 to 2018. *Lancet Public Health*. 2023;8(1):e36-e46. doi:https://dx.doi.org/10.1016/S2468-2667(22)00289-4 | NO CHD (in drinker) |
| Jaeger A, Oshman L. Is binge drinking in adults associated with increased mortality compared with moderate consumption of alcohol. 2021;24(8):4142. doi:https://dx.doi.org/10.1097/EBP.0000000000001152 | NO CHD (in drinker) |
| Mayer-Davis E, Leidy H, Mattes R, et al. *Alcohol Consumption and All-Cause Mortality: A Systematic Review*. 2020. | NO CHD (in drinker) |
| Zhang S, Wang L, Yang T, et al. Parental alcohol consumption and the risk of congenital heart diseases in offspring: An updated systematic review and meta-analysis. *European journal of preventive cardiology*. 2020;27(4):2047487319874530. doi:10.1177/2047487319874530 | NO CHD (in drinker) |
| Ali NA, Feroz A, Khoja A. Prevalence of hypertension and its risk factors among | No dose evaluation |
| cotton textile workers in low- and middle-income countries: a systematic review. *Public health*. 2022;211:128-135. doi:10.1016/j.puhe.2022.07.020 |  |
| Jolink WMT, Wiegertjes K, Rinkel GJE, Algra A, de Leeuw FE, Klijn CJM. Location specific risk factors for intracerebral hemorrhage: Systematic review and meta-analysis. *Neurology*. 2020;95(13):e1807-e1818. doi:10.1212/WNL.00000000000010418 | No dose evaluation |
| Malik KS, Adoubi KA, Kouame J, et al. Prevalence and Risks Factors of Prehypertension in Africa: A Systematic Review. *Annals of global health*. 2022;88(1):13. doi:10.5334/aogh.2769 | No dose evaluation |
| Marques AP, Szwarcwald CL, Pires DC, Rodrigues JM, Almeida WDS, Romero D. [Factors associated with arterial hypertension: a systematic review]. *Ciencia & saude coletiva*. 2020;25(6):2271-2282. doi:10.1590/1413-81232020256.26972018 | No dose evaluation |
| Parial LL, Lam SC, Ho JYS, Suen LKP, Leung AYM. Public knowledge of the influence of modifiable cardiovascular risk factors on dementia: a systematic literature review and meta-analysis. *Aging & mental health*. 2020:1-15. doi:10.1080/13607863.2020.1786801 | No dose evaluation |
| Tesfa E, Demeke D. Prevalence of and risk factors for hypertension in Ethiopia: A systematic review and meta-analysis. *Health science reports*. 2021;4(3):e372. doi:10.1002/hsr2.372 | No dose evaluation |
| Wang Z, Chen Q, Chen J, Yang N, Zheng K. Risk factors of cerebral small vessel disease: A systematic review and meta-analysis. *Medicine*. 2021;100(51):e28229. doi:10.1097/MD.00000000000028229 | No dose evaluation |
| Zeng Z, Chen J, Qian J, Ma F, Lv M, Zhang J. Risk Factors for Anticoagulant-Associated Intracranial Hemorrhage: A Systematic Review and Meta-analysis. *Neurocritical care*. 2023;doi:10.1007/s12028-022-01671-4 | No dose evaluation |
| Zhang Y, Fan X, Li S, et al. Prevalence and risk factors of hypertension among Hui population in China: A systematic review and meta-analysis based on 30,565 study participants. *Medicine*. 2021;100(18):e25192. doi:10.1097/MD.00000000000025192 | No dose evaluation |
| Zhang YB, Pan XF, Chen J, et al. Combined lifestyle factors, all-cause mortality and cardiovascular disease: a systematic review and meta-analysis of prospective cohort studies. *Journal of epidemiology and community health*. 2021;75(1):92-99. doi:10.1136/jech-2020-214050 | No dose evaluation |
| Zyriax BC, Windler E. Lifestyle changes to prevent cardio- and cerebrovascular disease at midlife: A systematic review. *Maturitas*. 2022;167:60-65. doi:10.1016/j.maturitas.2022.09.003 | No dose evaluation |
| Bhandari B, Zeng L, Grafenauer S, Schutte AE, Xu X. Long-term consumption of 6 different beverages and cardiovascular disease-related mortality: a systematic review and meta-analysis of prospective cohort studies. *Curr Dev Nutr*. 2024;8(3):102095. doi:https://dx.doi.org/102095 | No dose evaluation |
| Farinelli LA, Piacentino D, Browning BD, Brewer BB, Leggio L. Cardiovascular Consequences of Excessive Alcohol Drinking via Electrocardiogram: A Systematic Review. *Journal of addictions nursing*. 2021;32(1):39-45. doi:10.1097/JAN.0000000000000384 | No dose evaluation |
| Barbaresko J, Rienks J, Nöthlings U. Lifestyle Indices and Cardiovascular Disease Risk: A Meta-analysis. *American journal of preventive medicine*. 2018;55(4):555-564. doi:10.1016/j.amepre.2018.04.046 | Not alcohol |
| Boushey C, Ard J, Bazzano L, et al. *Dietary Patterns and All-Cause Mortality: A Systematic Review*. 2020. | Not alcohol |
| Dietary Guidelines Advisory Committee DPS. *Dietary Patterns and Risk of Cardiovascular Disease: A Systematic Review*. 2020. | Not alcohol |
| Hutchesson M, Campbell L, Leonard A, Vincze L, Shrewsbury V, Collins C, Taylor R. Do modifiable risk factors for cardiovascular disease post-pregnancy influence the association between hypertensive disorders of pregnancy and cardiovascular health outcomes? A systematic review of observational studies. *Pregnancy hypertension*. 2022;27:138-147. doi:10.1016/j.preghy.2021.12.017 | Not alcohol |
| Ji X, Leng XY, Dong Y, et al. Modifiable risk factors for carotid atherosclerosis: a meta-analysis and systematic review. *Annals of Translational Medicine*. 2019;7(22):632. doi:10.21037/atm.2019.10.115 | Not alcohol |
| Micek A, Rażny U, Paweł K. Association between health risk factors and dietary flavonoid intake in cohort studies. *International journal of food sciences and nutrition*. 2021;1-16. doi:10.1080/09637486.2021.1908965 | Not alcohol |
| Zhang TN, Wu QJ, Liu YS, et al. Environmental Risk Factors and Congenital Heart Disease: An Umbrella Review of 165 Systematic Reviews and Meta-Analyses With More Than 120 Million Participants. *Frontiers in cardiovascular medicine*. 2021;8:640729. doi:10.3389/fcvm.2021.640729 | Not alcohol |
| Gallagher C, Hendriks JML, Elliott AD, et al. Alcohol and incident atrial fibrillation - a systematic review and meta-analysis. *Int J Cardiol*. 2017;246:46-52. doi:http://dx.doi.org/10.1016/j.ijcard.2017.05.133 | Pre 2020 |
| Huang Y, Li Y, Zheng S, Yang X, Wang T, Zeng J. Moderate alcohol consumption and atherosclerosis: meta-analysis of effects on lipids and inflammation. *Wien Klin Wochenschr*. 2017;129(21-22):835-43. doi:http://dx.doi.org/10.1007/s00508-017-1235-6 | Pre 2020 |
| Larsson SC, Drca N, Wolk A. Alcohol consumption and risk of atrial fibrillation: a prospective study and dose-response meta-analysis. *J Am Coll Cardiol*. 2014;64(3):281-9. | Pre 2020 |
| Larsson SC, Orsini N, Wolk A. Alcohol consumption and risk of heart failure: a dose-response meta-analysis of prospective studies. *Eur J Heart Fail*. 2015;17(4):367-73. | Pre 2020 |
| Larsson SC, Wallin A, Wolk A, Markus HS. Differing association of alcohol consumption with different stroke types: a systematic review and meta-analysis. *BMC Med*. 2016;14(178):1-11. doi:http://dx.doi.org/10.1186/s12916-016-0721-4 | Pre 2020 |
| Manolis TA, Manolis AA, Manolis AS. Cardiovascular effects of alcohol: a double-edged sword how to remain at the nadir point of the J-Curve? *Alcohol*. 2019;76:117-29. doi:http://dx.doi.org/10.1016/j.alcohol.2018.08.011 | Pre 2020 |
| Mostofsky E, Chahal HS, Mukamal KJ, Rimm EB, Mittleman MA. Alcohol and immediate risk of cardiovascular events: a systematic review and dose-response meta-analysis. *Circulation*. 2016;133(10):979-87. doi:http://dx.doi.org/10.1161/CIRCULATIONAHA.115.019743 | Pre 2020 |
| Park JE, Choi TY, Ryu Y, Cho SI. The relationship between mild alcohol consumption and mortality in Koreans: a systematic review and meta-analysis. *BMC Public Health*. 2015;15(1):918. doi:http://dx.doi.org/10.1186/s12889-015-2263-7 | Pre 2020 |
| Rehm J, Hasan OSM, Imtiaz S, Neufeld M. Quantifying the contribution of alcohol to cardiomyopathy: a systematic review. *Alcohol*. 2017;61:9-15. doi:http://dx.doi.org/10.1016/j.alcohol.2017.01.011 | Pre 2020 |
| Roerecke M, Kaczorowski J, Tobe SW, Gmel G, Hasan OSM, Rehm J. The effect of a reduction in alcohol consumption on blood pressure: a systematic review and meta-analysis. *Lancet Public Health*. 2017;2(2):e108-e20. doi:http://dx.doi.org/10.1016/S2468-2667%2817%2930003-8 | Pre 2020 |
| Roerecke M, Rehm J. Cause-specific mortality risk in alcohol use disorder treatment patients: a systematic review and meta-analysis. *Int J Epidemiol*. 2014;43(3):906-19. | Pre 2020 |
| Samokhvalov AV, Irving HM, Rehm J. Alcohol consumption as a risk factor for atrial | Pre 2020 |
| fibrillation: a systematic review and meta-analysis. *Eur J Cardiovasc Prev Rehabil*. 2010;17(6):706-12. doi:http://dx.doi.org/10.1097/HJR.0b013e32833a1947 |  |
| Sun J, Chen X, Chen H, Ma Z, Zhou J. Maternal alcohol consumption before and during pregnancy and the risks of congenital heart defects in offspring: a systematic review and meta-analysis. *Congenit Heart Dis*. 2015;10(5):E216-24. doi:https://dx.doi.org/10.1111/chd.12271 | Pre 2020 |
| Wang C, Xue H, Wang Q, Hao Y, Li D, Gu D, Huang J. Effect of drinking on all-cause mortality in women compared with men: a meta-analysis. *J Womens Health*. 2014;23(5):373-81. | Pre 2020 |
| Wen Z, Yu D, Zhang W, et al. Association between alcohol consumption during pregnancy and risks of congenital heart defects in offspring: meta-analysis of epidemiological observational studies. *Ital J Pediatr*. 2016;42(1):12. doi:http://dx.doi.org/10.1186/s13052-016-0222-2 | Pre 2020 |
| Wenstrup TR, Singh HD, Le HD. Does alcohol consumption affect blood pressure? 2018;21(6):7-8. doi:https://dx.doi.org/10.1097/01.EBP.0000545154.61952.b4 | Pre 2020 |
| Yang J, Qiu H, Qu P, Zhang R, Zeng L, Yan H. Prenatal alcohol exposure and congenital heart defects: a meta-analysis. *PLoS One*. 2015;10(6):e0130681. doi:http://dx.doi.org/10.1371/journal.pone.0130681 | Pre 2020 |
| Zhang C, Qin YY, Chen Q, et al. Alcohol intake and risk of stroke: a dose-response meta-analysis of prospective studies. *Int J Cardiol*. 2014;174(3):669-77. | Pre 2020 |
| Zhang XY, Shu L, Si CJ, et al. Dietary patterns, alcohol consumption and risk of coronary heart disease in adults: a meta-analysis. *Nutrients*. 2015;7(8):6582-605. doi:https://dx.doi.org/10.3390/nu7085300 | Pre 2020 |
| Zhao J, Stockwell T, Roemer A, Naimi T, Chikritzhs T. Alcohol consumption and mortality from coronary heart disease: an updated meta-analysis of cohort studies. *J Stud Alcohol Drugs*. 2017;78(3):375-86. | Pre 2020 |
| Zheng YL, Lian F, Shi Q, Zhang C, Chen YW, Zhou YH, He J. Alcohol intake and associated risk of major cardiovascular outcomes in women compared with men: a systematic review and meta-analysis of prospective observational studies. *BMC Public Health*. 2015;15:773. doi:https://dx.doi.org/10.1186/s12889-015-2081-y | Pre 2020 |
| Levesque C, Sanger N, Edalati H, Sohi I, Shield KD, Sherk A, et al. A systematic review of relative risks for the relationships between chronic alcohol use and the occurrence of disease. *Alcohol (Hanover, York County, Pa)*. 2023;Epub ahead of printdoi:https://dx.doi.org/10.1111/acer.15121 | Wrong study design (umbrella review) |
| Okojie OM, Javed F, Chiwome L, Hamid P. Hypertension and Alcohol: A Mechanistic Approach. *Cureus* 2020;12(8):e10086. doi: 10.7759/cureus.10086 | Wrong study design (narrative review) |
| Vacca A, Bulfone L, Cicco S, et al. Alcohol Intake and Arterial Hypertension: Retelling of a Multifaceted Story. *Nutrients*. 2023;15(4)doi:10.3390/nu15040958 | Wrong study design (narrative review) |

**Supplementary Table S2:**
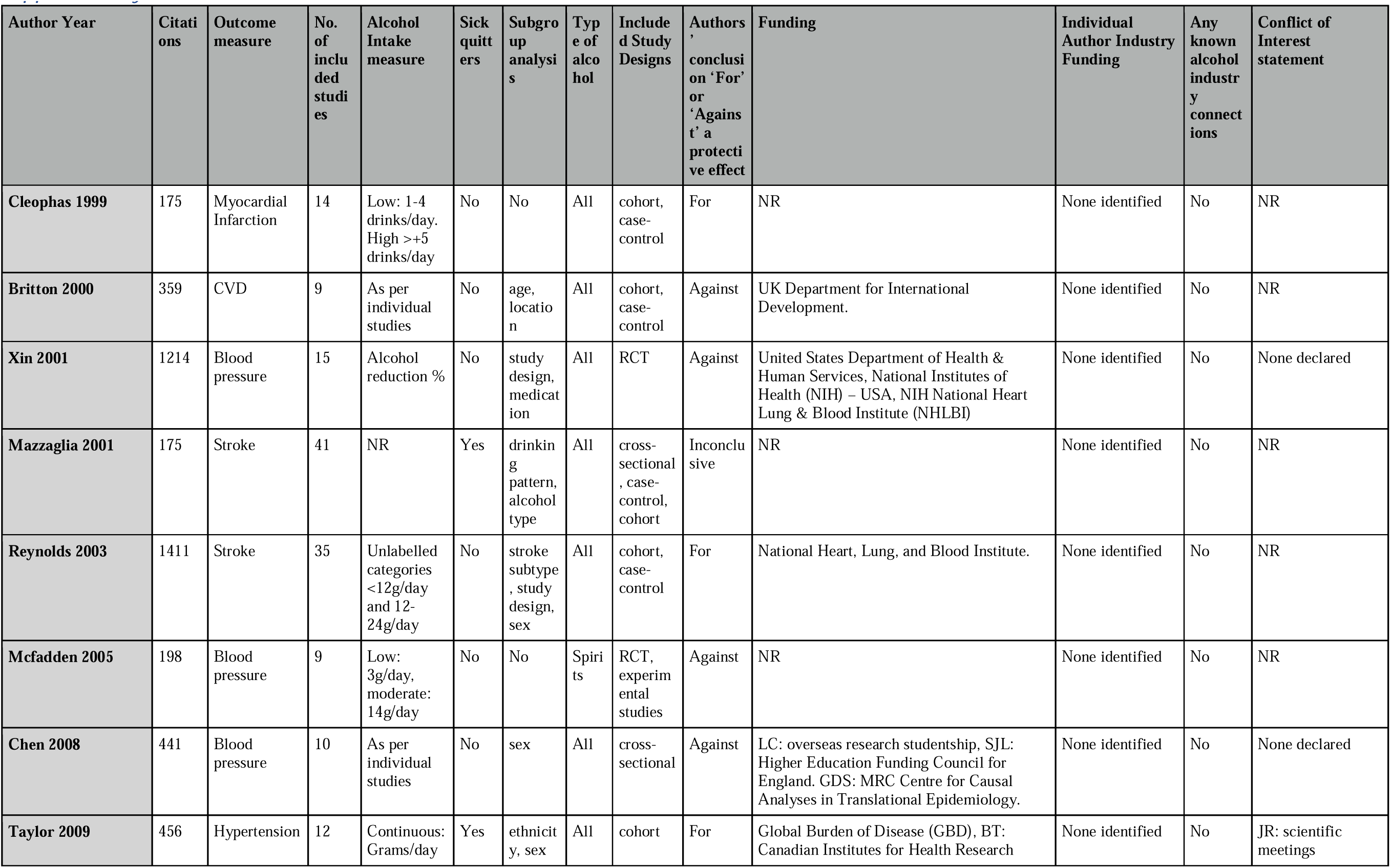
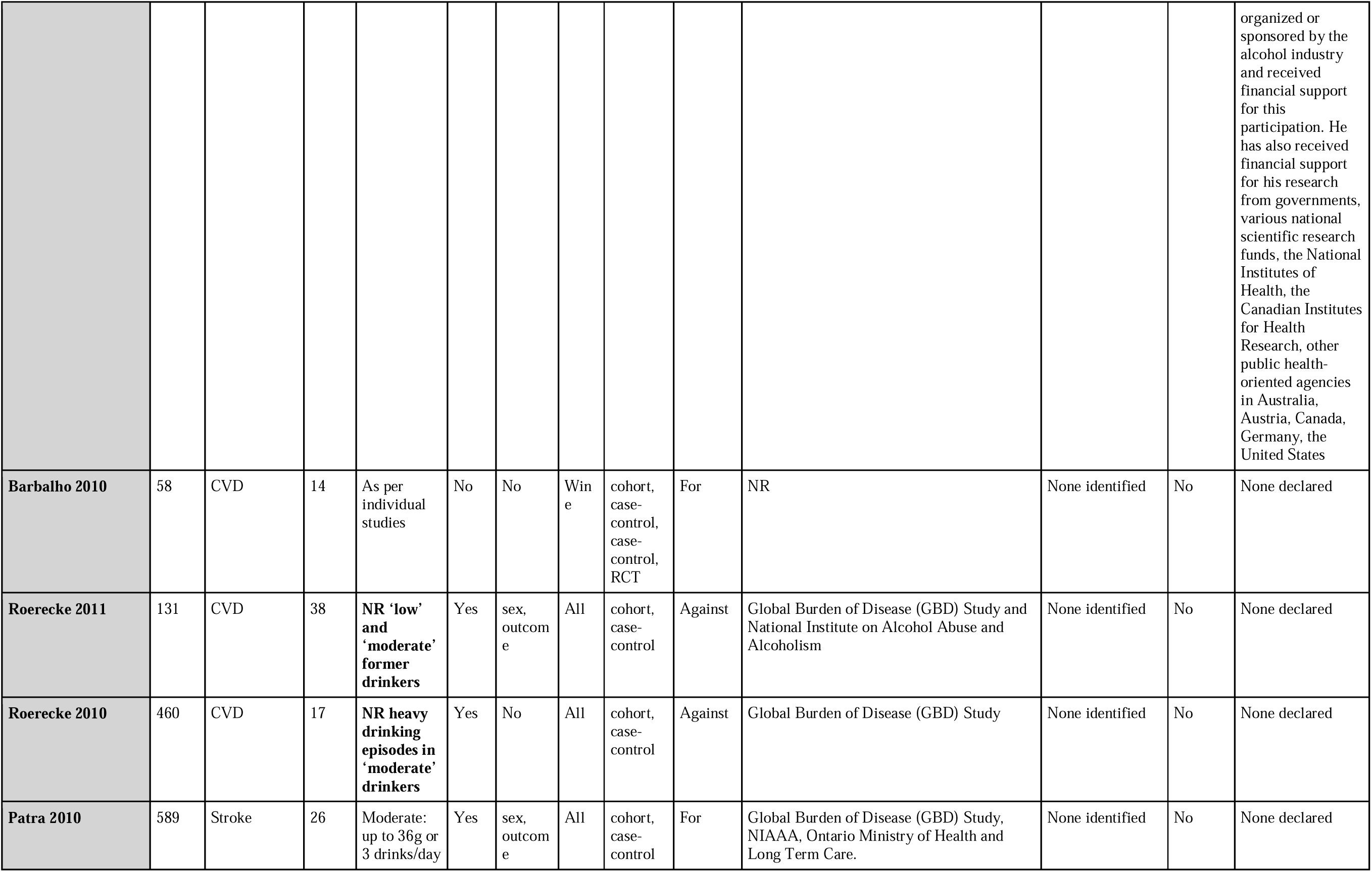
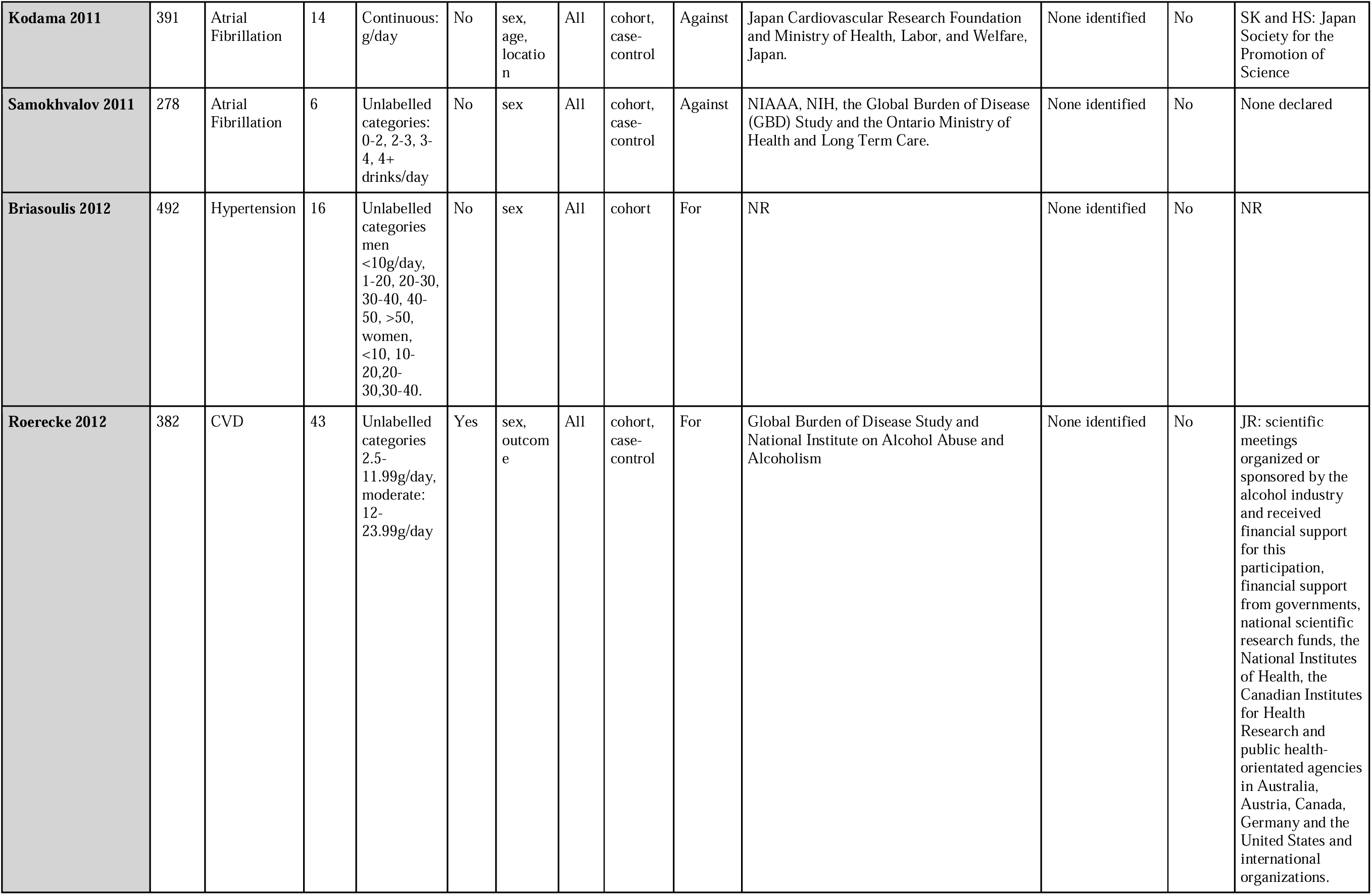
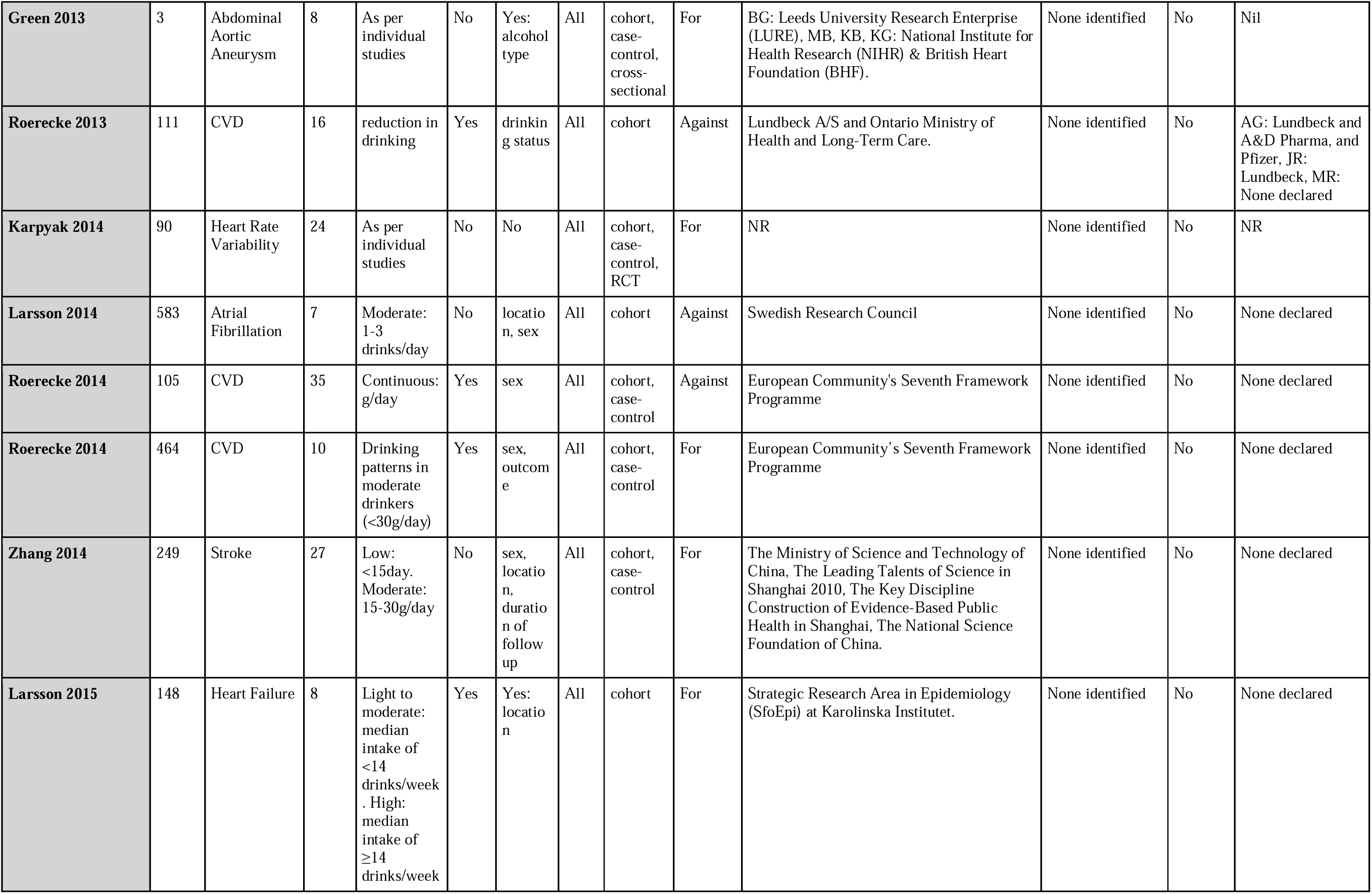
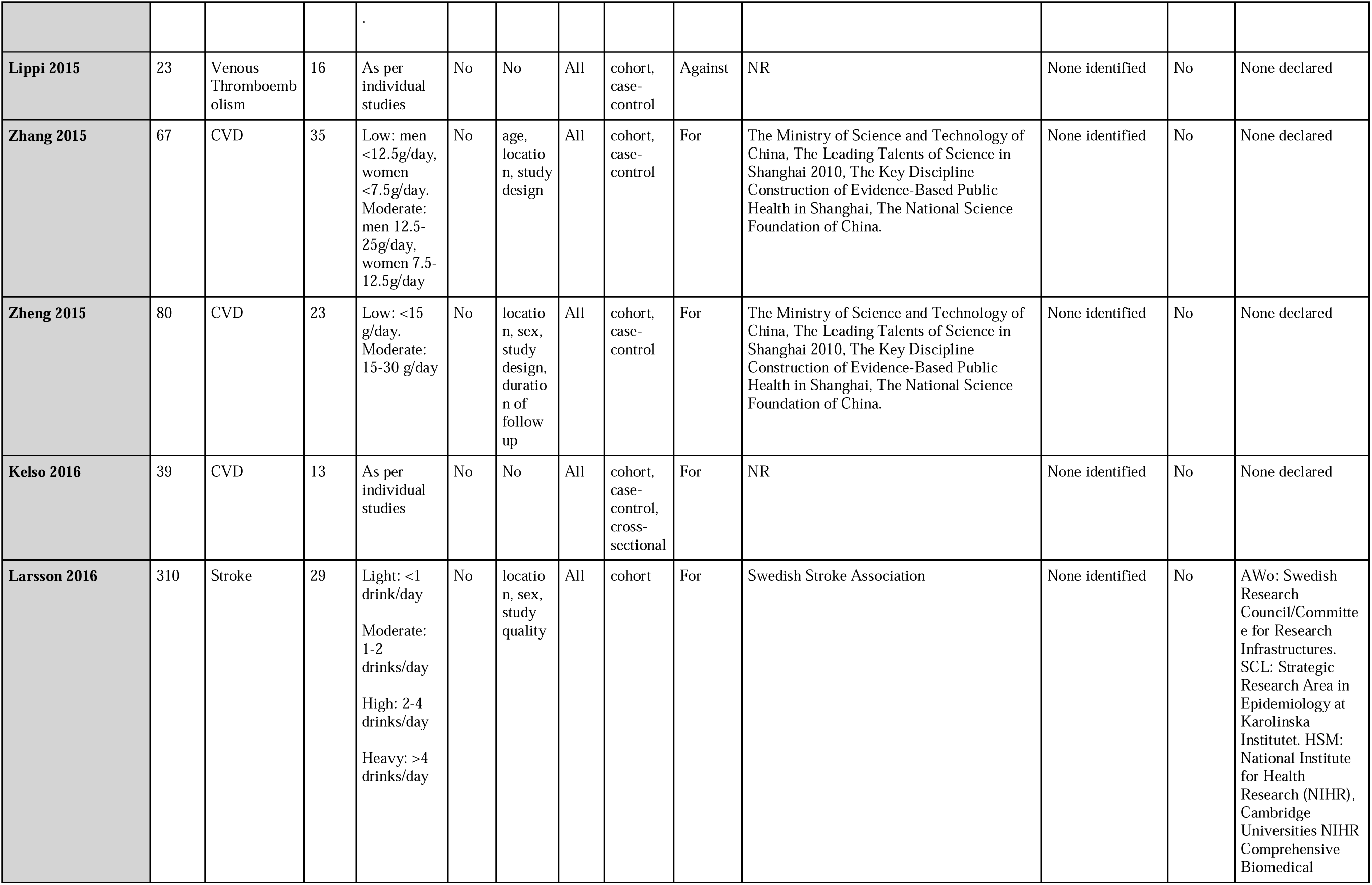
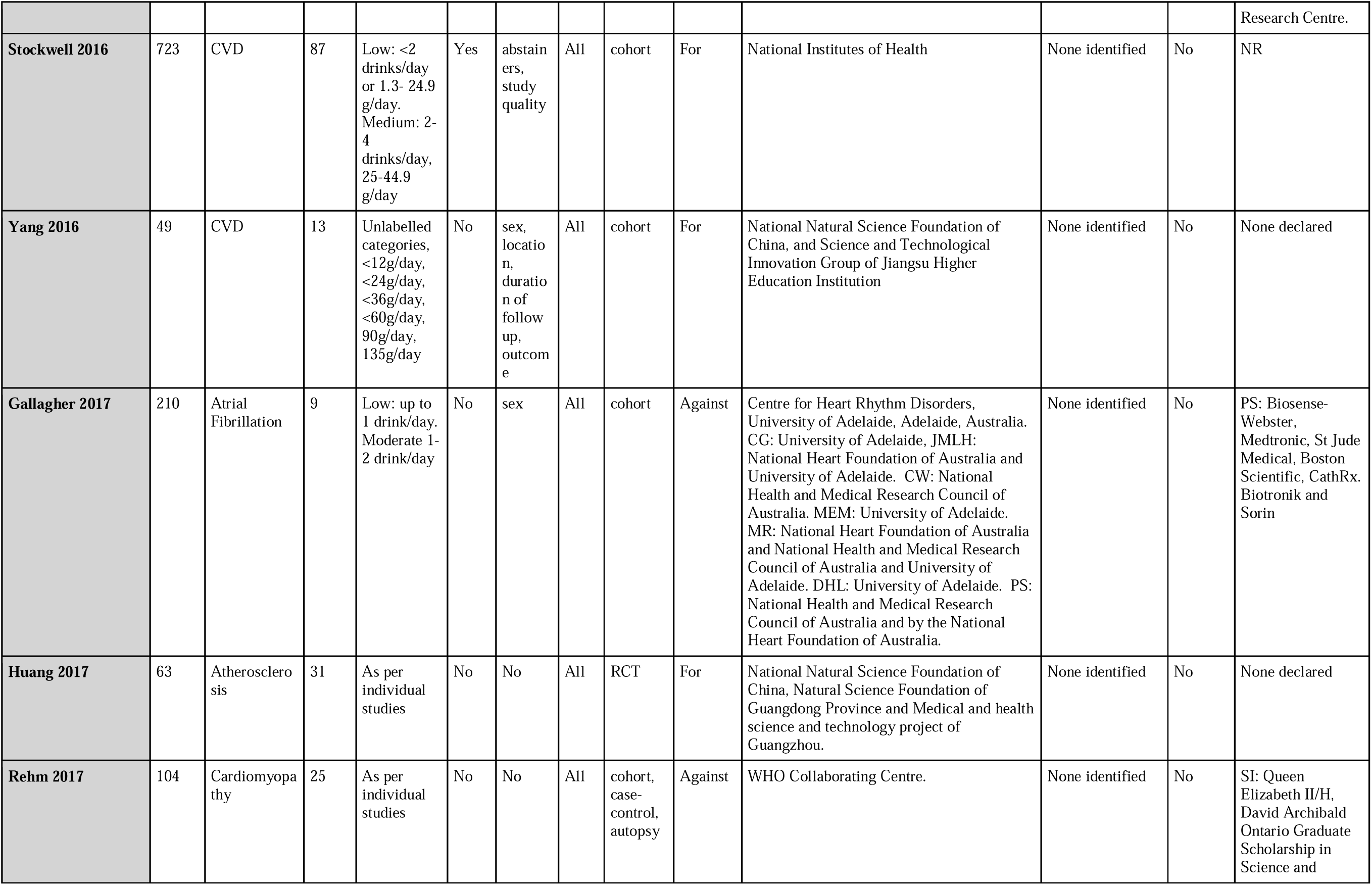
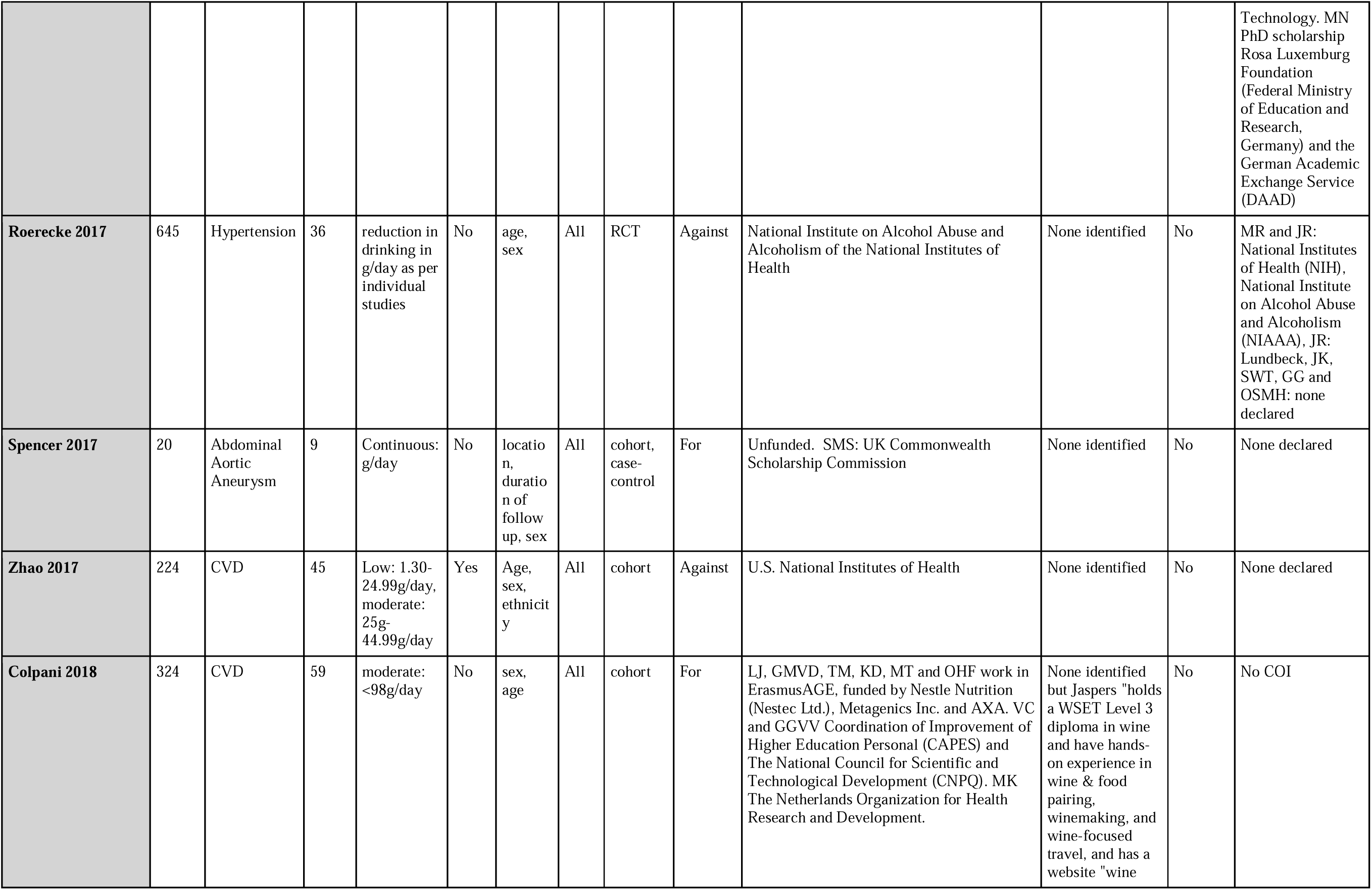
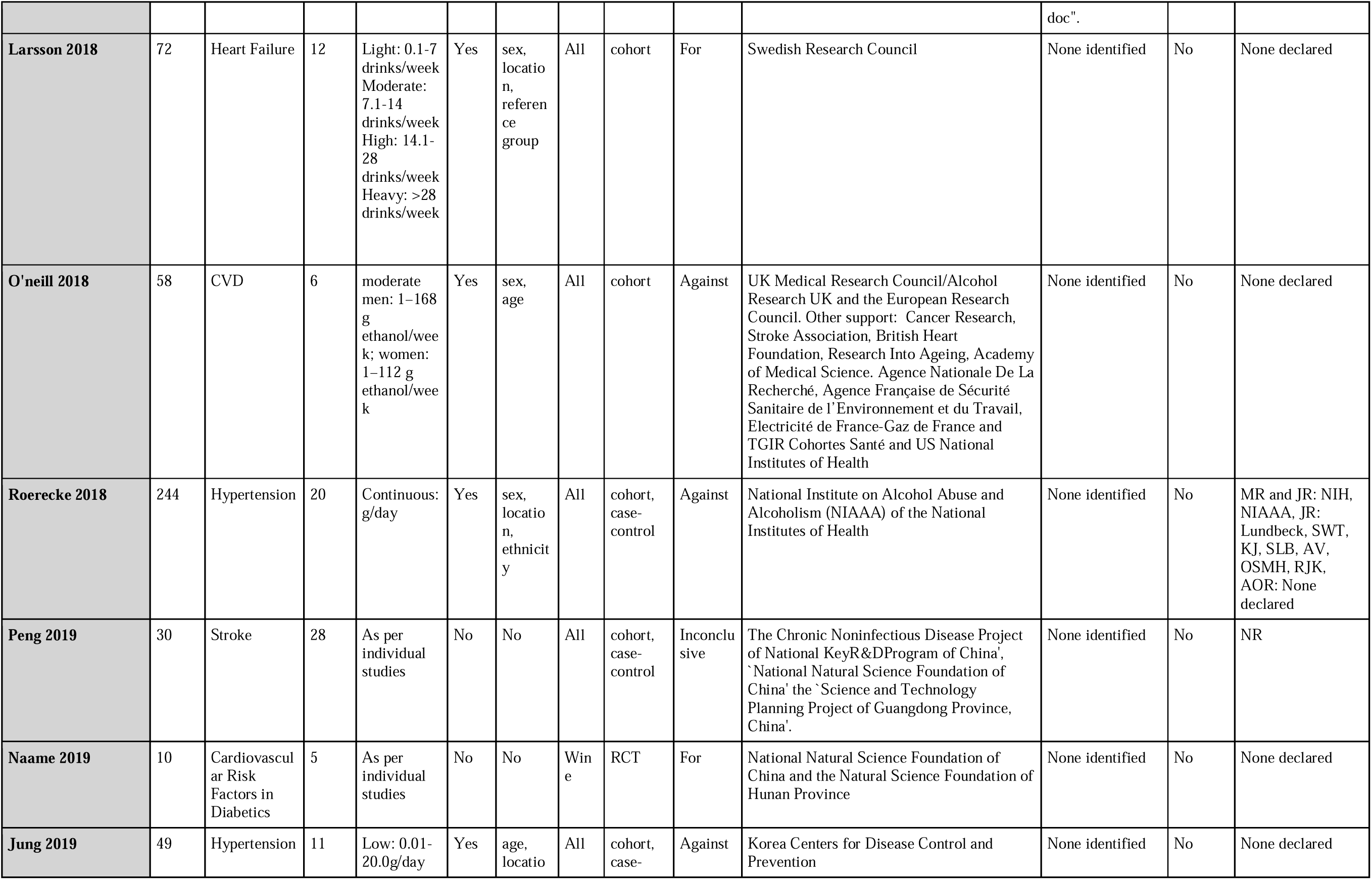
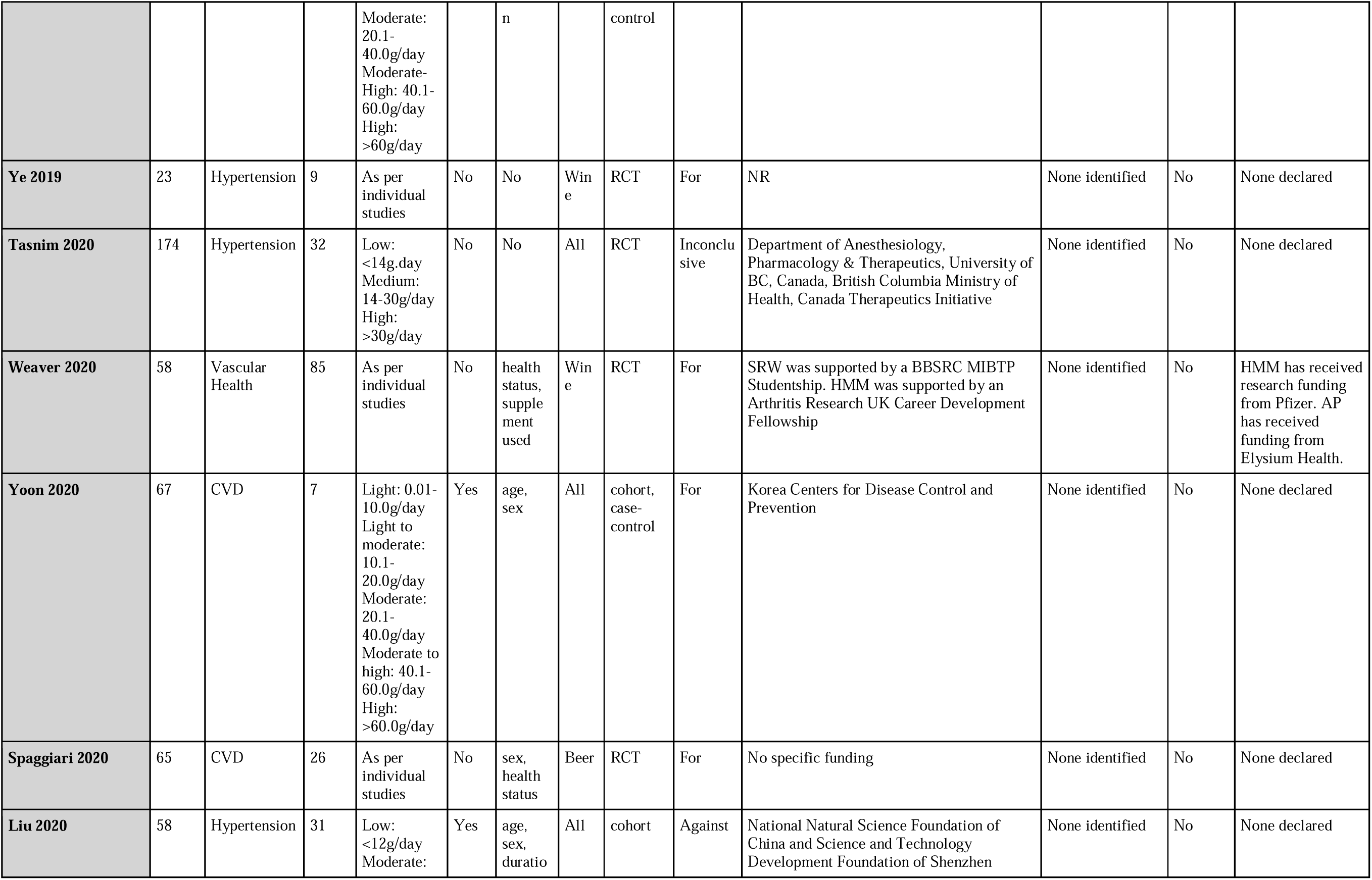
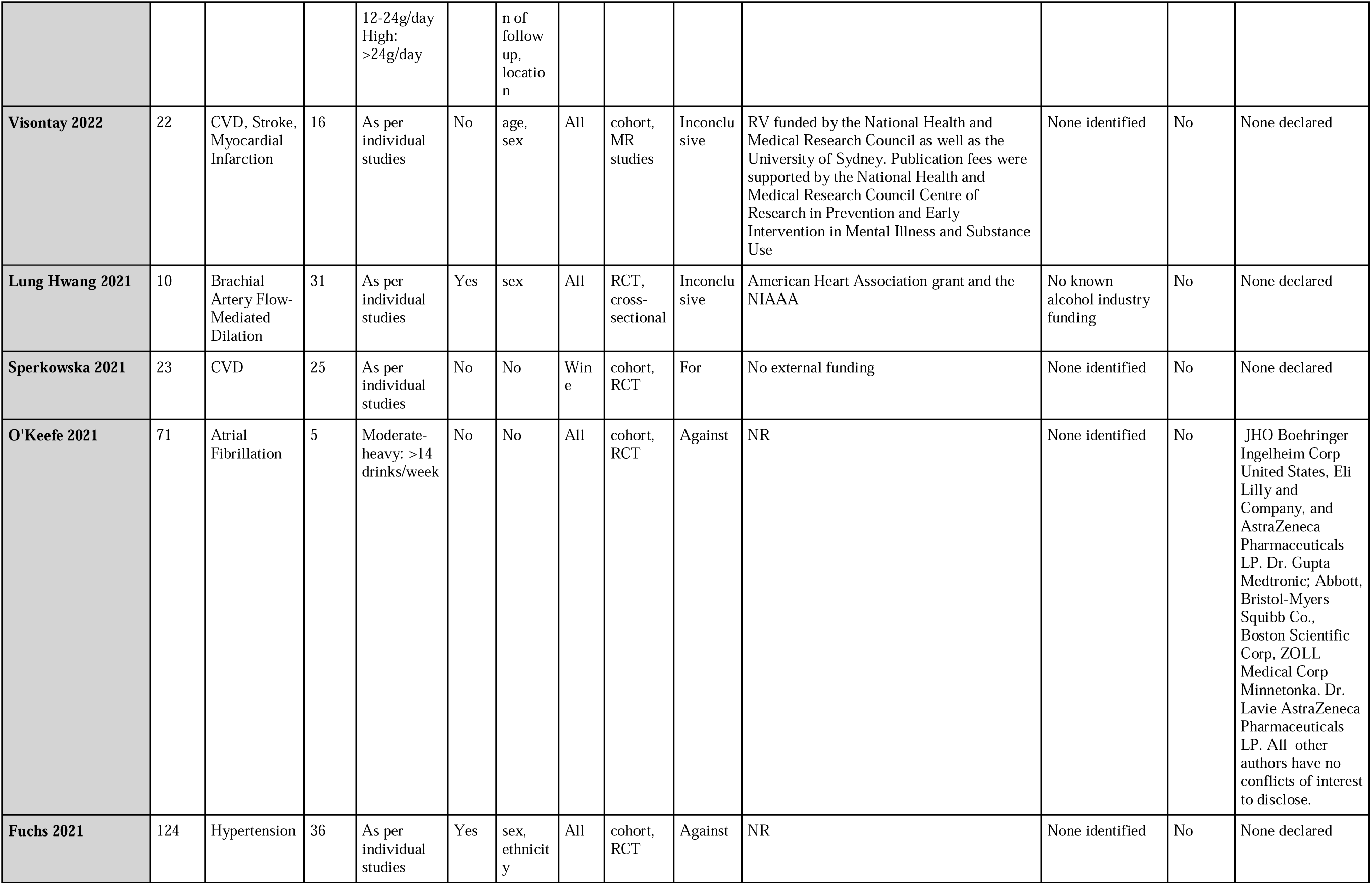
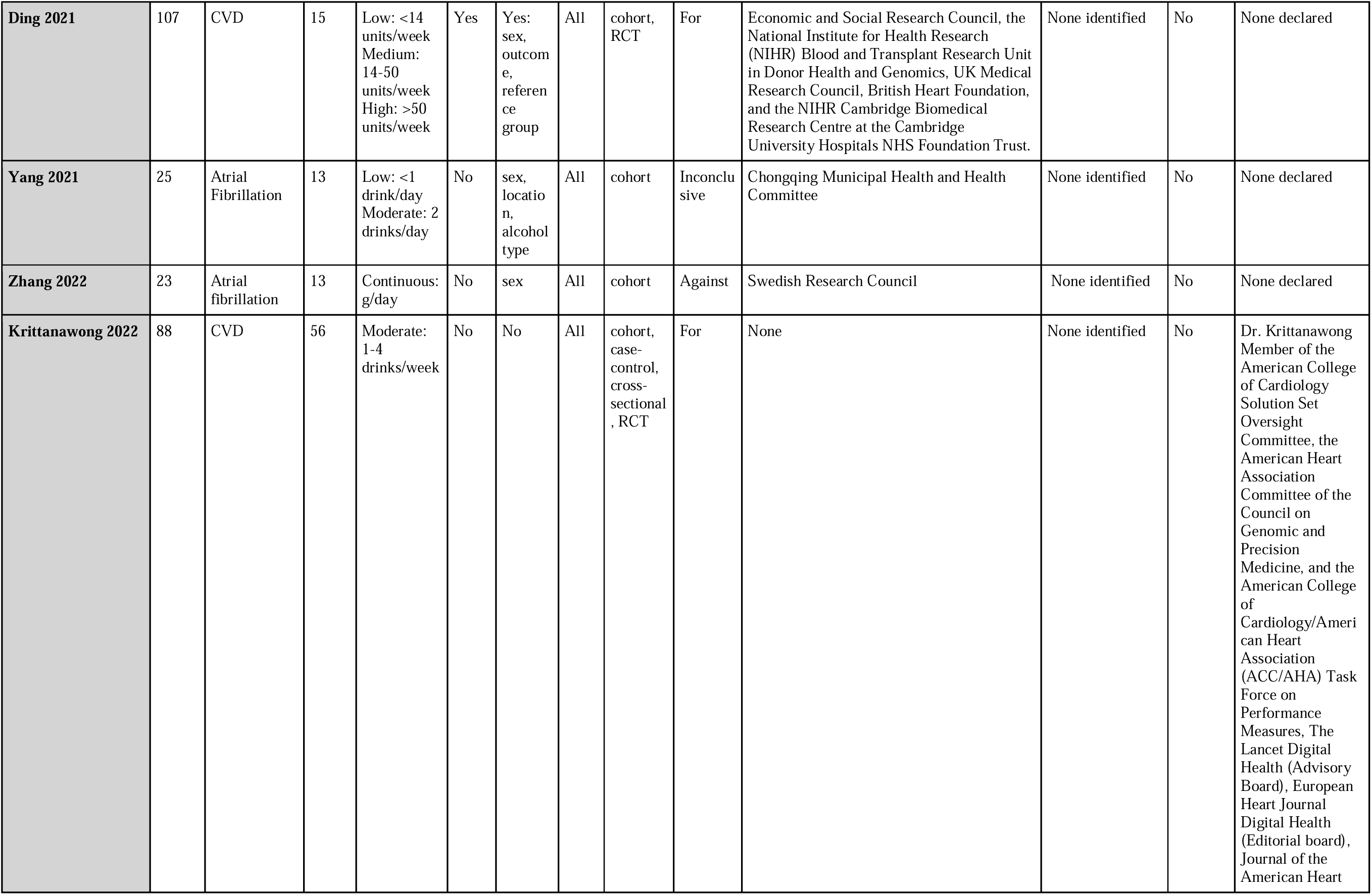
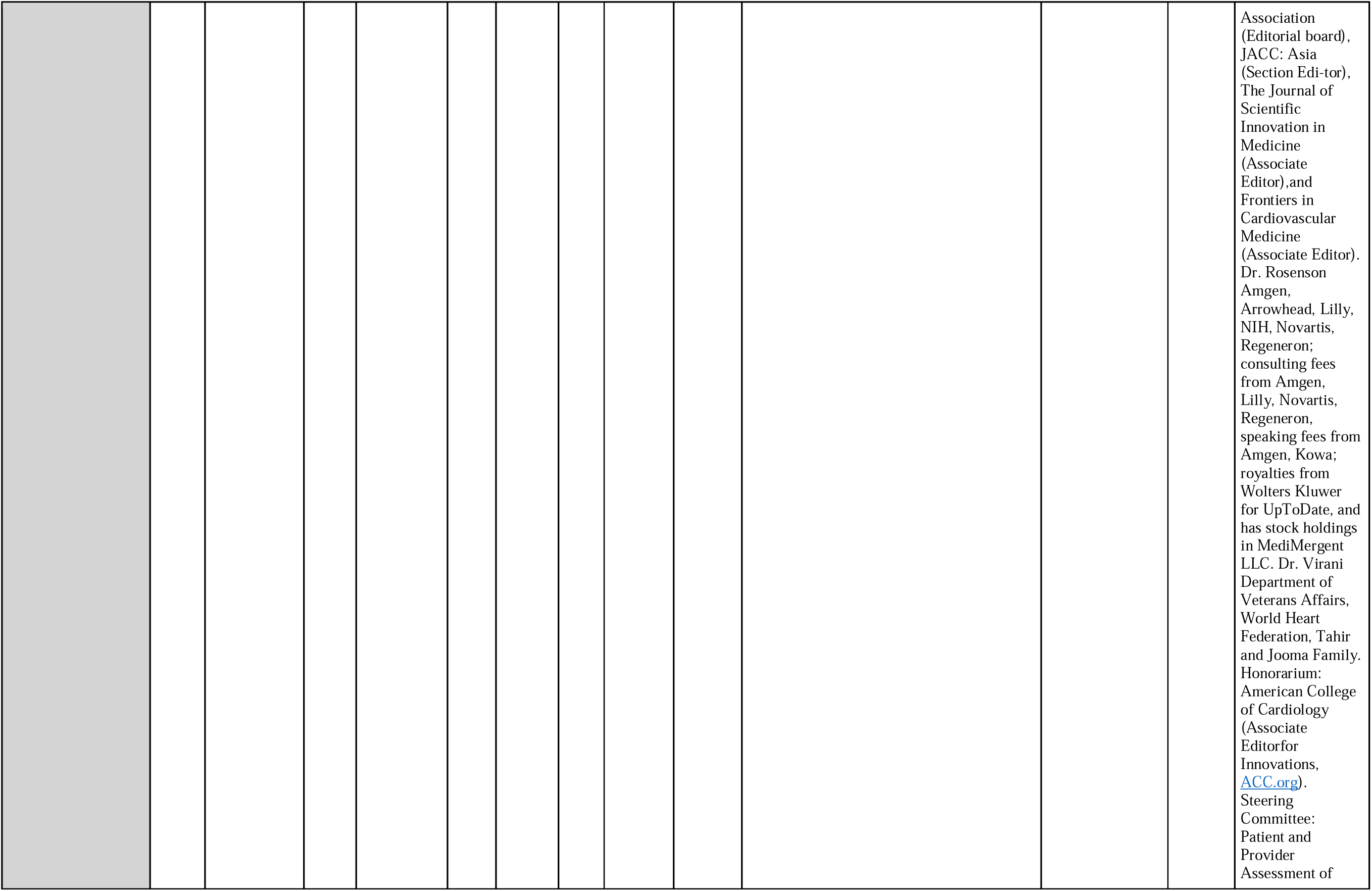
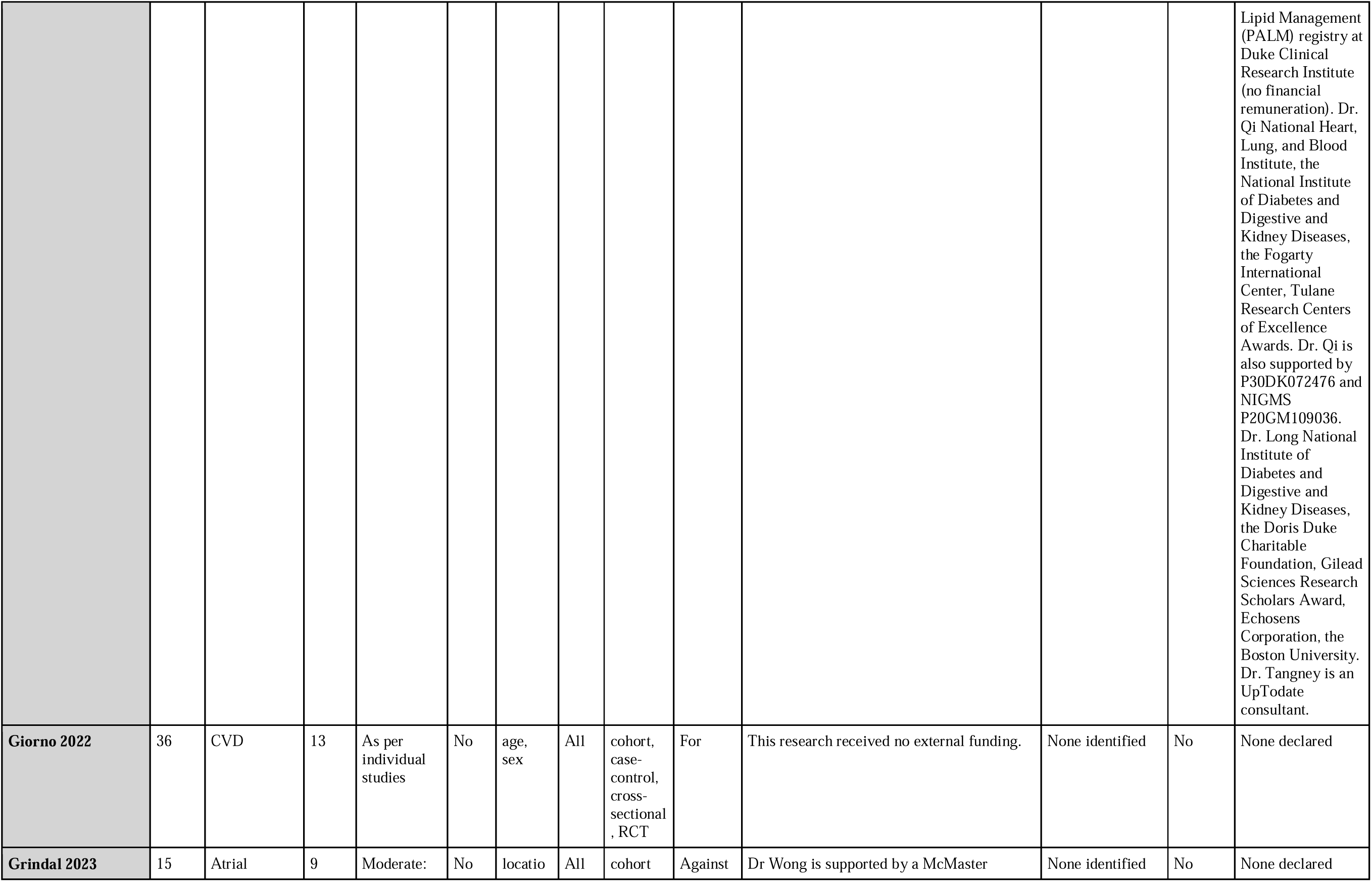
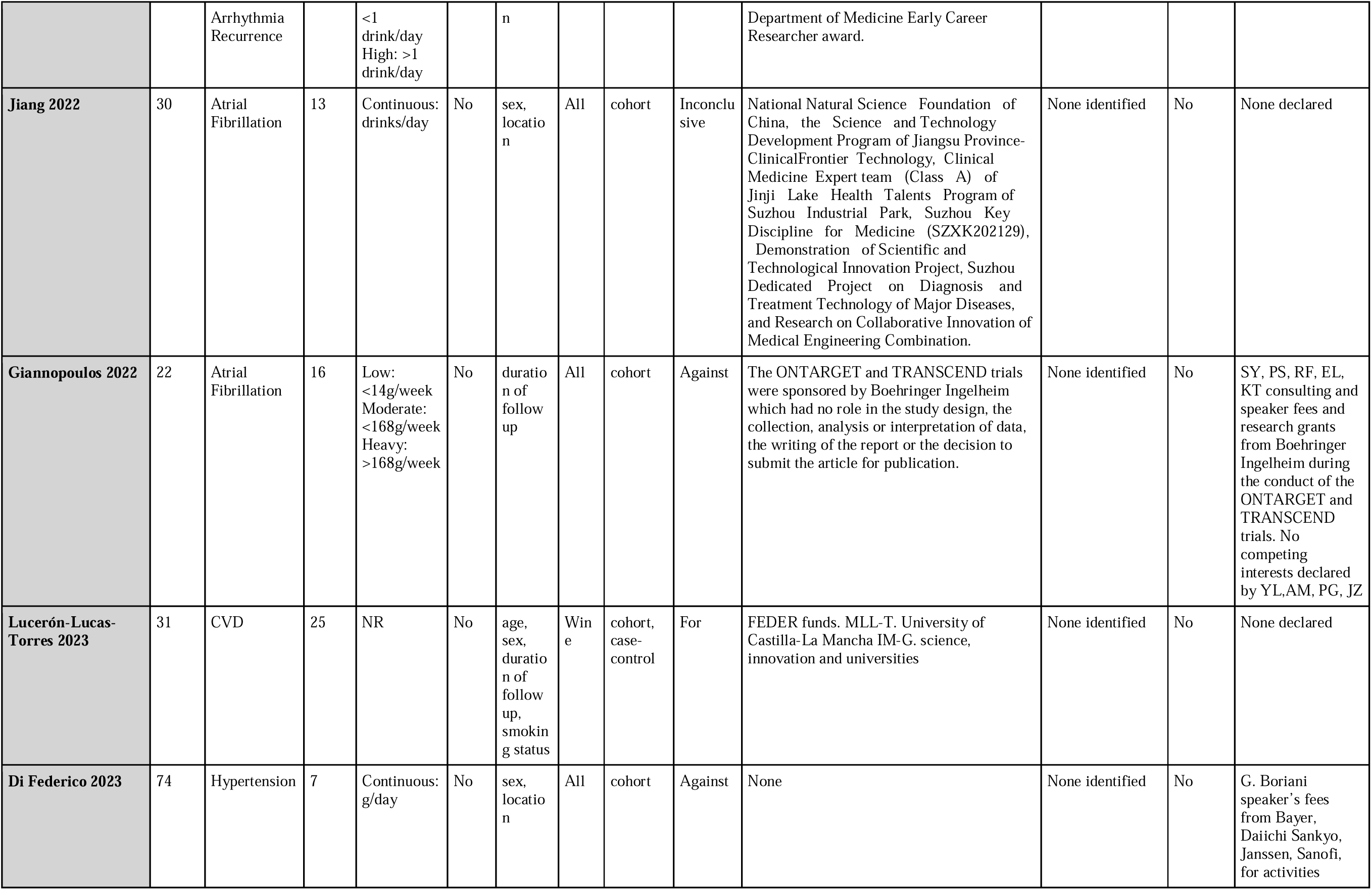
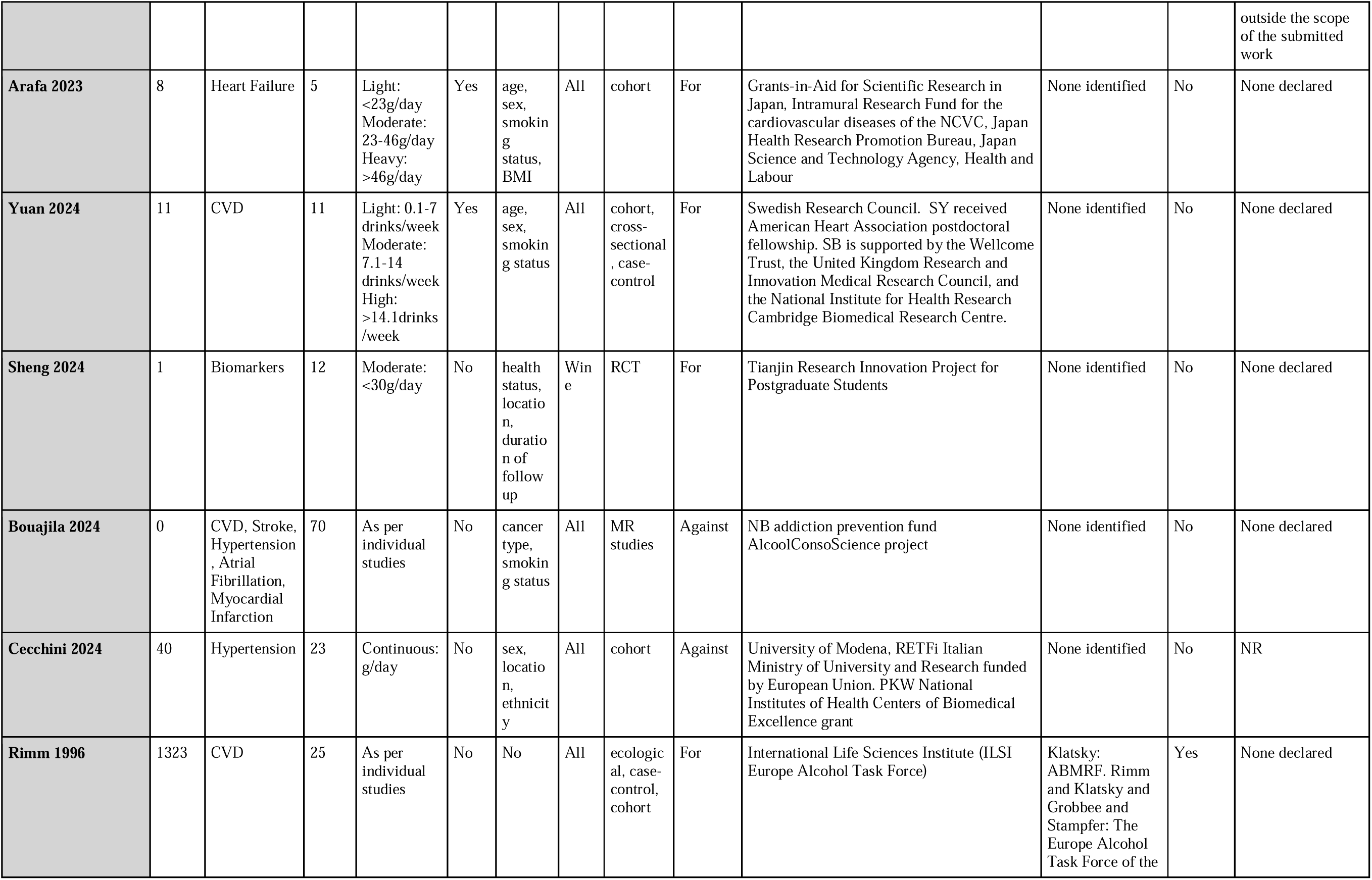
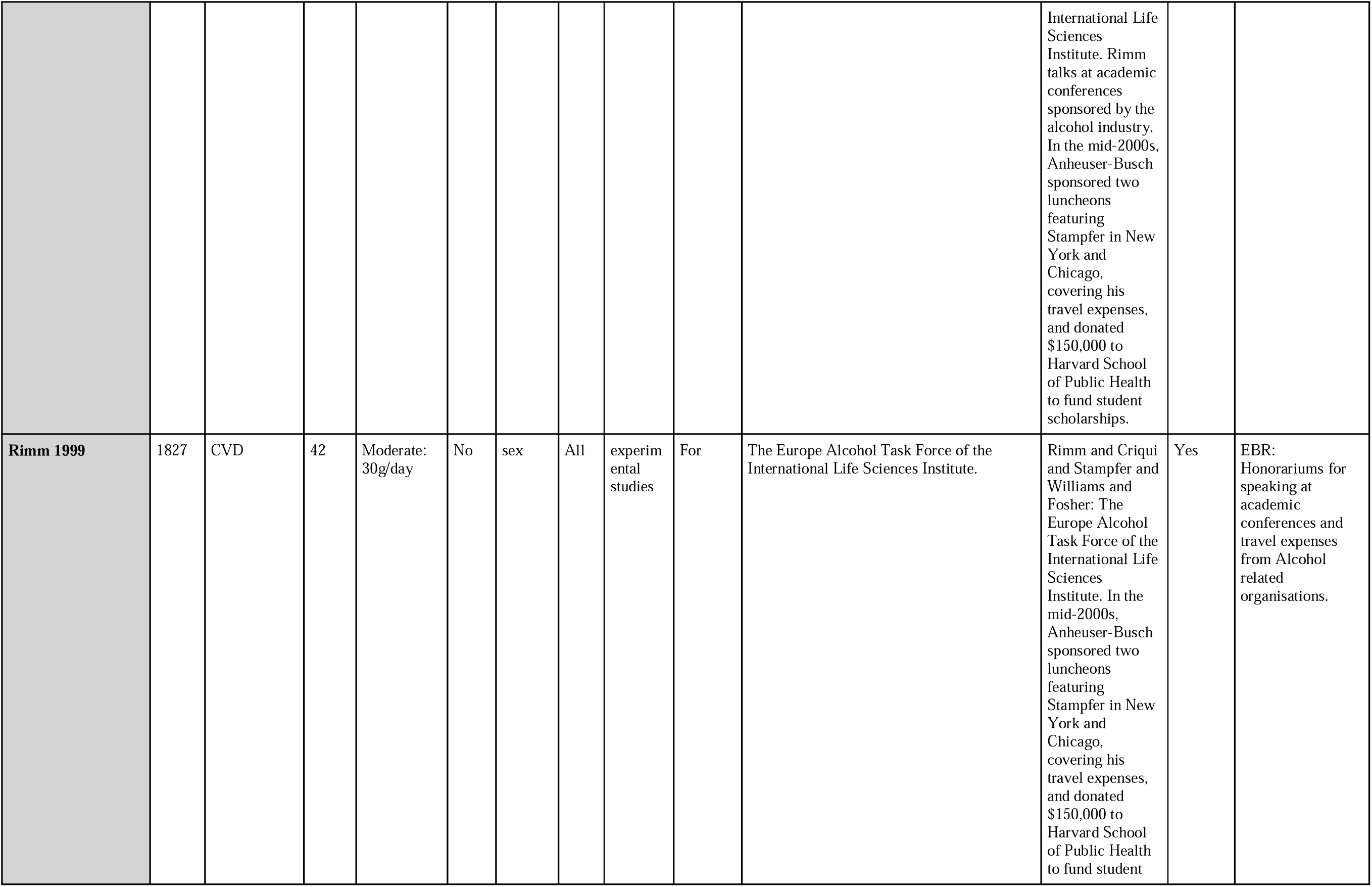
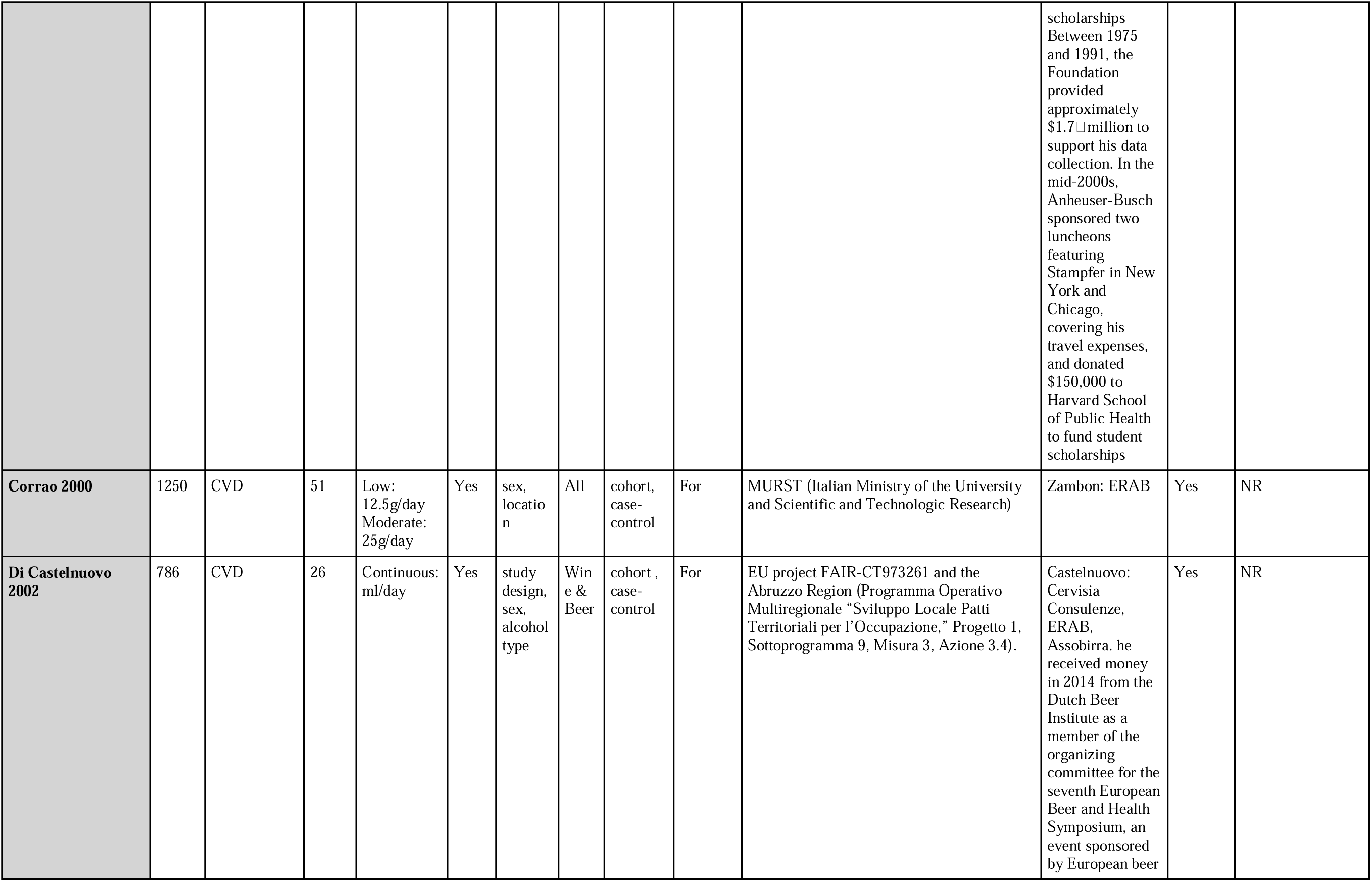
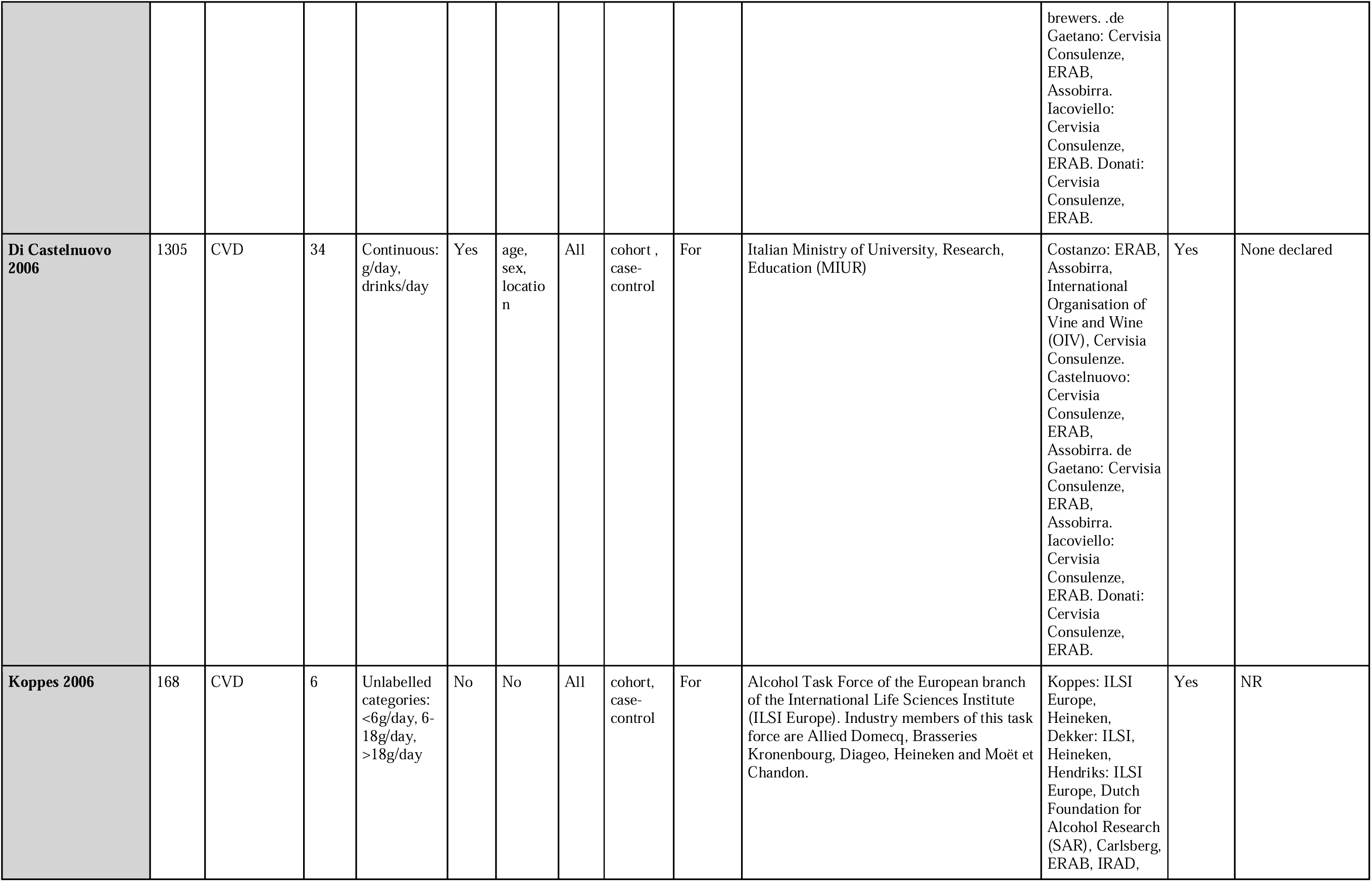
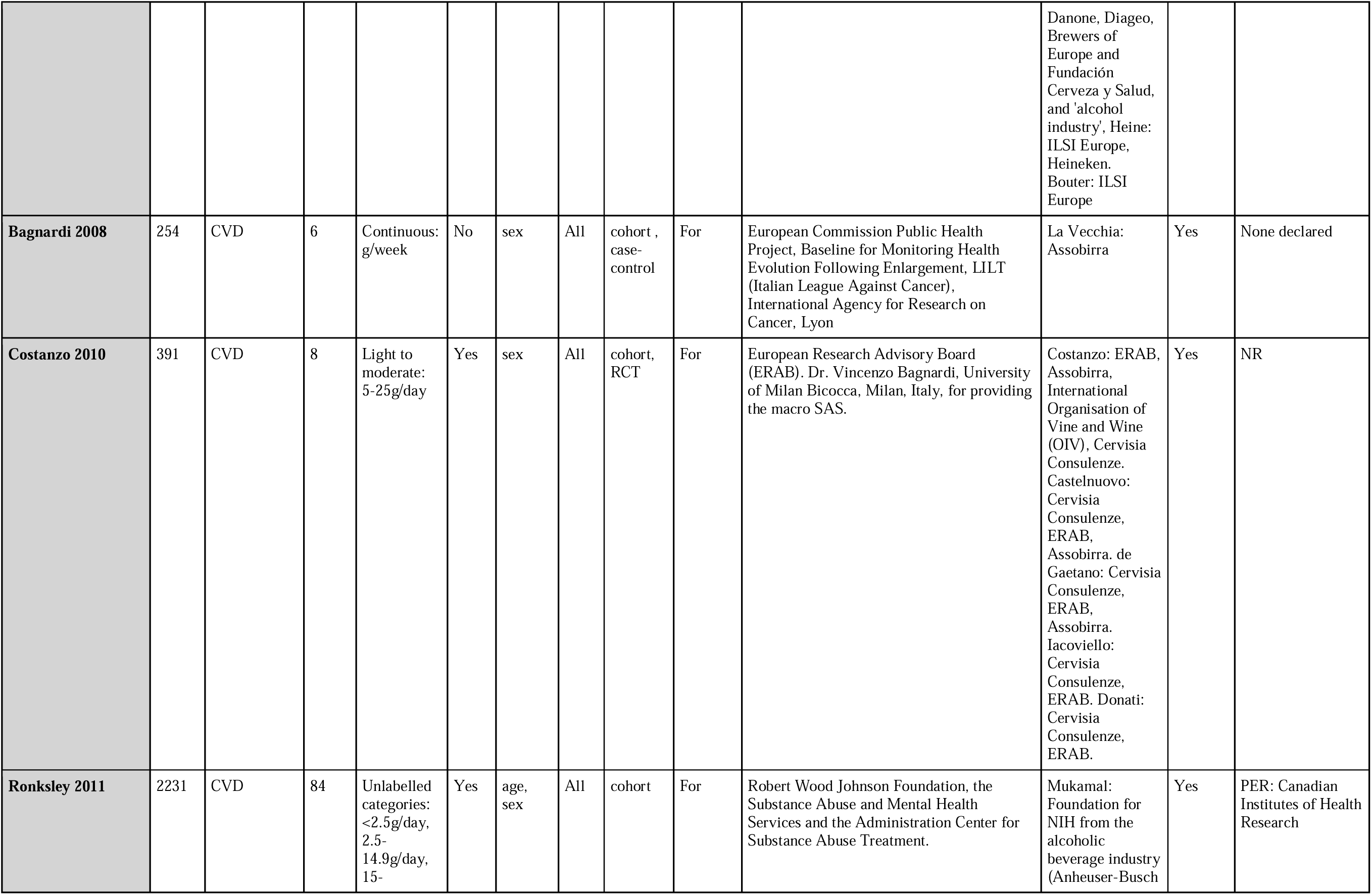
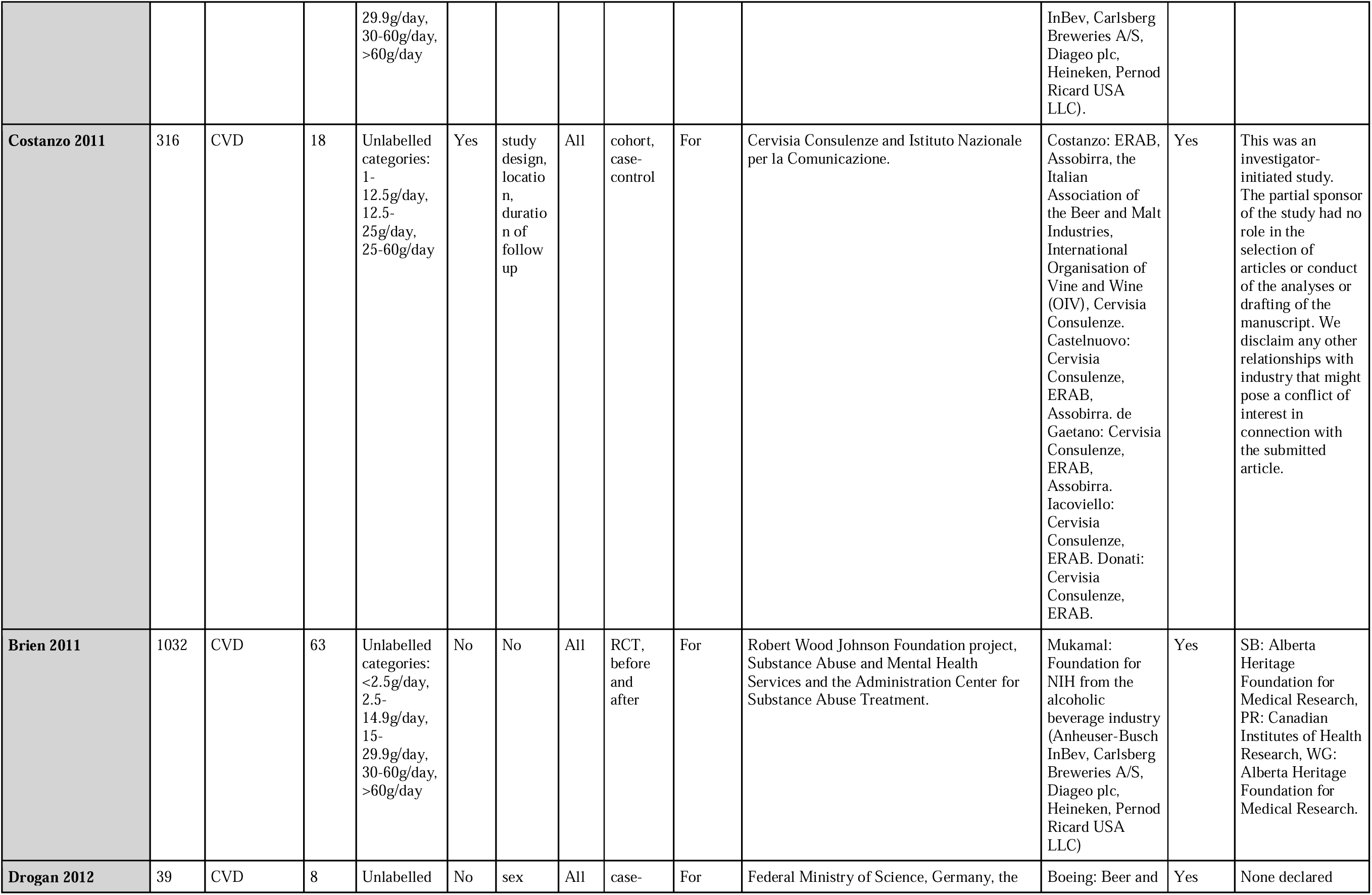
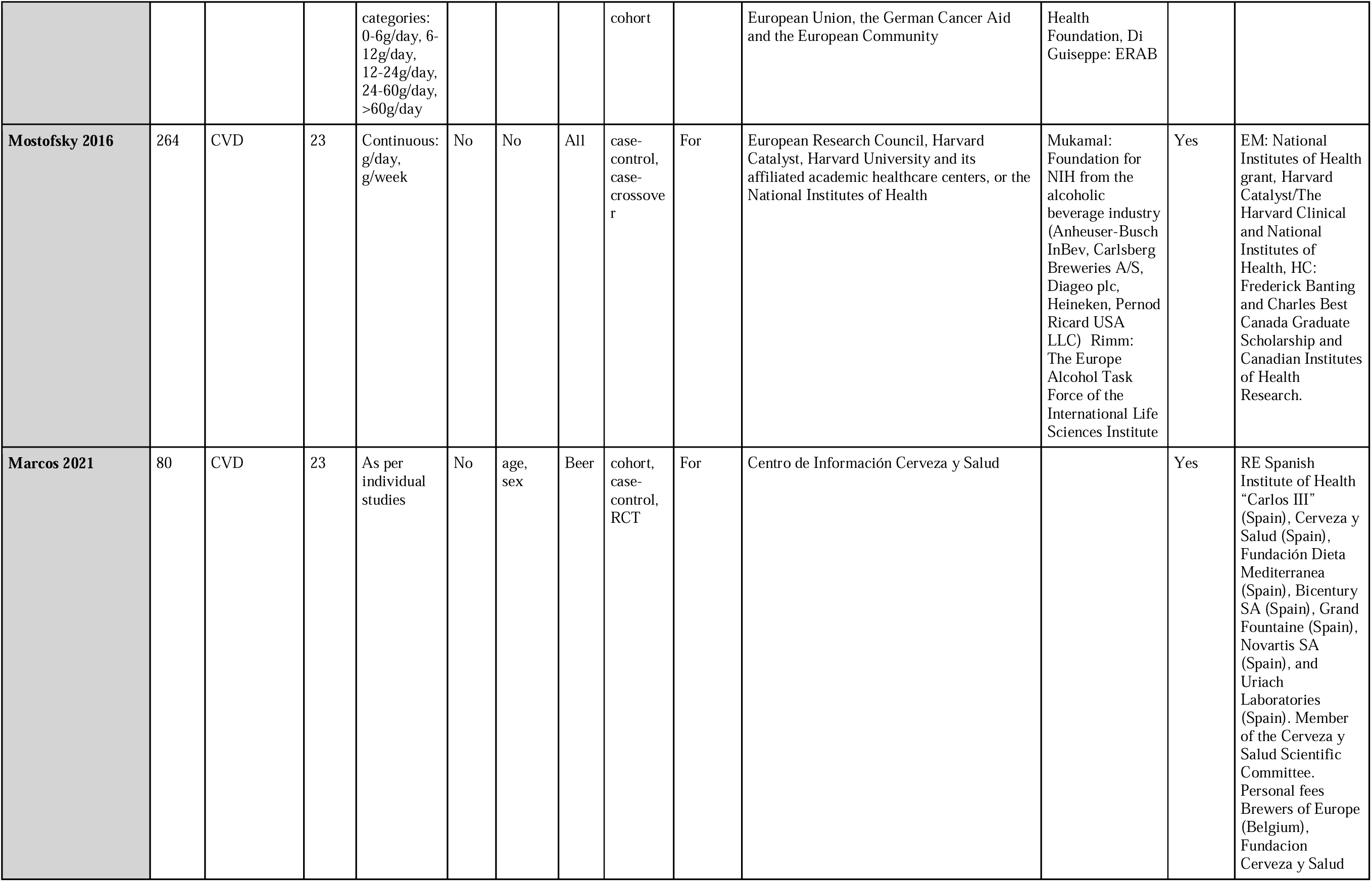
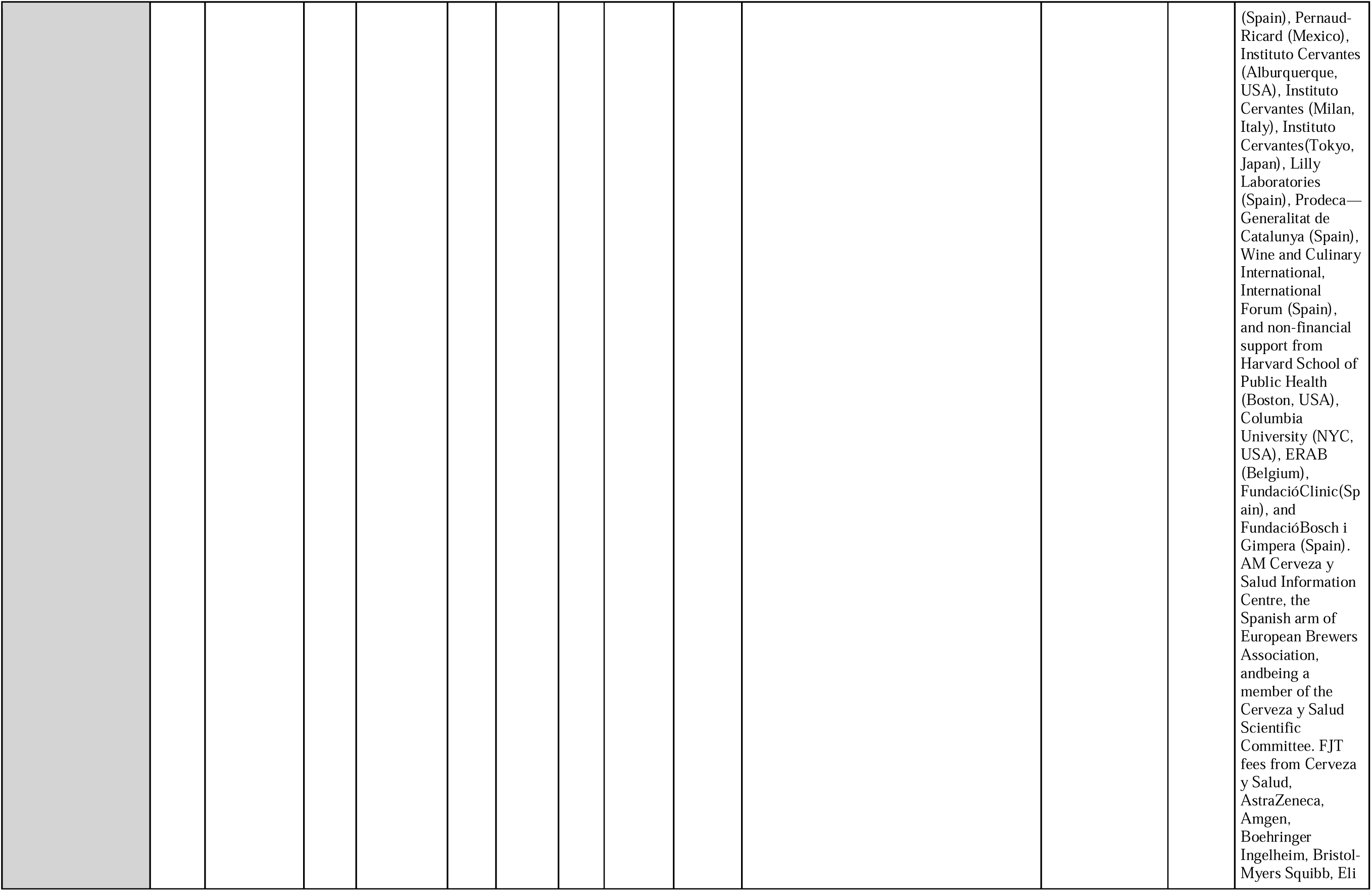
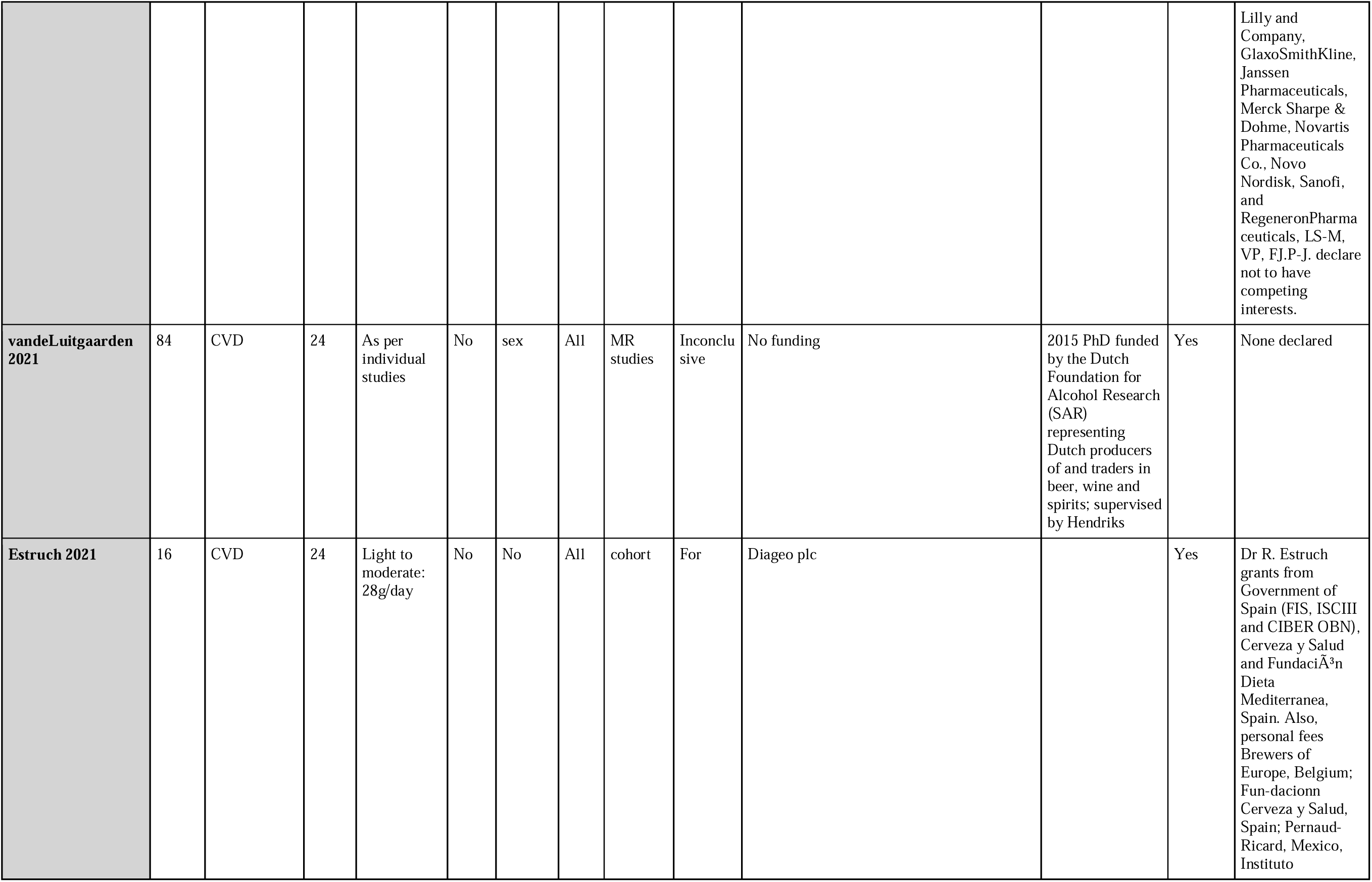
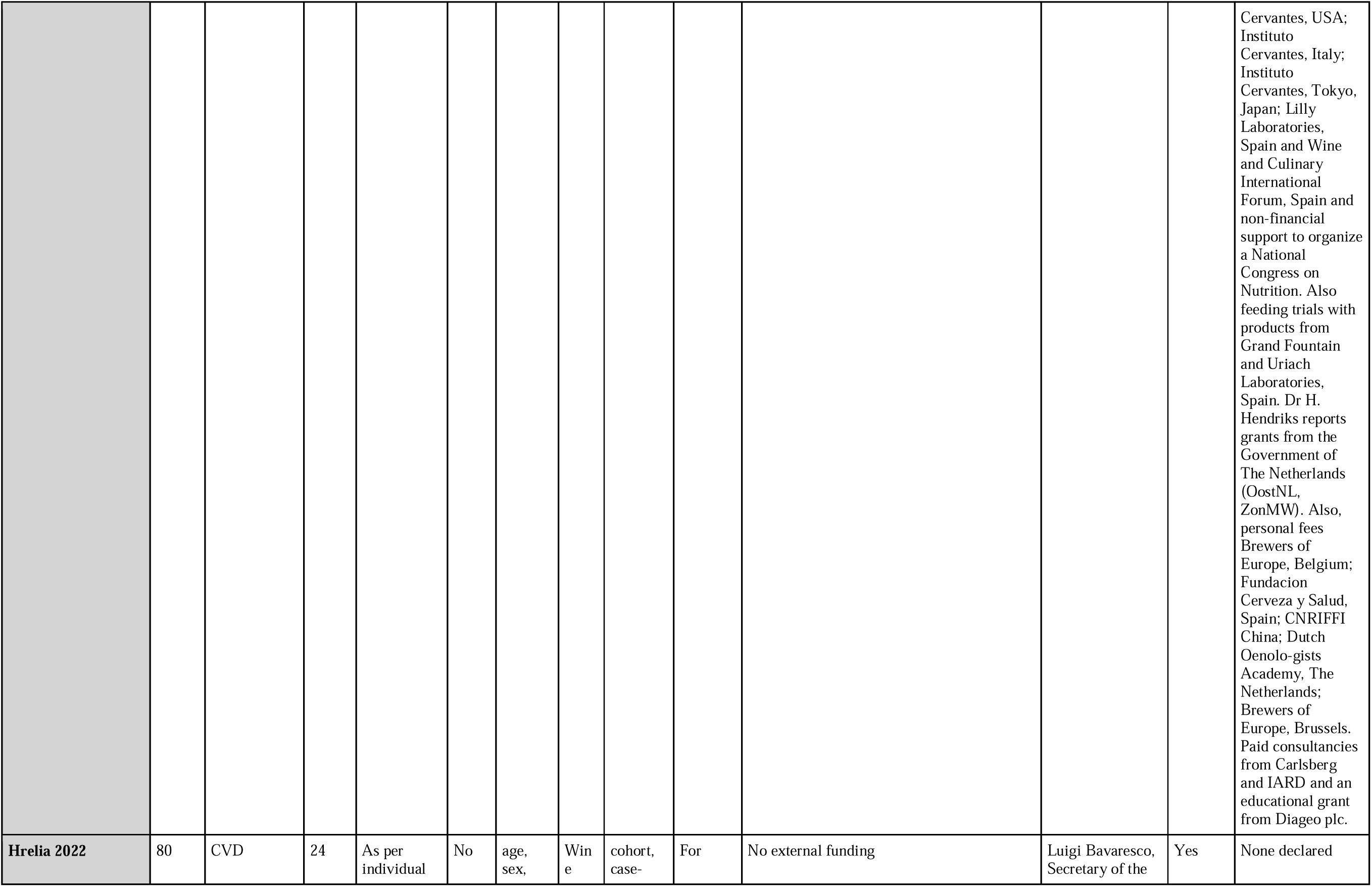
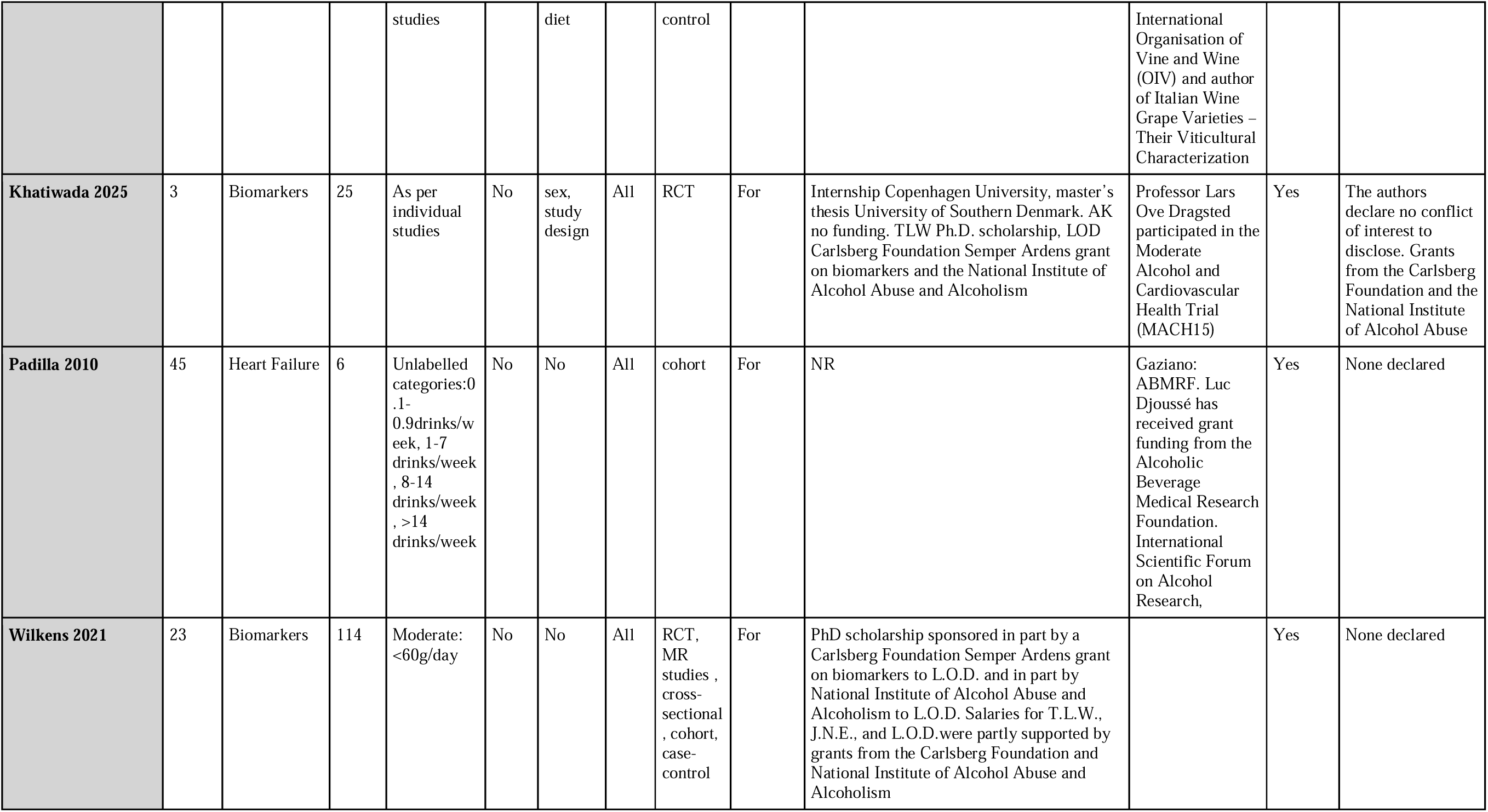
Characteristics of Included Studies.

**Supplementary Table S3:**
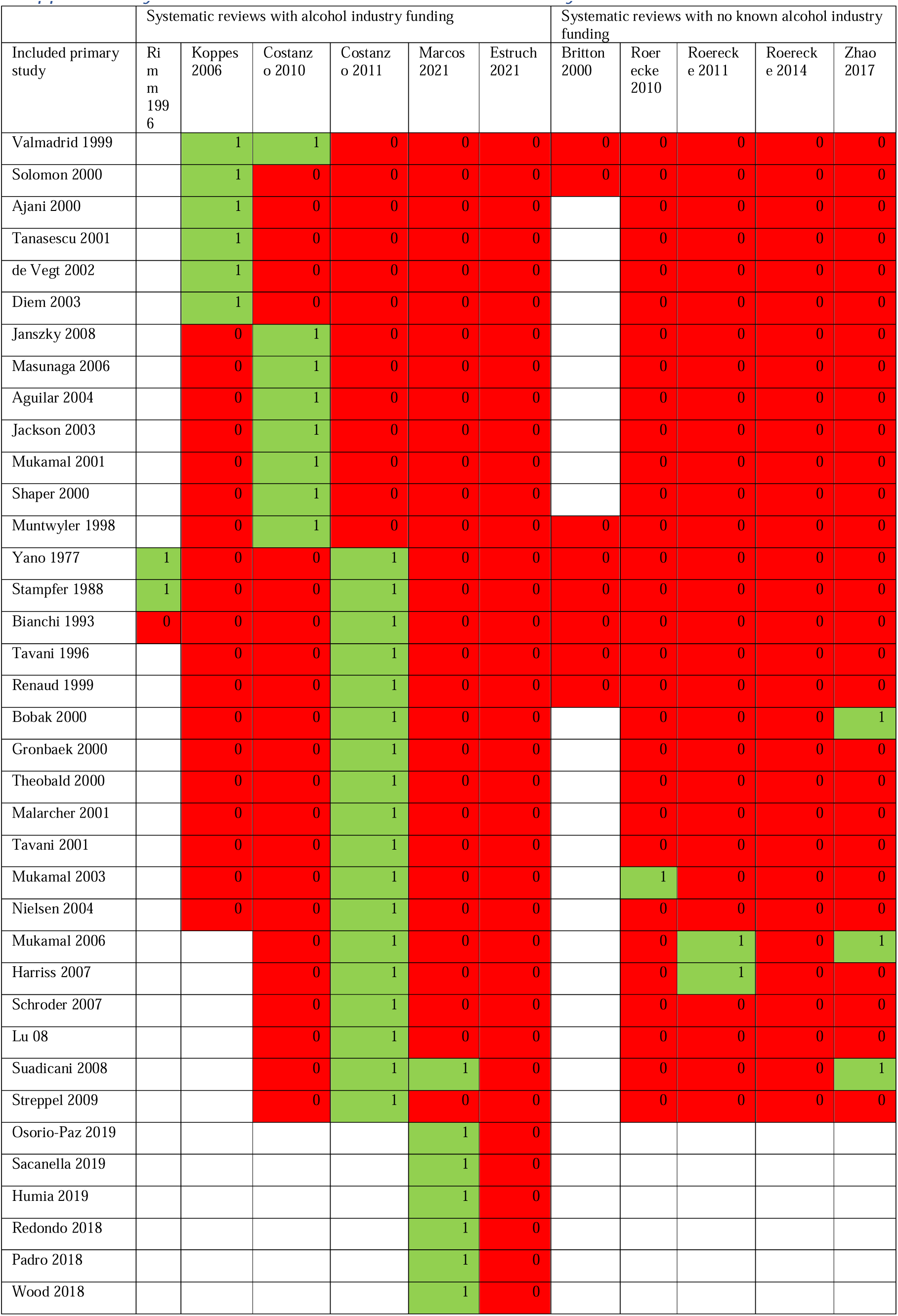
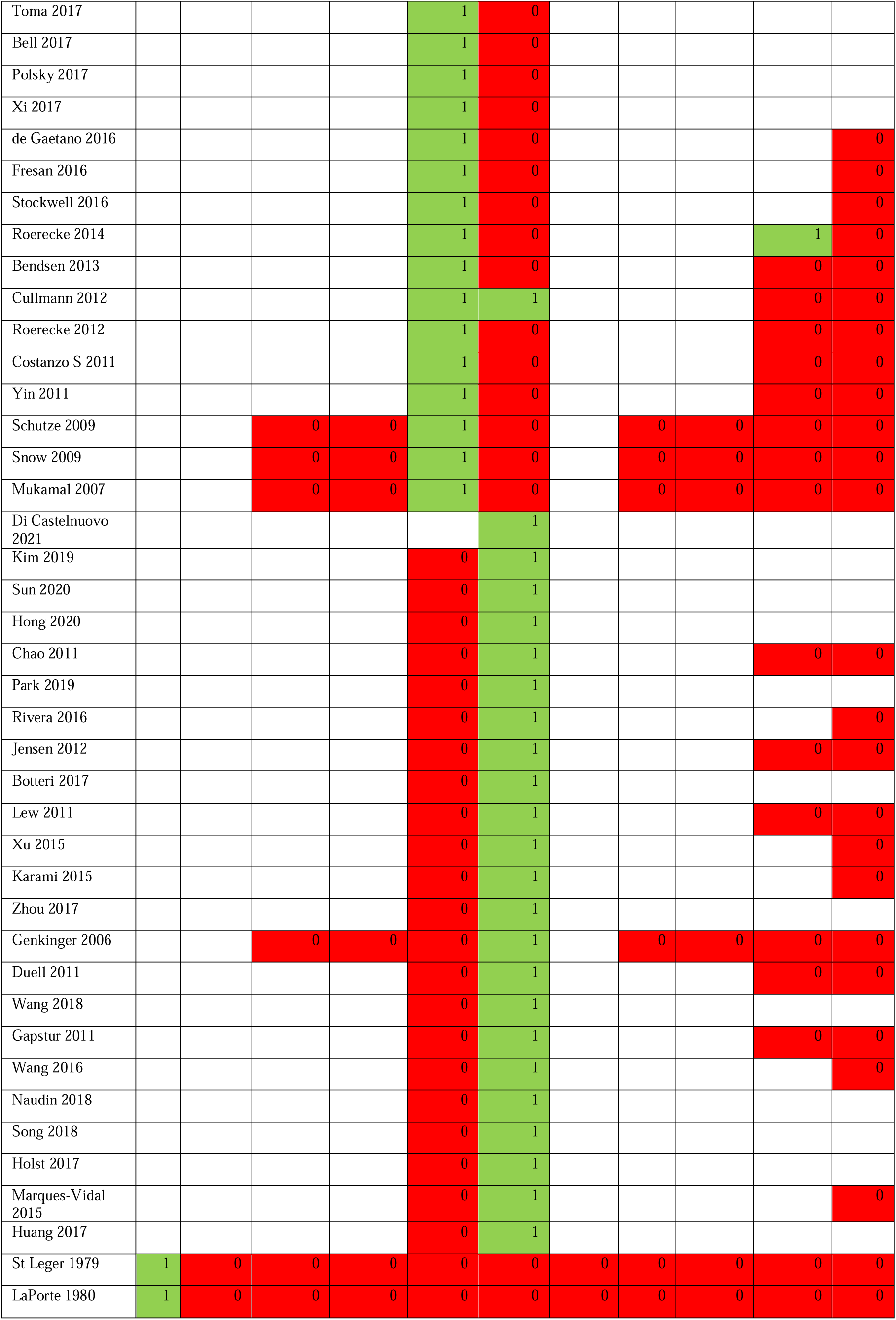
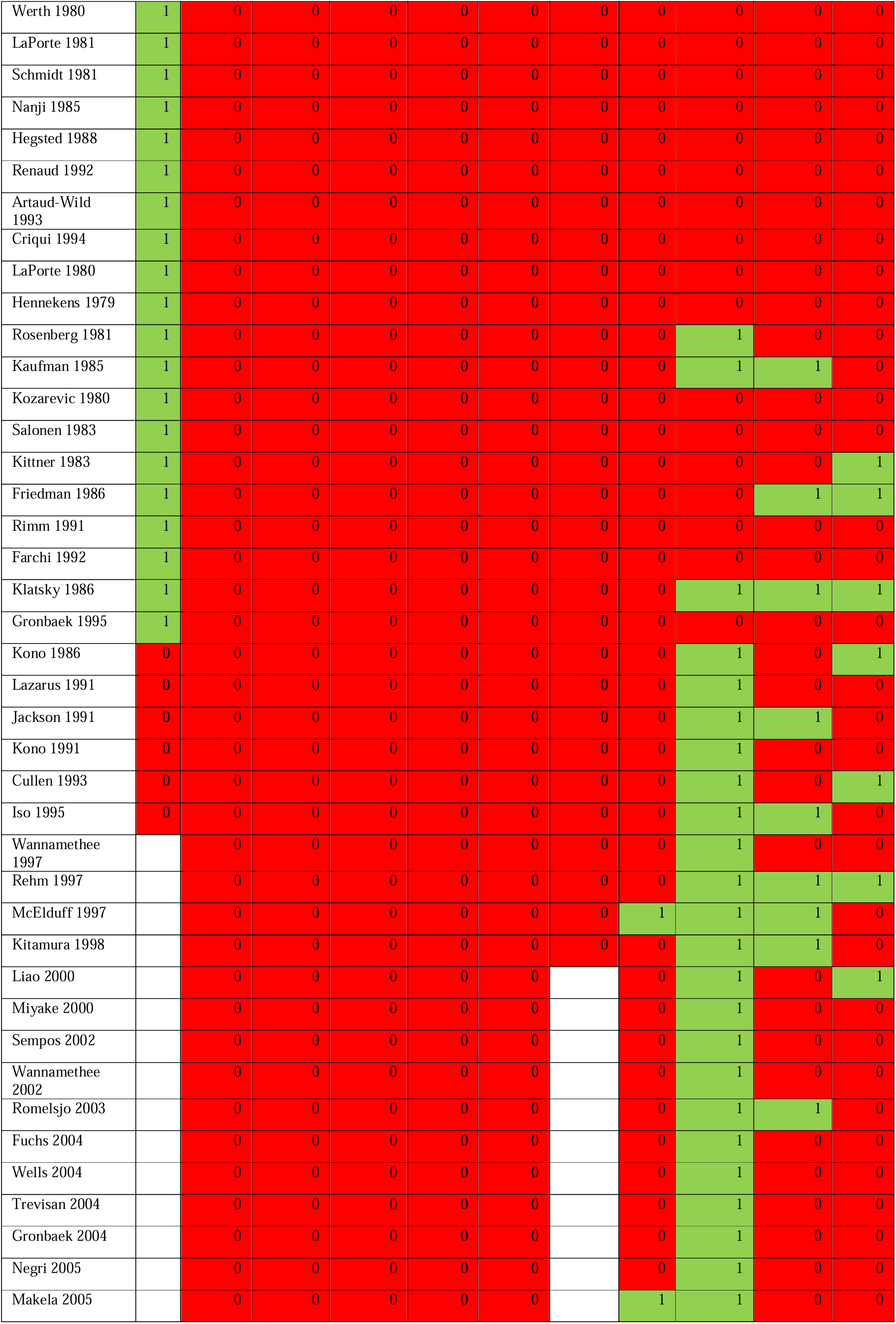
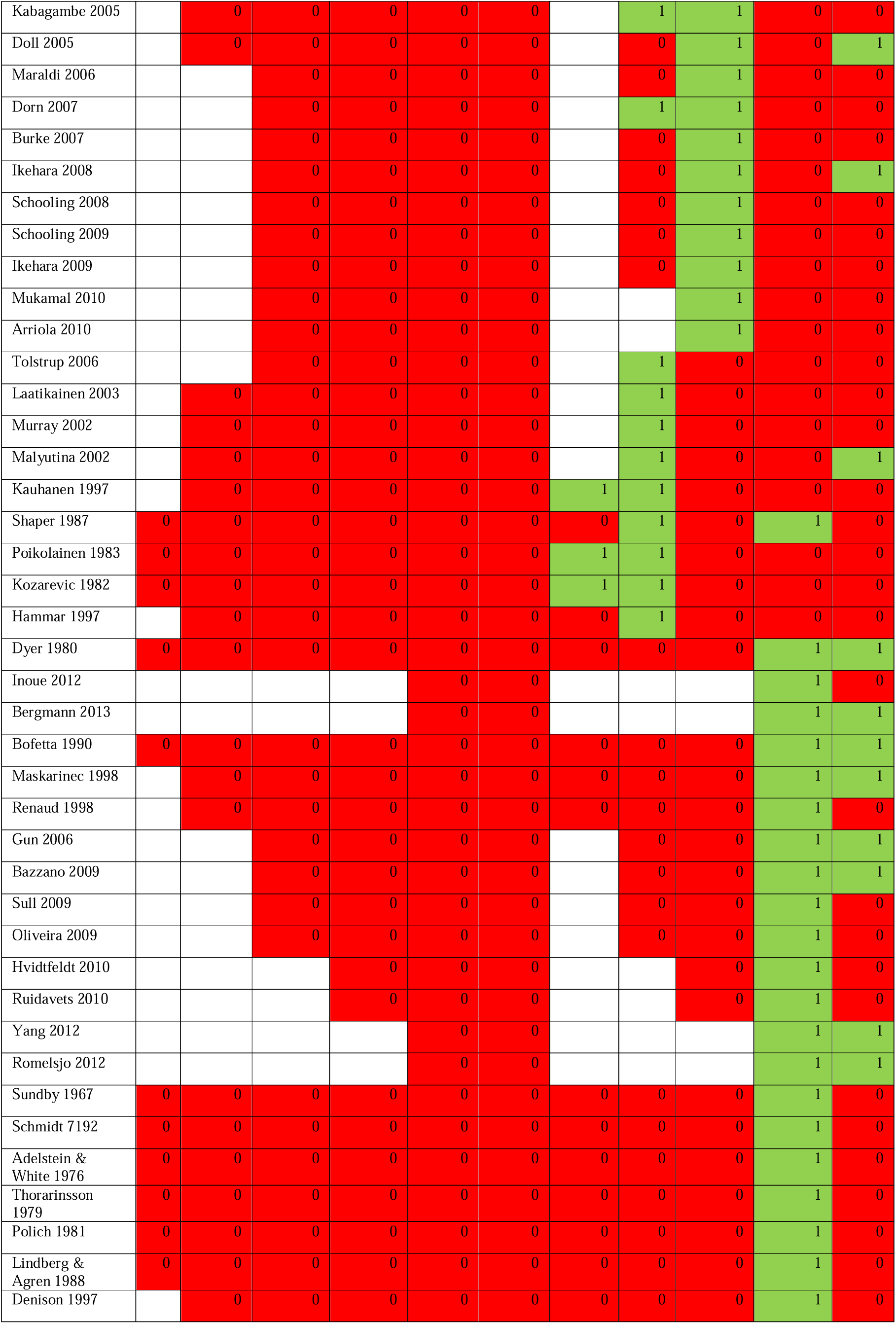
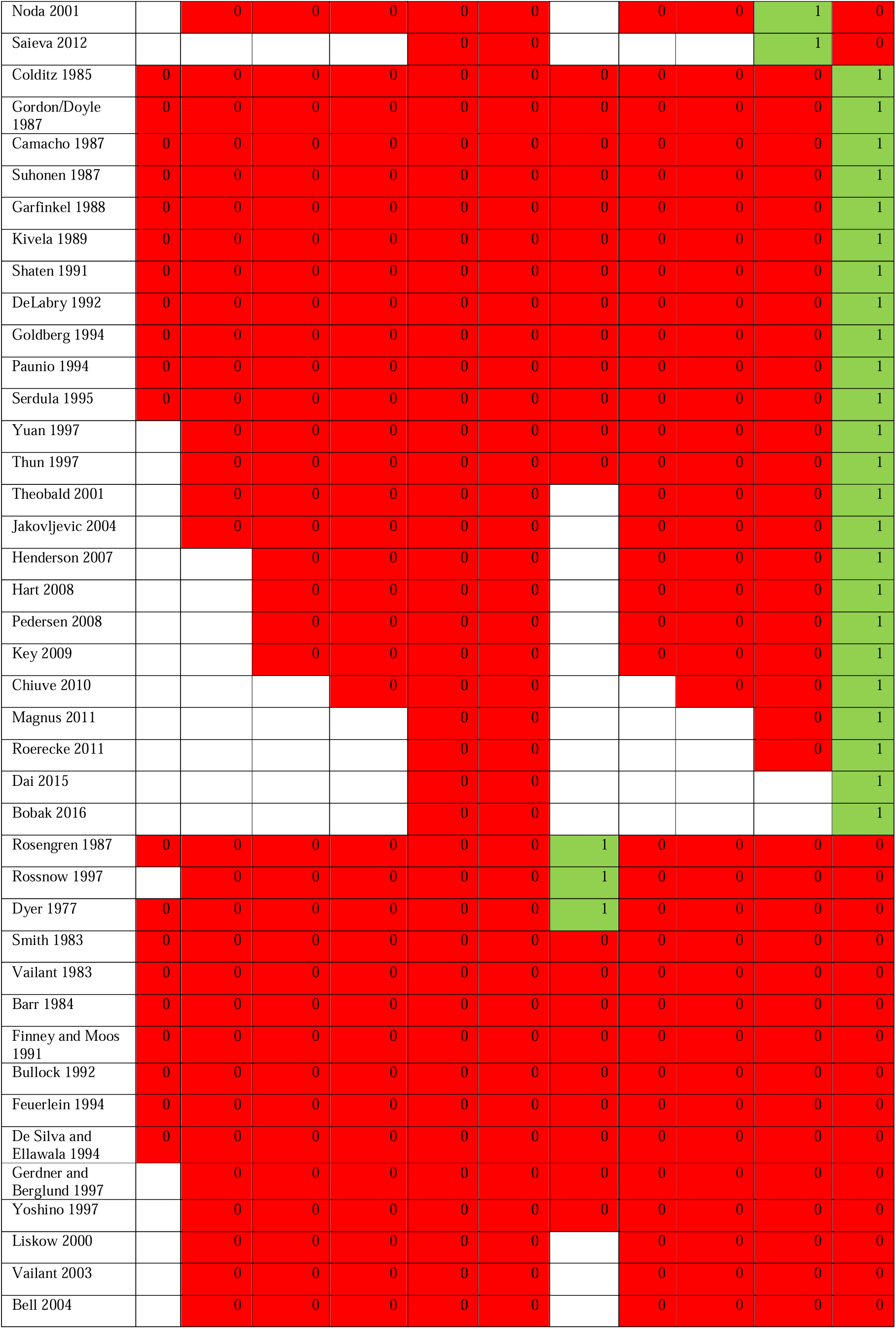
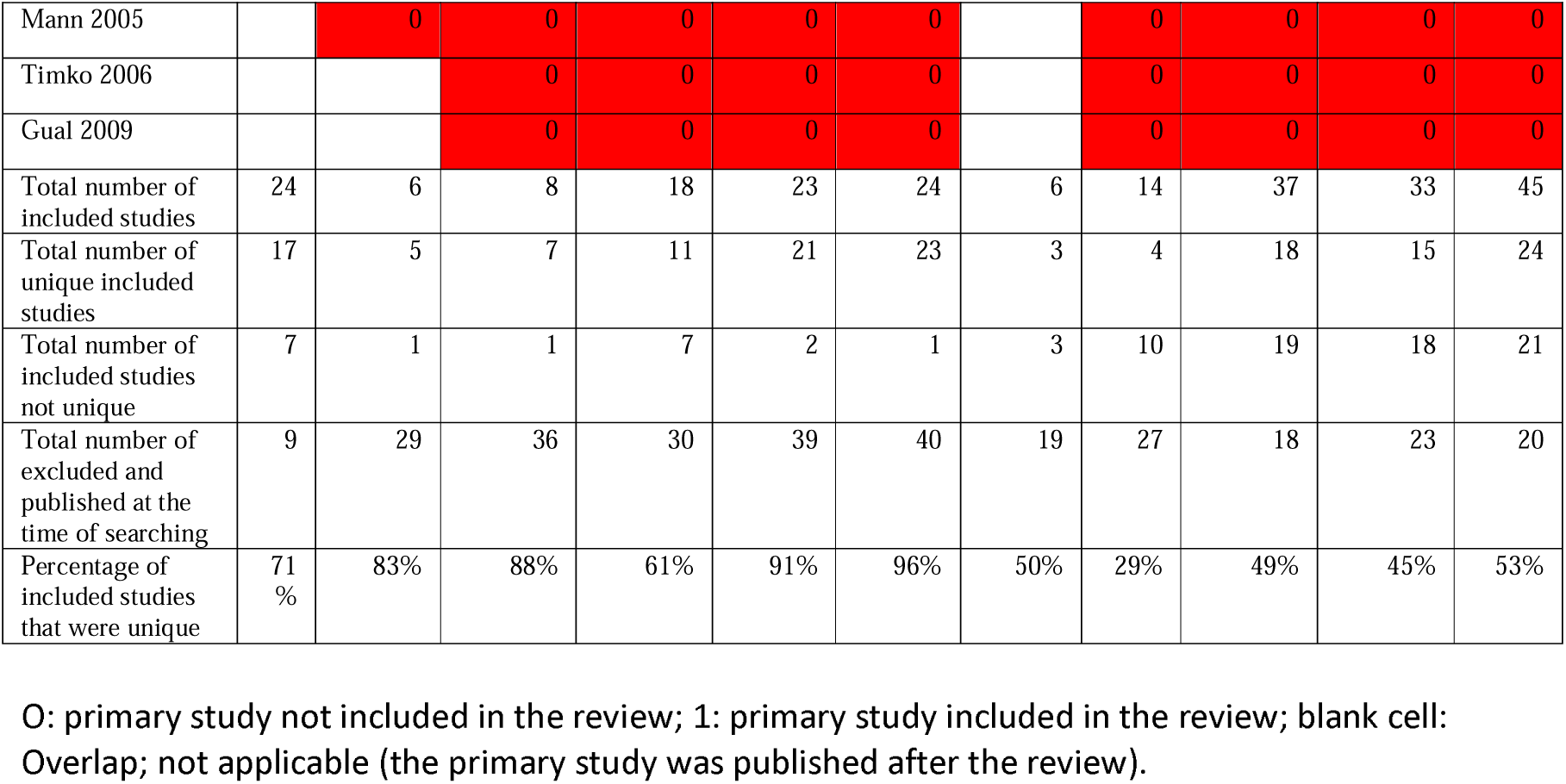
Matrix of included studies in 11 systematic reviews.

**Supplementary Table S4:**
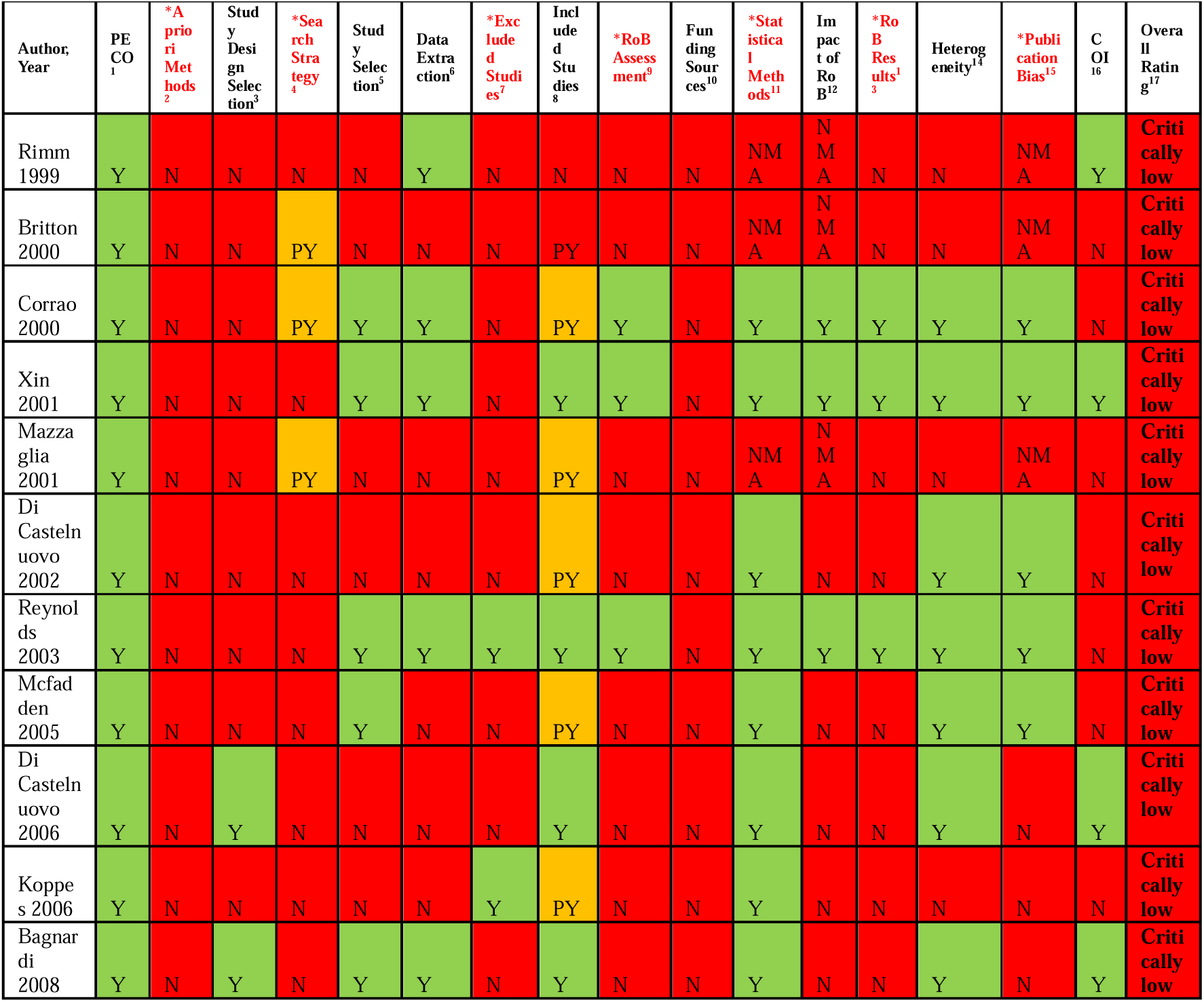
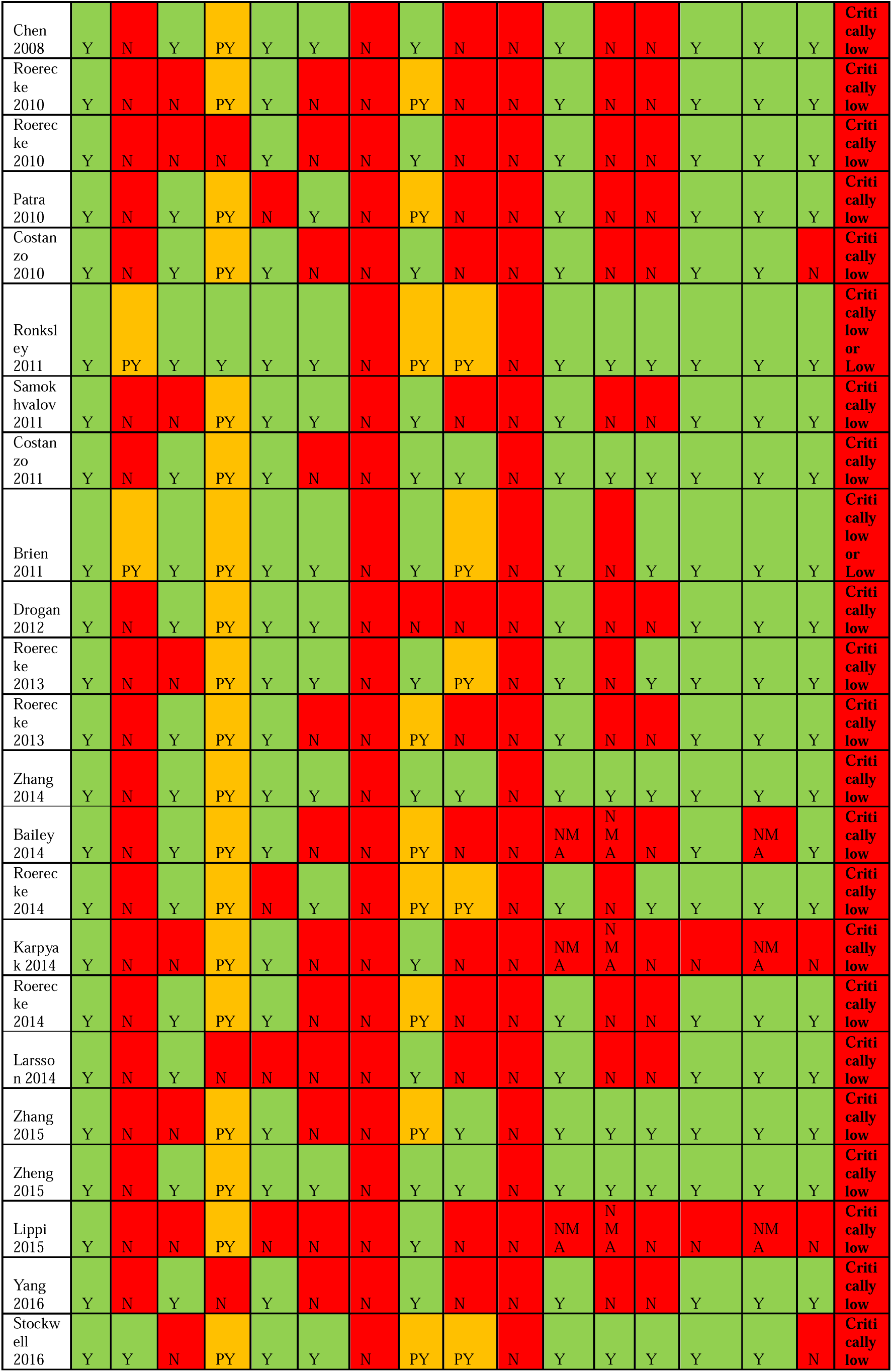
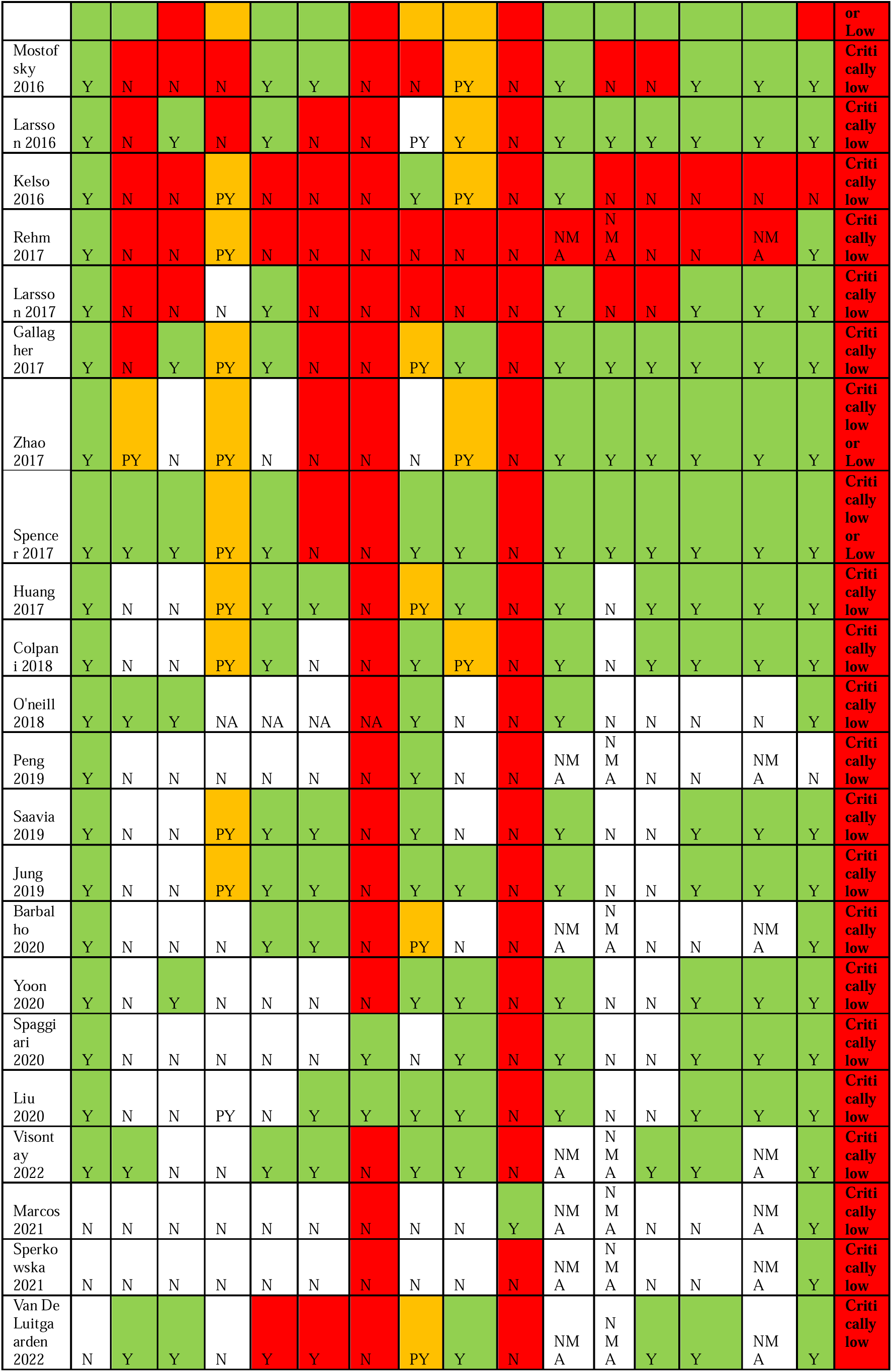
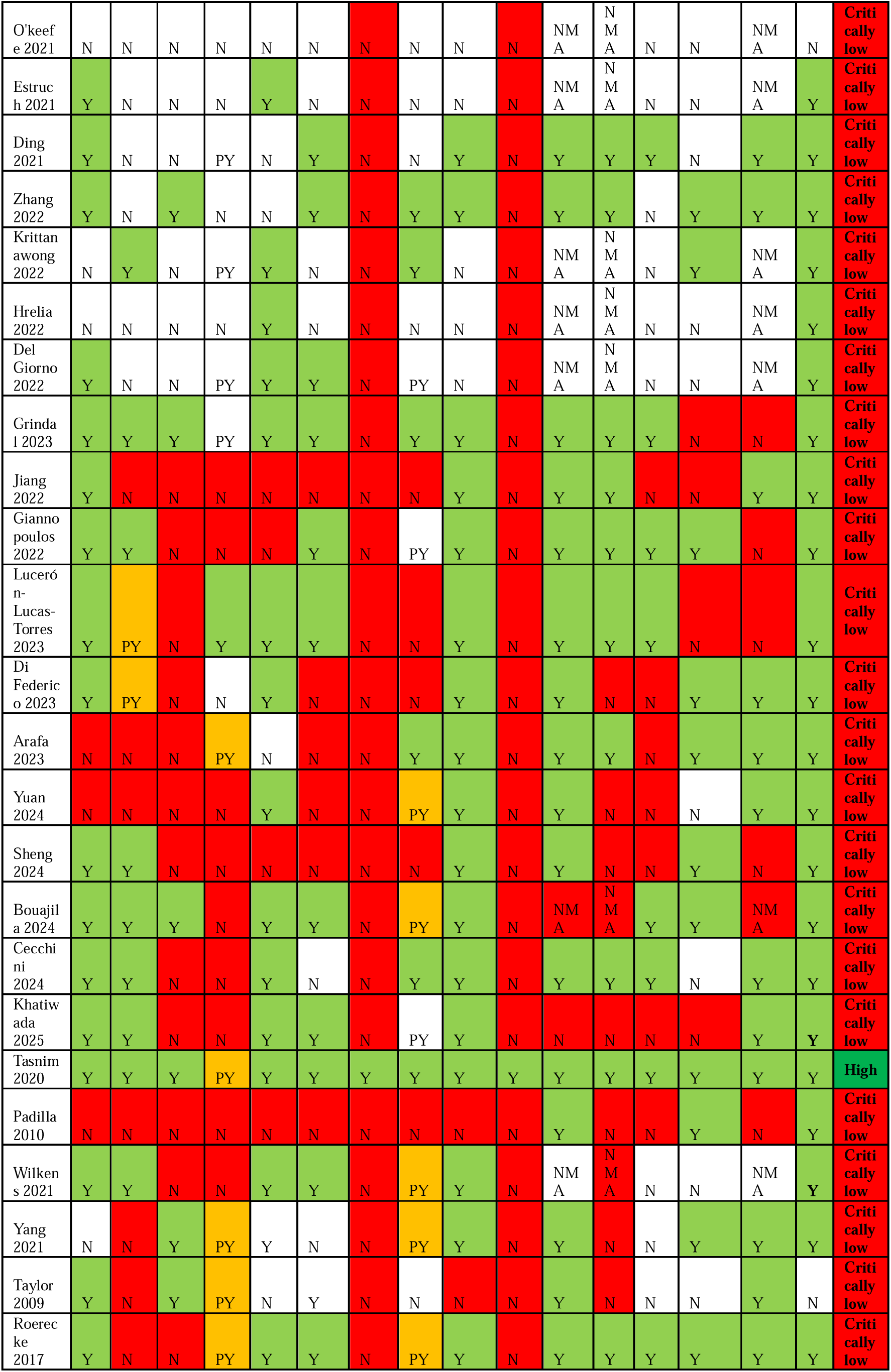
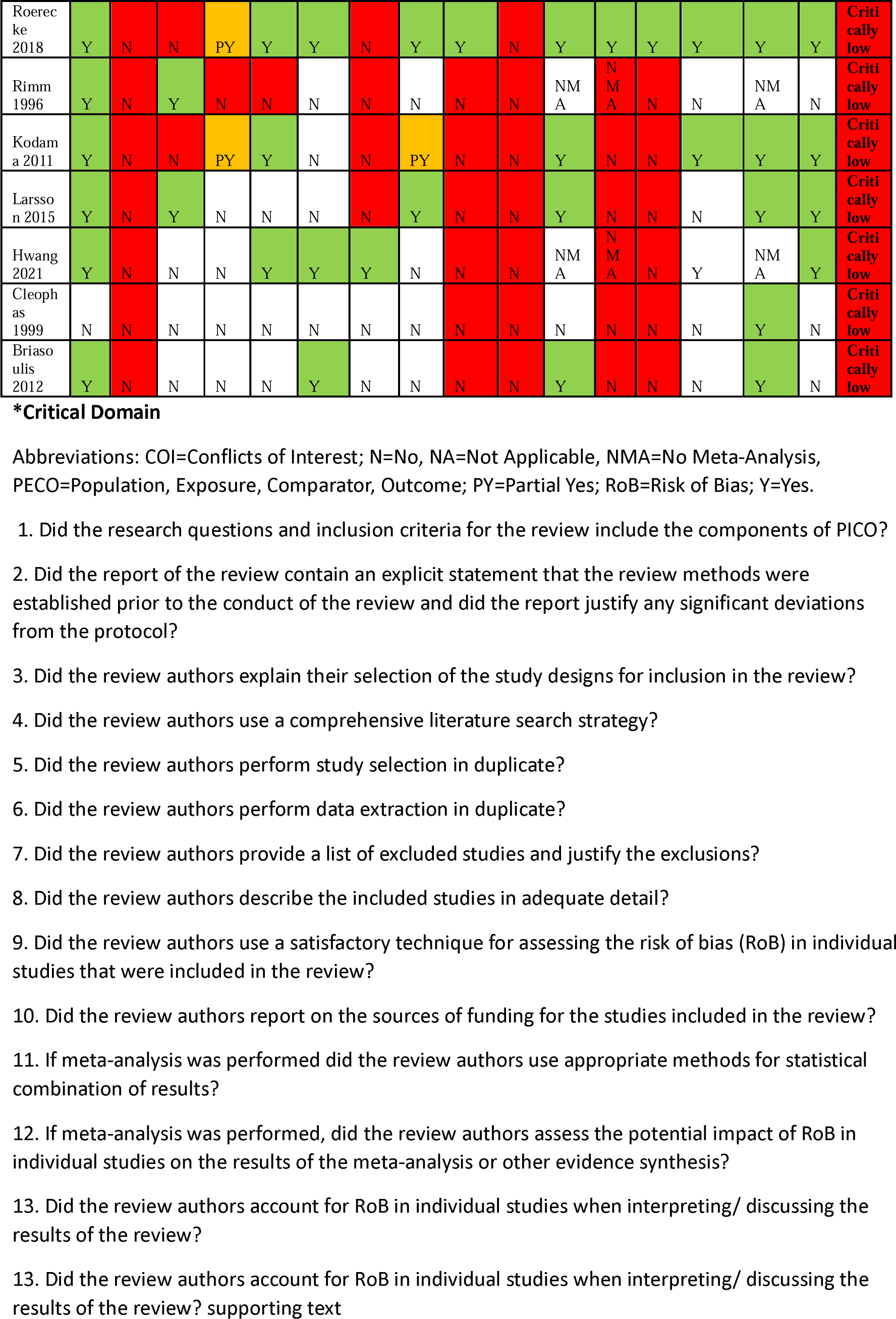
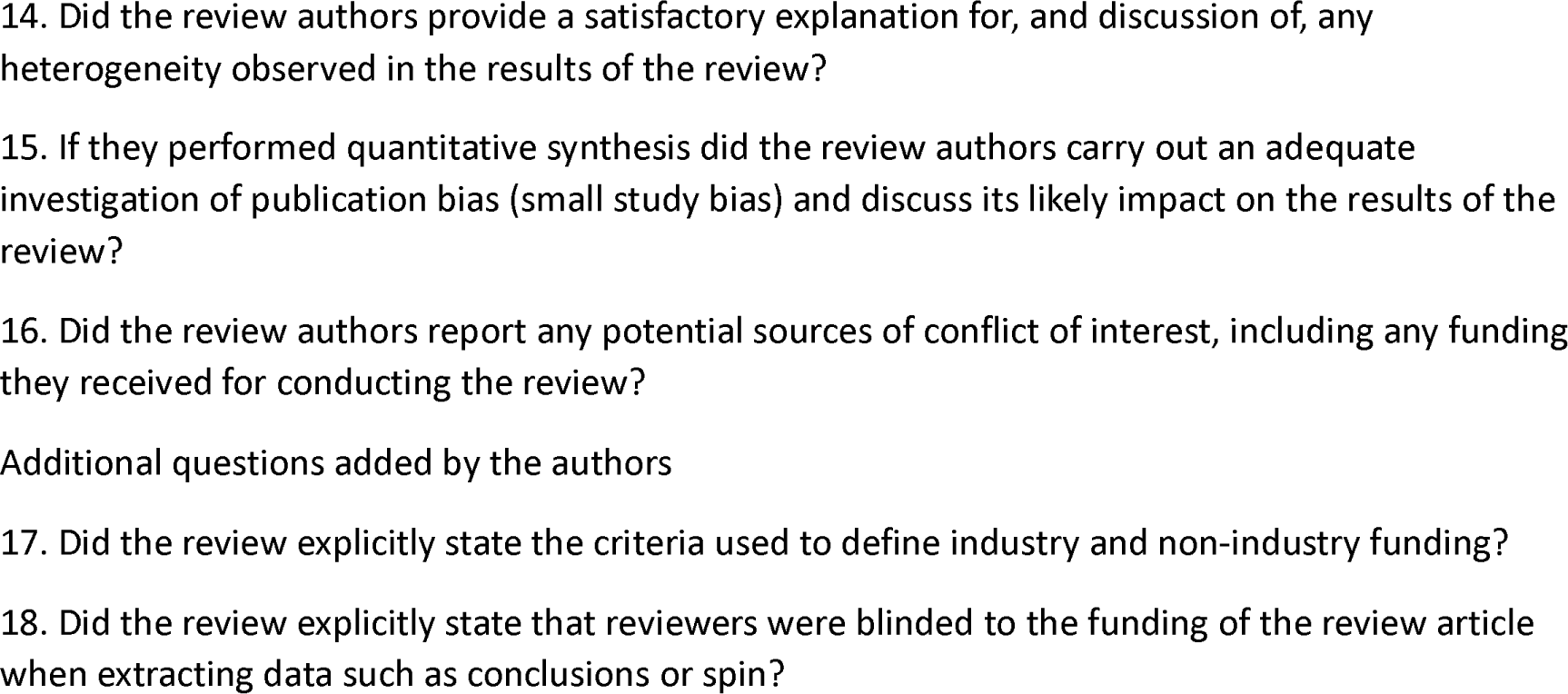
AMSTAR 2 quality checklist ratings for included systematic reviews.

## References

Altman, D. G. (1994). The scandal of poor medical research. Bmj, 308(6924), 283. 10.1136/bmj.308.6924.283

Arafa, A., Kashima, R., Kokubo, Y., Teramoto, M., Sakai, Y., Nosaka, S., &, et al. (2023). Alcohol consumption and the risk of heart failure: the Suita study and meta-analysis of prospective cohort studies. Environ Health Prev Med, 28, 26. 10.1265/ehpm.22-00231

Bagnardi, V., Zatonski, W., Scotti, L., La Vecchia, C., & Corrao, G. (2008). Does drinking pattern modify the effect of alcohol on the risk of coronary heart disease? Evidence from a meta-analysis. Journal of epidemiology and community health, 62(7), 615–619. 10.1136/jech.2007.065607

Barbalho, S. M., Bueno Ottoboni, A. M. M., Fiorini, A. M. R., Guiguer É, L., Nicolau, C. C. T., Goulart, R. A., & Flato, U. A. P. (2020). Grape juice or wine: which is the best option? Critical reviews in food science and nutrition, 1-14. 10.1080/10408398.2019.1710692

Bartlett, A., & McCambridge, J. (2021). Appropriating the Literature: Alcohol Industry Actors’ Interventions in Scientific Journals. J Stud Alcohol Drugs, 82(5), 595–601.

Bartlett, A., & McCambridge, J. (2023). The international center for alcohol policies (ICAP) book series: a key resource globally for alcohol industry political strategies. Subst Abuse Treat Prev Policy, 18(1), 49. 10.1186/s13011-023-00556-9

Bouajila, N., Domenighetti, C., Aubin, H. J., & Naassila, M. (2024). Alcohol consumption and its association with cancer, cardiovascular, liver and brain diseases: a systematic review of Mendelian randomization studies. Front Epidemiol, 4, 1385064. 10.3389/fepid.2024.1385064

Briasoulis, A., Agarwal, V., & Messerli, F. H. (2012). Alcohol consumption and the risk of hypertension in men and women: a systematic review and meta-analysis. J Clin Hypertens (Greenwich), 14(11), 792–798. 10.1111/jch.12008

Brien, S. E., Ronksley, P. E., Turner, B. J., Mukamal, K. J., & Ghali, W. A. (2011). Effect of alcohol consumption on biological markers associated with risk of coronary heart disease: systematic review and meta-analysis of interventional studies. BMJ (Clinical research ed.), 342, d636. 10.1136/bmj.d636

Britton, A., McKee, M., European Centre on Health of Societies in Transition, L. S. o. H., & Tropical Medicine, K. S. L. W. C. E. H. T. (2000). The relation between alcohol and cardiovascular disease in Eastern Europe: explaining the paradox. Journal of Epidemiology & Community Health, 54(5), 328-332. 10.1136/jech.54.5.328

Bryazka, D., Reitsma, M. B., Griswold, M. G., Abate, K. H., Abbafati, C., Abbasi-Kangevari, M., Abbasi-Kangevari, Z., Abdoli, A., Abdollahi, M., Abdullah, A. Y. M., Abhilash, E. S., Abu-Gharbieh, E., Acuna, J. M., Addolorato, G., Adebayo, O. M., Adekanmbi, V., Adhikari, K., Adhikari, S., Adnani, Q. E. S., . . . Gakidou, E. (2022). Population-level risks of alcohol consumption by amount, geography, age, sex, and year: a systematic analysis for the Global Burden of Disease Study 2020. The Lancet, 400(10347), 185–235. 10.1016/S0140-6736(22)00847-9

Cecchini, M., Filippini, T., Whelton, P. K., Iamandii, I., Di Federico, S., Boriani, G., &, et al. (2024). Alcohol intake and risk of hypertension: a systematic review and dose-response meta-analysis of nonexperimental cohort studies. Hypertension, Epub ahead of print. 10.1161/HYPERTENSIONAHA.124.22703

Chen, L., Smith, G. D., Harbord, R. M., & Lewis, S. J. (2008). Alcohol intake and blood pressure: a systematic review implementing a Mendelian randomization approach. PLoS Med, 5(3), e52. 10.1371/journal.pmed.0050052

Clay, J. M., Stockwell, T., Golder, S., Lawrence, K., McCambridge, J., Vishnevsky, N., Zuckermann, A., & Naimi, T. (2025). The International Scientific Forum on Alcohol Research (ISFAR) critiques of alcohol research: Promoting health benefits and downplaying harms. Addiction, 120(11), 2319–2328. 10.1111/add.70132

Cleophas, T. J. (1999). Wine, beer and spirits and the risk of myocardial infarction: a systematic review. Biomedicine & pharmacotherapy = Biomédecine & pharmacothérapie, 53(9), 417–423. 10.1016/S0753-3322(99)80121-8

Closs, S. J., Dowding, D., Allcock, N., Hulme, C., Keady, J., Sampson, E. L., Briggs, M., Corbett, A., Esterhuizen, P., & Holmes, J. (2016). Towards improved decision support in the assessment and management of pain for people with dementia in hospital: a systematic meta-review and observational study.

Colpani, V., Baena, C. P., Jaspers, L., van Dijk, G. M., Farajzadegan, Z., Dhana, K., Tielemans, M. J., Voortman, T., Freak-Poli, R., Veloso, G. G. V., Chowdhury, R., Kavousi, M., Muka, T., & Franco, O. H. (2018). Lifestyle factors, cardiovascular disease and all-cause mortality in middle-aged and elderly women: a systematic review and meta-analysis. European journal of epidemiology, 33(9), 831–845. 10.1007/s10654-018-0374-z

Corrao, G., Rubbiati, L., Bagnardi, V., Zambon, A., & Poikolainen, K. (2000). Alcohol and coronary heart disease: a meta-analysis. Addiction (Abingdon, England), 95(10), 1505–1523. 10.1046/j.1360-0443.2000.951015056.x

Costanzo, S., Di Castelnuovo, A., Donati, M. B., Iacoviello, L., & de Gaetano, G. (2010). Alcohol consumption and mortality in patients with cardiovascular disease: a meta-analysis. Journal of the American College of Cardiology, 55(13), 1339–1347. 10.1016/j.jacc.2010.01.006

Costanzo, S., Di Castelnuovo, A., Donati, M. B., Iacoviello, L., & de Gaetano, G. (2011). Wine, beer or spirit drinking in relation to fatal and non-fatal cardiovascular events: a meta-analysis. European journal of epidemiology, 26(11), 833–850. 10.1007/s10654-011-9631-0

De Santis, K. K., Pieper, D., Lorenz, R. C., Wegewitz, U., Siemens, W., & Matthias, K. (2023). User experience of applying AMSTAR 2 to appraise systematic reviews of healthcare interventions: a commentary. BMC medical research methodology, 23(1), 63. 10.1186/s12874-023-01879-8

Dekkers, O. M., Vandenbroucke, J. P., Cevallos, M., Renehan, A. G., Altman, D. G., & Egger, M. (2019). COSMOS-E: Guidance on conducting systematic reviews and meta-analyses of observational studies of etiology. PLoS Med, 16(2), e1002742. 10.1371/journal.pmed.1002742

Del Giorno, R., Maddalena, A., Bassetti, S., & Gabutti, L. (2022). Association between Alcohol Intake and Arterial Stiffness in Healthy Adults: A Systematic Review. Nutrients, 14(6). 10.3390/nu14061207

Di Castelnuovo, A., Costanzo, S., Bagnardi, V., Donati, M. B., Iacoviello, L., & de Gaetano, G. (2006). Alcohol dosing and total mortality in men and women: an updated meta-analysis of 34 prospective studies. Arch Intern Med, 166(22), 2437–2445. 10.1001/archinte.166.22.2437

Di Castelnuovo, A., Rotondo, S., Iacoviello, L., Donati, M. B., & De Gaetano, G. (2002). Meta-analysis of wine and beer consumption in relation to vascular risk. Circulation, 105(24), 2836–2844. 10.1161/01.CIR.0000018653.19696.01

Di Federico, S., Filippini, T., Whelton, P. K., Cecchini, M., Iamandii, I., Boriani, G., & Vinceti, M. (2023). Alcohol intake and blood pressure levels: a dose-response meta-analysis of nonexperimental cohort studies. Hypertension, Epub ahead of print. 10.1161/HYPERTENSIONAHA.123.21224

Ding, C., O’Neill, D., Bell, S., Stamatakis, E., & Britton, A. (2021). Association of alcohol consumption with morbidity and mortality in patients with cardiovascular disease: original data and meta-analysis of 48,423 men and women. BMC medicine, 19(1), 167. 10.1186/s12916-021-02040-2

Drogan, D., Sheldrick, A. J., Schütze, M., Knüppel, S., Andersohn, F., di Giuseppe, R., Herrmann, B., Willich, S. N., Garbe, E., Bergmann, M. M., Boeing, H., & Weikert, C. (2012). Alcohol consumption, genetic variants in alcohol deydrogenases, and risk of cardiovascular diseases: a prospective study and meta-analysis. PLoS ONE, 7(2), e32176. 10.1371/journal.pone.0032176

Estruch, R., & Hendriks, H. F. J. (2021). Associations between Low to Moderate Consumption of Alcoholic Beverage Types and Health Outcomes: A Systematic Review. Alcohol and alcoholism (Oxford, Oxfordshire). 10.1093/alcalc/agab082

Fuchs, F. D., & Fuchs, S. C. (2021). The Effect of Alcohol on Blood Pressure and Hypertension. Current hypertension reports, 23(10), 42. 10.1007/s11906-021-01160-7

Gallagher, C., Hendriks, J. M. L., Elliott, A. D., Wong, C. X., Rangnekar, G., Middeldorp, M. E., Mahajan, R., Lau, D. H., & Sanders, P. (2017). Alcohol and incident atrial fibrillation - A systematic review and meta-analysis. International journal of cardiology, 246, 46–52. 10.1016/j.ijcard.2017.05.133

Giannopoulos, G., Anagnostopoulos, I., Kousta, M., Vergopoulos, S., Deftereos, S., & Vassilikos, V. (2022). Alcohol Consumption and the Risk of Incident Atrial Fibrillation: A Meta-Analysis. Diagnostics (Basel, Switzerland), 12(2). 10.3390/diagnostics12020479

Golder, S., & McCambridge, J. (2021). Alcohol, cardiovascular disease and industry funding: A co-authorship network analysis of systematic reviews. Soc Sci Med, 289, 114450. 10.1016/j.socscimed.2021.114450

Green, B., Bailey, M., Bridge, K., & Scott, J. (2014). Alcohol consumption as a risk factor for abdominal aortic aneurysm. J Vasc Med Surg, 2(140), 2.

Grindal, A. W., Sparrow, R. T., McIntyre, W. F., Conen, D., Healey, J. S., & Wong, J. A. (2022). Alcohol Consumption and Atrial Arrhythmia Recurrence After Atrial Fibrillation Ablation: A Systematic Review and Meta-analysis. The Canadian journal of cardiology. 10.1016/j.cjca.2022.12.010

Hennessy, E. A., & Johnson, B. T. (2020). Examining overlap of included studies in meta-reviews: Guidance for using the corrected covered area index. Res Synth Methods, 11(1), 134–145. 10.1002/jrsm.1390

Higgins JPT, T. J., Chandler J, Cumpston M, Li T, Page MJ, Welch VA (editors). . (2024). Cochrane Handbook for Systematic Reviews of Interventions version 6.5 (updated August 2024). Cochrane, 2024. Available from www.cochrane.org/handbook.

Hrelia, S., Di Renzo, L., Bavaresco, L., Bernardi, E., Malaguti, M., & Giacosa, A. (2022). Moderate Wine Consumption and Health: A Narrative Review. Nutrients, 15(1). 10.3390/nu15010175

Huang, C., Zhan, J., Liu, Y. J., Li, D. J., Wang, S. Q., & He, Q. Q. (2014). Association between alcohol consumption and risk of cardiovascular disease and all-cause mortality in patients with hypertension: a meta-analysis of prospective cohort studies. Mayo Clinic proceedings, 89(9), 1201–1210. 10.1016/j.mayocp.2014.05.014

Huang, Y., Li, Y., Zheng, S., Yang, X., Wang, T., & Zeng, J. (2017). Moderate alcohol consumption and atherosclerosis: Meta-analysis of effects on lipids and inflammation. Wiener Klinische Wochenschrift, 129(21-22), 1–9. 10.1007/s00508-017-1235-6

Hwang, C. L., Piano, M. R., & Phillips, S. A. (2021). The effects of alcohol consumption on flow-mediated dilation in humans: A systematic review. Physiological reports, 9(10), e14872. 10.14814/phy2.14872

Jiang, H., Mei, X., Jiang, Y., Yao, J., Shen, J., Chen, T., & Zhou, Y. (2022). Alcohol consumption and atrial fibrillation risk: An updated dose-response meta-analysis of over 10 million participants. Frontiers in cardiovascular medicine, 9, 979982. 10.3389/fcvm.2022.979982

Jung, M. H., Shin, E. S., Ihm, S. H., Jung, J. G., Lee, H. Y., & Kim, C. H. (2020). The effect of alcohol dose on the development of hypertension in Asian and Western men: systematic review and meta-analysis. Korean J Intern Med, 35(4), 906–916. 10.3904/kjim.2019.016

Karpyak, V. M., Romanowicz, M., Schmidt, J. E., Lewis, K. A., & Bostwick, J. M. (2014). Characteristics of heart rate variability in alcohol-dependent subjects and nondependent chronic alcohol users. Alcoholism, clinical and experimental research, 38(1), 9–26. 10.1111/acer.12270

Kelso, N. E., Sheps, D. S., & Cook, R. L. (2015). The association between alcohol use and cardiovascular disease among people living with HIV: a systematic review. The American journal of drug and alcohol abuse, 41(6), 479–488. 10.3109/00952990.2015.1058812

Khatiwada, A., Christensen, S. H., Rawal, A., Dragsted, L. O., Berg-Beckhoff, G., & Wilkens, T. L. (2025). Effect of moderate alcohol intake on blood apolipoproteins concentrations: a meta-analysis of human intervention studies. Nutr Metab Cardiovasc Dis, Epub ahead of print, 103854. 10.1016/j.numecd.2025.103854

Kodama, S., Saito, K., Tanaka, S., Horikawa, C., Saito, A., Heianza, Y., Anasako, Y., Nishigaki, Y., Yachi, Y., Iida, K. T., Ohashi, Y., Yamada, N., & Sone, H. (2011). Alcohol consumption and risk of atrial fibrillation: a meta-analysis. Journal of the American College of Cardiology, 57(4), 427–436. 10.1016/j.jacc.2010.08.641

Koppes, L. L., Dekker, J. M., Hendriks, H. F., Bouter, L. M., & Heine, R. J. (2006). Meta-analysis of the relationship between alcohol consumption and coronary heart disease and mortality in type 2 diabetic patients. Diabetologia, 49(4), 648–652. 10.1007/s00125-005-0127-x

Krittanawong, C., Isath, A., Rosenson, R. S., Khawaja, M., Wang, Z., Fogg, S. E., Virani, S. S., Qi, L., Cao, Y., Long, M. T., Tangney, C. C., & Lavie, C. J. (2022). Alcohol Consumption and Cardiovascular Health. The American journal of medicine. 10.1016/j.amjmed.2022.04.021

Larsson, S. C., Drca, N., & Wolk, A. (2014). Alcohol consumption and risk of atrial fibrillation: a prospective study and dose-response meta-analysis. Journal of the American College of Cardiology, 64(3), 281–289. 10.1016/j.jacc.2014.03.048

Larsson, S. C., Orsini, N., & Wolk, A. (2015). Alcohol consumption and risk of heart failure: A dose-response meta-analysis of prospective studies. European Journal of Heart Failure, 17(4), 367–373. 10.1002/ejhf.228

Larsson, S. C., Wallin, A., & Wolk, A. (2018). Alcohol consumption and risk of heart failure: Meta-analysis of 13 prospective studies. Clinical Nutrition, 37(4), 1247–1251. 10.1016/j.clnu.2017.05.007

Larsson, S. C., Wallin, A., Wolk, A., & Markus, H. S. (2016). Differing association of alcohol consumption with different stroke types: a systematic review and meta-analysis. BMC medicine, 14(1), 178. 10.1186/s12916-016-0721-4

Lippi, G., Mattiuzzi, C., & Franchini, M. (2015). Alcohol consumption and venous thromboembolism: friend or foe? Internal and emergency medicine, 10(8), 907–913. 10.1007/s11739-015-1327-0

Litten, R., & Gardner, M. (2022). The basics: defining how much alcohol is too much. National Institute on Alcohol Abuse and Alcoholism (NIAAA). https://www.niaaa.nih.gov/health-professionals-communities/core-resource-on-alcohol/basics-defining-how-much-alcohol-too-much

Liu, F., Zhao, Y., Wang, B., Ren, Y., Liu, X., Zhang, D., Chen, X., Cheng, C., Liu, L., Liu, D., Zhang, M., Hu, D., Liu, F., Liu, Y., Sun, X., Yin, Z., Li, H., Zhao, Y., Wang, B., . . . Hong, S. (2020). Race- and sex-specific association between alcohol consumption and hypertension in 22 cohort studies: A systematic review and meta-analysis. Nutr. Metab. Cardiovasc. Dis., 30(8), 1249–1259. 10.1016/j.numecd.2020.03.018

Luceron-Lucas-Torres, M., Saz-Lara, A., Diez-Fernandez, A., Martinez-Garcia, I., Martinez-Vizcaino, V., Cavero-Redondo, I., &, et al. (2023). Association between wine consumption with cardiovascular disease and cardiovascular mortality: a systematic review and meta-analysis. Nutrients, Epub ahead of print. 10.3390/nu15122785

Marcos, A., Serra-Majem, L., Pérez-Jiménez, F., Pascual, V., Tinahones, F. J., & Estruch, R. (2021). Moderate Consumption of Beer and Its Effects on Cardiovascular and Metabolic Health: An Updated Review of Recent Scientific Evidence. Nutrients, 13(3). 10.3390/nu13030879

Mazzaglia, G., Britton, A. R., Altmann, D. R., & Chenet, L. (2001). Exploring the relationship between alcohol consumption and non-fatal or fatal stroke: a systematic review. Addiction, 96(12), 1743–1756. 10.1046/j.1360-0443.2001.961217434.x

McCambridge, J., & Golder, S. (2024). Alcohol, cardiovascular disease and industry funding: A co-authorship network analysis of epidemiological studies. Addictive Behaviors, 151, 107932. 10.1016/j.addbeh.2023.107932

McCambridge, J., & Mialon, M. (2018). Alcohol industry involvement in science: A systematic review of the perspectives of the alcohol research community. Drug Alcohol Rev, 37(5), 565–579. 10.1111/dar.12826

McCambridge, J., Mitchell, G., Lesch, M., Filippou, A., Golder, S., Garry, J., Bartlett, A., & Madden, M. (2023). The emperor has no clothes: a synthesis of findings from the Transformative Research on the Alcohol industry, Policy and Science research programme. Addiction, 118(3), 558–566. 10.1111/add.16058

McFadden, C. B., Brensinger, C. M., Berlin, J. A., & Townsend, R. R. (2005). Systematic review of the effect of daily alcohol intake on blood pressure. Am J Hypertens, 18(2 Pt 1), 276-286. 10.1016/j.amjhyper.2004.07.020

Mitchell, G., Lesch, M., & McCambridge, J. (2020). Alcohol Industry Involvement in the Moderate Alcohol and Cardiovascular Health Trial. Am J Public Health, 110(4), 485–488. 10.2105/ajph.2019.305508

Mostofsky, E., Chahal, H. S., Mukamal, K. J., Rimm, E. B., & Mittleman, M. A. (2016). Alcohol and Immediate Risk of Cardiovascular Events: A Systematic Review and Dose-Response Meta-Analysis. Circulation, 133(10), 979–987. 10.1161/CIRCULATIONAHA.115.019743

Naame, S. A., Li, D., & Huang, R. (2019). Effects of moderate red wine on cardiovascular risk factors in diabetics: A systematic review and meta-analysis of randomized controlled trials. Toxicology Research, 8(6), 979–987. 10.1039/c9tx00227h

Naimi, T. S., Stockwell, T., Zhao, J., Xuan, Z., Dangardt, F., Saitz, R., Liang, W., & Chikritzhs, T. (2017). Selection biases in observational studies affect associations between ‘moderate’ alcohol consumption and mortality. Addiction, 112(2), 207–214. 10.1111/add.13451

Naimi, T. S., Xuan, Z., Brown, D. W., & Saitz, R. (2013). Confounding and studies of ‘moderate’ alcohol consumption: the case of drinking frequency and implications for low-risk drinking guidelines. Addiction, 108(9), 1534–1543. 10.1111/j.1360-0443.2012.04074.x

O’Keefe, E. L., Sturgess, J. E., O’Keefe, J. H., Gupta, S., & Lavie, C. J. (2021). Prevention and Treatment of Atrial Fibrillation via Risk Factor Modification. The American journal of cardiology. 10.1016/j.amjcard.2021.08.042

O’Neill, D., Britton, A., Hannah, M. K., Goldberg, M., Kuh, D., Khaw, K. T., & Bell, S. (2018). Association of longitudinal alcohol consumption trajectories with coronary heart disease: a meta-analysis of six cohort studies using individual participant data. BMC medicine, 16(1), 124. 10.1186/s12916-018-1123-6

Okojie, O. M., Javed, F., Chiwome, L., & Hamid, P. (2020). Hypertension and Alcohol: A Mechanistic Approach. Cureus, 12(8), e10086. 10.7759/cureus.10086

Oppenheimer, G. M., & Bayer, R. (2020). Is Moderate Drinking Protective Against Heart Disease? The Science, Politics and History of a Public Health Conundrum. Milbank Q, 98(1), 39–56. 10.1111/1468-0009.12437

Padilla, H., Michael Gaziano, J., & Djoussé, L. (2010). Alcohol consumption and risk of heart failure: a meta-analysis. The Physician and sportsmedicine, 38(3), 84–89. 10.3810/psm.2010.10.1812

Patra, J., Taylor, B., Irving, H., Roerecke, M., Baliunas, D., Mohapatra, S., & Rehm, J. (2010). Alcohol consumption and the risk of morbidity and mortality for different stroke types--a systematic review and meta-analysis. BMC Public Health, 10, 258. 10.1186/1471-2458-10-258

Peake, L., van Schalkwyk, M. C. I., Maani, N., & Petticrew, M. (2021). Analysis of the accuracy and completeness of cardiovascular health information on alcohol industry-funded websites. Eur J Public Health, 31(6), 1197–1204. 10.1093/eurpub/ckab135

Peng, J., Wang, H., Rong, X., He, L., Xiangpen, L., Shen, Q., & Peng, Y. (2020). Cerebral Hemorrhage and Alcohol Exposure: A Review. Alcohol Alcohol, 55(1), 20–27. 10.1093/alcalc/agz087

Qureshi, R., Mayo-Wilson, E., Rittiphairoj, T., McAdams-DeMarco, M., Guallar, E., & Li, T. (2022). Harms in Systematic Reviews Paper 3: Given the same data sources, systematic reviews of gabapentin have different results for harms. J Clin Epidemiol, 143, 224–241. 10.1016/j.jclinepi.2021.10.025

Qureshi, R., Naaman, K., Quan, N. G., Mayo-Wilson, E., Page, M. J., Cornelius, V., Chou, R., Boutron, I., Golder, S., Bero, L., Doshi, P., Vassar, M., Meursinge Reynders, R., & Li, T. (2024). Development and Evaluation of a Framework for Identifying and Addressing Spin for Harms in Systematic Reviews of Interventions. Ann Intern Med, 177(8), 1089–1098. 10.7326/m24-0771

Rehm, J., Hasan, O. S. M., Imtiaz, S., & Neufeld, M. (2017). Quantifying the contribution of alcohol to cardiomyopathy: A systematic review. Alcohol (Fayetteville, N.Y.), 61, 9–15. 10.1016/j.alcohol.2017.01.011

Reynolds, K., Lewis, B., Nolen, J. D., Kinney, G. L., Sathya, B., & He, J. (2003). Alcohol consumption and risk of stroke: a meta-analysis. Jama, 289(5), 579–588. 10.1001/jama.289.5.579

Rimm, E. B., Klatsky, A., Grobbee, D., & Stampfer, M. J. (1996). Review of moderate alcohol consumption and reduced risk of coronary heart disease: is the effect due to beer, wine, or spirits. Bmj, 312(7033), 731–736. 10.1136/bmj.312.7033.731

Rimm, E. B., Williams, P., Fosher, K., Criqui, M., & Stampfer, M. J. (1999). Moderate alcohol intake and lower risk of coronary heart disease: meta-analysis of effects on lipids and haemostatic factors. BMJ (Clinical research ed.), 319(7224), 1523–1528. 10.1136/bmj.319.7224.1523

Roerecke, M., Gual, A., & Rehm, J. (2013). Reduction of alcohol consumption and subsequent mortality in alcohol use disorders: systematic review and meta-analyses. J Clin Psychiatry, 74(12), e1181–1189. 10.4088/JCP.13r08379

Roerecke, M., Kaczorowski, J., Tobe, S. W., Gmel, G., Hasan, O. S. M., & Rehm, J. (2017). The effect of a reduction in alcohol consumption on blood pressure: a systematic review and meta-analysis. The Lancet. Public health, 2(2), e108–e120. 10.1016/S2468-2667(17)30003-8

Roerecke, M., & Rehm, J. (2010). Irregular heavy drinking occasions and risk of ischemic heart disease: a systematic review and meta-analysis. American journal of epidemiology, 171(6), 633–644. 10.1093/aje/kwp451

Roerecke, M., & Rehm, J. (2011). Ischemic heart disease mortality and morbidity rates in former drinkers: a meta-analysis. American journal of epidemiology, 173(3), 245–258. 10.1093/aje/kwq364

Roerecke, M., & Rehm, J. (2012). The cardioprotective association of average alcohol consumption and ischaemic heart disease: a systematic review and meta-analysis. Addiction (Abingdon, England), 107(7), 1246–1260. 10.1111/j.1360-0443.2012.03780.x

Roerecke, M., & Rehm, J. (2014a). Alcohol consumption, drinking patterns, and ischemic heart disease: a narrative review of meta-analyses and a systematic review and meta-analysis of the impact of heavy drinking occasions on risk for moderate drinkers. BMC medicine, 12(1), 182. 10.1186/s12916-014-0182-6

Roerecke, M., & Rehm, J. (2014b). Chronic heavy drinking and ischaemic heart disease: a systematic review and meta-analysis. Open Heart, 1(1), e000135. 10.1136/openhrt-2014-000135

Roerecke, M., Tobe, S. W., Kaczorowski, J., Bacon, S. L., Vafaei, A., Hasan, O. S. M., Krishnan, R. J., Raifu, A. O., & Rehm, J. (2018). Sex-Specific Associations Between Alcohol Consumption and Incidence of Hypertension: A Systematic Review and Meta-Analysis of Cohort Studies. Journal of the American Heart Association, 7(13), 1–27. 10.1161/JAHA.117.008202

Ronksley, P. E., Brien, S. E., Turner, B. J., Mukamal, K. J., & Ghali, W. A. (2011). Association of alcohol consumption with selected cardiovascular disease outcomes: a systematic review and meta-analysis. BMJ (Clinical research ed.), 342, d671. 10.1136/bmj.d671

Samokhvalov, A. V., Irving, H. M., & Rehm, J. (2010). Alcohol consumption as a risk factor for atrial fibrillation: a systematic review and meta-analysis. European journal of cardiovascular prevention and rehabilitation : official journal of the European Society of Cardiology, Working Groups on Epidemiology & Prevention and Cardiac Rehabilitation and Exercise Physiology, 17(6), 706–712. 10.1097/HJR.0b013e32833a1947

Shea, B. J., Reeves, B. C., Wells, G., Thuku, M., Hamel, C., Moran, J., Moher, D., Tugwell, P., Welch, V., & Kristjansson, E. (2017). AMSTAR 2: a critical appraisal tool for systematic reviews that include randomised or non-randomised studies of healthcare interventions, or both. Bmj, 358.

Sheng, Y., Meng, G., Li, G., & Wang, J. (2024). Red wine alleviates atherosclerosis-related inflammatory markers in healthy subjects rather than in high cardiovascular risk subjects: a systematic review and meta-analysis. Medicine, 103(23), e38229. 10.1097/MD.0000000000038229

Spaggiari, G., Cignarelli, A., Sansone, A., Baldi, M., & Santi, D. (2020). To beer or not to beer: A meta-analysis of the effects of beer consumption on cardiovascular health. PLoS ONE, 15(6), e0233619. 10.1371/journal.pone.0233619

Spencer, S. M., Trower, A. J., Jia, X., Scott, D. J. A., & Greenwood, D. C. (2017). Meta-analysis of the association between alcohol consumption and abdominal aortic aneurysm. The British journal of surgery, 104(13), 1756–1764. 10.1002/bjs.10674

Sperkowska, B., Murawska, J., Przybylska, A., Gackowski, M., Kruszewski, S., Durmowicz, M., & Rutkowska, D. (2021). Cardiovascular Effects of Chocolate and Wine-Narrative Review. Nutrients, 13(12). 10.3390/nu13124269

Stockwell, T., Zhao, J., & Macdonald, S. (2014). Who under-reports their alcohol consumption in telephone surveys and by how much? An application of the ‘yesterday method’ in a national Canadian substance use survey. Addiction, 109(10), 1657–1666. 10.1111/add.12609

Stockwell, T., Zhao, J., Panwar, S., Roemer, A., Naimi, T., & Chikritzhs, T. (2016). Do "Moderate" Drinkers Have Reduced Mortality Risk? A Systematic Review and Meta-Analysis of Alcohol Consumption and All-Cause Mortality. J Stud Alcohol Drugs, 77(2), 185–198. 10.15288/jsad.2016.77.185

Tasnim, S., Tang, C., Musini, V. M., & Wright, J. M. (2020). Effect of alcohol on blood pressure. The Cochrane database of systematic reviews, 7, CD012787. 10.1002/14651858.CD012787.pub2

Taylor, B., Irving, H. M., Baliunas, D., Roerecke, M., Patra, J., Mohapatra, S., & Rehm, J. (2009). Alcohol and hypertension: gender differences in dose-response relationships determined through systematic review and meta-analysis. Addiction, 104(12), 1981–1990. 10.1111/j.1360-0443.2009.02694.x

van de Luitgaarden, I. A. T., van Oort, S., Bouman, E. J., Schoonmade, L. J., Schrieks, I. C., Grobbee, D. E., van der Schouw, Y. T., Larsson, S. C., Burgess, S., van Ballegooijen, A. J., Onland-Moret, N. C., & Beulens, J. W. J. (2021). Alcohol consumption in relation to cardiovascular diseases and mortality: a systematic review of Mendelian randomization studies. European journal of epidemiology. 10.1007/s10654-021-00799-5

Visontay, R., Sunderland, M., Slade, T., Wilson, J., & Mewton, L. (2022). Are there non-linear relationships between alcohol consumption and long-term health?: a systematic review of observational studies employing approaches to improve causal inference. BMC medical research methodology, 22(1), 16. 10.1186/s12874-021-01486-5

Weaver, S. R., Rendeiro, C., McGettrick, H. M., Philp, A., & Lucas, S. J. E. (2020). Fine wine or sour grapes? A systematic review and meta-analysis of the impact of red wine polyphenols on vascular health. European journal of nutrition, 60(1). 10.1007/s00394-020-02247-8

Whitman, I. R., Pletcher, M. J., Vittinghoff, E., Imburgia, K. E., Maguire, C., Bettencourt, L., Sinha, T., Parsnick, T., Tison, G. H., Mulvanny, C. G., Olgin, J. E., & Marcus, G. M. (2015). Perceptions, Information Sources, and Behavior Regarding Alcohol and Heart Health. Am J Cardiol, 116(4), 642–646. 10.1016/j.amjcard.2015.05.029

Wilkens, T. L., Tranæs, K., Eriksen, J. N., & Dragsted, L. O. (2021). Moderate alcohol consumption and lipoprotein subfractions: a systematic review of intervention and observational studies. Nutrition reviews. 10.1093/nutrit/nuab102

Xin, X., He, J., Frontini, M. G., Ogden, L. G., Motsamai, O. I., & Whelton, P. K. (2001). Effects of alcohol reduction on blood pressure: a meta-analysis of randomized controlled trials. Hypertension, 38(5), 1112–1117. 10.1161/hy1101.093424

Yang, L., Chen, H., Shu, T., Pan, M., & Huang, W. (2021). Risk of incident atrial fibrillation with low-to-moderate alcohol consumption is associated with gender, region, alcohol category: a systematic review and meta-analysis. Europace : European pacing, arrhythmias, and cardiac electrophysiology : journal of the working groups on cardiac pacing, arrhythmias, and cardiac cellular electrophysiology of the European Society of Cardiology. 10.1093/europace/euab266

Yang, Y., Liu, D. C., Wang, Q. M., Long, Q. Q., Zhao, S., Zhang, Z., Ma, Y., Wang, Z. M., Chen, L. L., & Wang, L. S. (2016). Alcohol consumption and risk of coronary artery disease: A dose-response meta-analysis of prospective studies. Nutrition, 32(6), 637–644. 10.1016/j.nut.2015.11.013

Ye, J., Chen, X., & Bao, L. (2019). Medicine, 98(23), e15771. 10.1097/MD.0000000000015771

Yoon, S. J., Jung, J. G., Lee, S., Kim, J. S., Ahn, S. K., Shin, E. S., Jang, J. E., & Lim, S. H. (2020). The protective effect of alcohol consumption on the incidence of cardiovascular diseases: is it real? A systematic review and meta-analysis of studies conducted in community settings. BMC Public Health, 20(1), 90. 10.1186/s12889-019-7820-z

Yuan, S., Wu, J., Chen, J., Sun, Y., Burgess, S., Li, X., Akesson, A., & Larsson, S. C. (2024). Association between alcohol consumption and peripheral artery disease: two de novo prospective cohorts and a systematic review with meta-analysis. Eur J Prev Cardiol, Epub ahead of print. 10.1093/eurjpc/zwae142

Zhang, C., Qin, Y. Y., Chen, Q., Jiang, H., Chen, X. Z., Xu, C. L., Mao, P. J., He, J., & Zhou, Y. H. (2014). Alcohol intake and risk of stroke: a dose-response meta-analysis of prospective studies. International journal of cardiology, 174(3), 669–677. 10.1016/j.ijcard.2014.04.225

Zhang, H. Z., Shao, B., Wang, Q. Y., Wang, Y. H., Cao, Z. Z., Chen, L. L., Sun, J. Y., & Gu, M. F. (2022). Alcohol Consumption and Risk of Atrial Fibrillation: A Dose-Response Meta-Analysis of Prospective Studies. Frontiers in cardiovascular medicine, 9, 802163. 10.3389/fcvm.2022.802163

Zhang, X. Y., Shu, L., Si, C. J., Yu, X. L., Liao, D., Gao, W., Zhang, L., & Zheng, P. F. (2015). Dietary Patterns, Alcohol Consumption and Risk of Coronary Heart Disease in Adults: A Meta-Analysis. Nutrients, 7(8), 6582–6605. 10.3390/nu7085300

Zhao, J., Stockwell, T., Roemer, A., Naimi, T., & Chikritzhs, T. (2017). Alcohol Consumption and Mortality From Coronary Heart Disease: An Updated Meta-Analysis of Cohort Studies. Journal of studies on alcohol and drugs, 78(3), 375–386. https://www.jsad.com/doi/10.15288/jsad.2017.78.375

Zheng, Y. L., Lian, F., Shi, Q., Zhang, C., Chen, Y. W., Zhou, Y. H., & He, J. (2015). Alcohol intake and associated risk of major cardiovascular outcomes in women compared with men: a systematic review and meta-analysis of prospective observational studies. BMC Public Health, 15(1), 773. 10.1186/s12889-015-2081-y

